# Assessing physical fitness during pregnancy: validity and reliability of fitness tests, and relationship with maternal and neonatal health-related outcomes. A systematic review

**DOI:** 10.1101/2021.06.26.21259584

**Authors:** L. Romero-Gallardo, O Roldan-Reoyo, J. Castro-Piñero, L May, O. Ocón, V.A Aparicio, A Soriano-Maldonado

## Abstract

**Background:** Physical fitness (PF) is a powerful marker of health throughout the lifespan. In pregnant women, higher PF is associated with better maternal and fetal health, better delivery outcomes and earlier postpartum recovery. The assessment of PF during pregnancy requires special considerations to preserve fetal and maternal health; thus, providing a compilation of the most frequently used fitness tests, and assessing their validity, reliability, and association with maternal and neonatal health-related outcomes is of scientific and clinical interest.

**Objectives:** To systematically review studies evaluating one or more components of PF in pregnant women, to answer two research questions: 1) What fitness tests have been previously employed in pregnant women? and 2) What is the validity and reliability of these tests and their relationship with health-related outcomes?

**Data Sources:** PubMed and Web of Science.

**Methods:** Two independent reviewers systematically examined the articles in each database. The information from the included articles was summarized by a single researcher.

**Results:** A total of 149 articles containing a sum of 191 fitness tests were included. Among the 191 fitness tests, 99 (i.e.,52%) assessed cardiorespiratory fitness through 75 different protocols, 28 (15%) assessed muscular fitness through 16 different protocols, 14 (7%) assessed flexibility through 13 different protocols, 45 (24%) assessed balance through 40 different protocols, 2 assessed speed with the same protocol and 3 were multidimensional tests using one protocol. A total of 19 articles with 23 tests (13%) assessed either validity (n=4), reliability (n=6), or the relationship of PF with maternal and neonatal health-related outcomes (n=16).

**Conclusion:** PF during pregnancy has been assessed through a wide variety of protocols, mostly lacking validity and reliability data.

PROSPERO Registration Number: CRD42018117554

**Key points:**

- Physical Fitness during pregnancy has been assessed through a wide variety of tests (n=191) and different protocols (n=149).
- We found that cardiorespiratory fitness has been assessed with 75 different protocols (many of them made *ad hoc*), muscular fitness with 16 different protocols, flexibility with 13 different protocols, and balance with 40 different protocols.
- Most of the protocols lacked validity and reliability data, which limits the confidence on the association of fitness with maternal and neonatal health-related outcomes, which is still scarce.
- We advocate for an expert consensus to be developed in the following years to achieve the goal of appropriate and effective PF assessment during pregnancy.

**Running Heading:** Assessing physical fitness during pregnancy: validity, reliability and relationship with maternal and neonatal health-related outcomes.

## 1. Introduction

Physical Fitness has been defined as the ability to carry out daily tasks with vigor and alertness, without undue fatigue and with ample energy to enjoy leisure-time pursuits and meet unforeseen emergencies [1, 2]. Physical fitness (PF) is considered a powerful marker of health that is associated with a lower risk of cardiovascular events, cancer and all-cause mortality in all ages [3–7]. In pregnant women, some studies have recently underlined the potential impact of PF on maternal and fetal health [8–15]. Low PF levels are associated with low infant birth weight [8], increased risk of gestational diabetes mellitus [9, 10], poor postpartum recovery [11] and worse delivery outcomes [12, 13]. Moreover, the anatomical, biomechanical, physiological, and psychological changes during pregnancy might compromise PF levels [16–18]. Consequently, it is of clinical and public health interest to assess PF during pregnancy, and to understand what the best available tests to assess PF during this critical period of women’s life are.

Two categories of PF components have been defined: 1) *Health-related components* (cardiorespiratory fitness (CRF), muscular fitness, muscular endurance, and flexibility) 2) *Skill-related components* (ability, coordination, balance, power, reaction time and speed) [1, 2]. These PF components can be assessed subjectively through questionnaires [15], objectively and accurately through laboratory tests, and efficiently, economically and easily through field-based tests. During pregnancy, a wide variety of fitness tests have been used to assess PF; although a compilation of these tests has not been published to date. Compiling all fitness tests performed in pregnant women would help practitioners to select the most useful test according to their purpose. It is also important to note that, although laboratory tests are generally the gold standard for assessing PF, these tests are not generally accessible to everyone because they need sophisticated and expensive equipment, and it is not possible to evaluate a relatively large sample in a short period of time. As an alternative, a number of field test exist that provide an opportunity to assess PF in a more accessible way [2]. However, there is no consensus on which fitness tests should be used to assess PF in pregnant women, and the validity and reliability of many of the tests used to assess PF during pregnancy is unknown [19].

Since the assessment of PF in pregnant women requires special considerations to preserve fetal and maternal health [18,20,21], understanding which fitness tests are valid, reliable, and associated with maternal and neonatal health-related outcomes, would provide a framework for improving PF assessment during pregnancy and also for improving exercise prescription in this population.

The aims of this systematic review were to 1) describe which fitness tests have been used to evaluate PF in pregnant women and, 2) to evaluate the validity and reliability of the fitness tests, and their relationship with maternal and neonatal health-related outcomes.

## 2. Methods

### 2.1. Registration and Elaboration

This systematic review was prospectively registered at PROSPERO (CRD42018117554; available at http://www.t.ly/fS6a). In addition, the review followed the PRISMA explanation and elaboration [22] and the PRISMA Checklist [23] is included as supplementary material. See electronic supplementary material 1 (ESM 1).

### 2.2. Search Strategy

Articles were searched by two independent reviewers from two major databases, MEDLINE (PubMed) and the Web of Science (WOS) from inception until January 2021. The complete search strategy and further details are presented in ESM 2 (tables S1 and S2).

### 2.3. Inclusion Criteria

The inclusion criteria were: 1) Healthy pregnant women (no restriction regarding gestational week), 2) At least one component of PF assessed either through field-based or laboratory tests, 3) Access to full text, 4) Only one original article from the same study/project using the same test were included, and 5) Text in English or Spanish.

### 2.4. Quality assessment of the articles

To assess the quality of the articles included for aim 2, we used three quality scores that are comprehensively described in ESM 3 and ESM 4.

### 2.5. Process and data extraction

After checking title and abstract, only the studies meeting all inclusion criteria were introduced in a reference manager software (Mendeley). In the event of disagreement between the two independent reviewers concerning the inclusion/exclusion of an article, a consensus was reached (there was no need of a third person). The snowball strategy was also used. Information including reference, age, sample size and fitness test description are summarized in ESM 5-Table S6.

## 3. Results

A comprehensive PRISMA flow diagram is presented in Figure 1.

**Figure 1.**
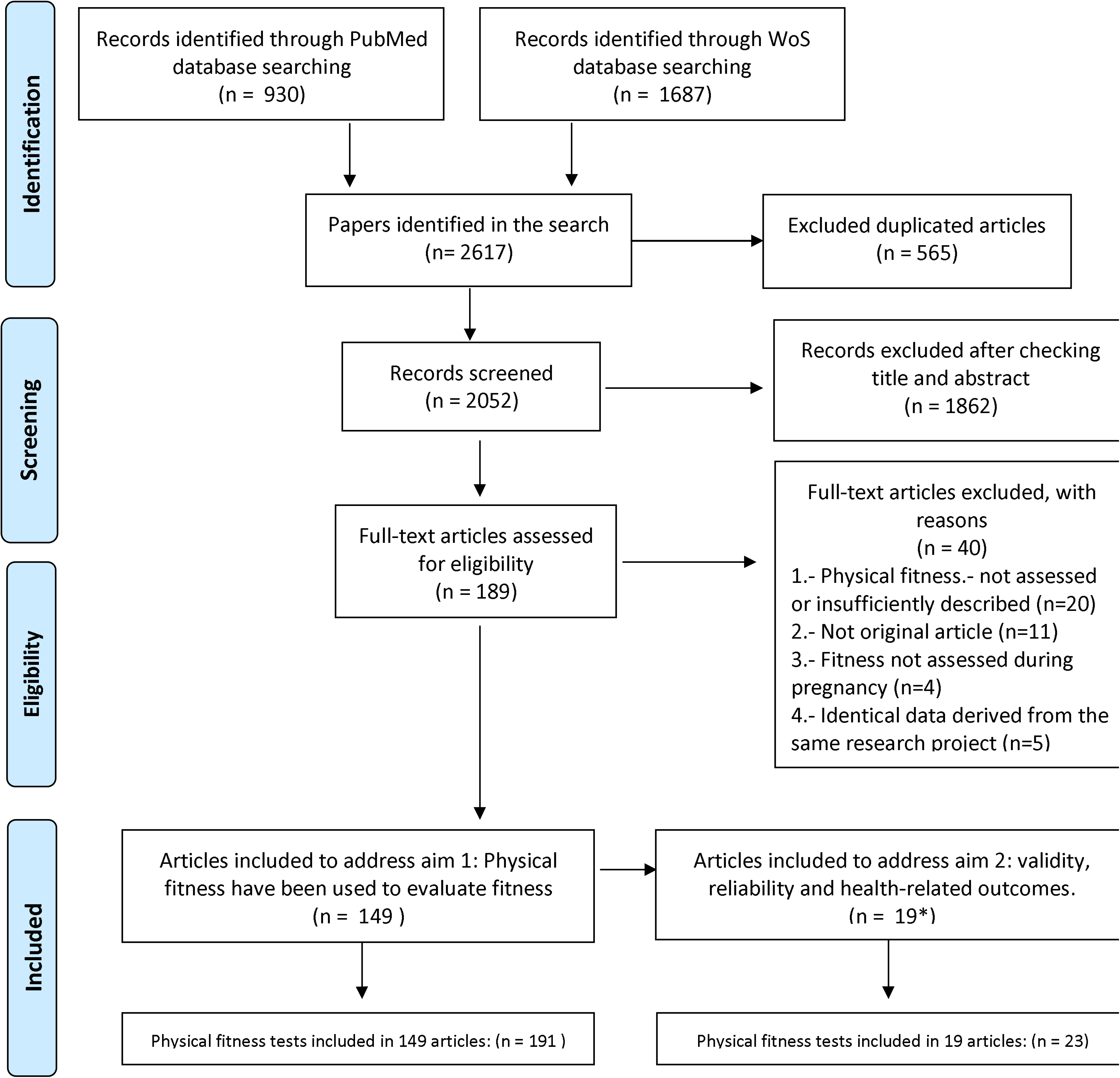
Flow chart of the literature search and paper selection process.

### 3.1. Overall results, quality assessment and gestational week

We identified 2617 studies, of which 149 were included (Figure 1). These articles contained 191 fitness tests, using 149 different protocols that were included for aim 1. A comprehensive scheme of the fitness tests and the different protocols performed to date, divided by PF component, is presented in figure 2. A summary of the number and percentage of articles that assessed PF during pregnancy and protocols used for its assessment, divided by PF components, is presented in table 1.

**Table 1.** Number (%) of articles that assessed the different components of physical fitness during pregnancy and protocols used for its assessment.

|  | **TOTAL** | **CRF** | **MF** | **Flexibility** | **Balance** | **Speed** | **Multidimensional** |
| --- | --- | --- | --- | --- | --- | --- | --- |
| **Fitness tests** | 191 | 99 (52) | 28 (15) | 14 (7) | 45 (24) | 2 (1) | 3 (1) |
| **Protocols** | 149 | 75 (50) | 16 (11) | 13 (9) | 40 (29) | 1 (0.5) | 1 (0.5) |

CRF: Cardio-respiratory fitness; MF: Muscular Fitness

**Figure 2.**
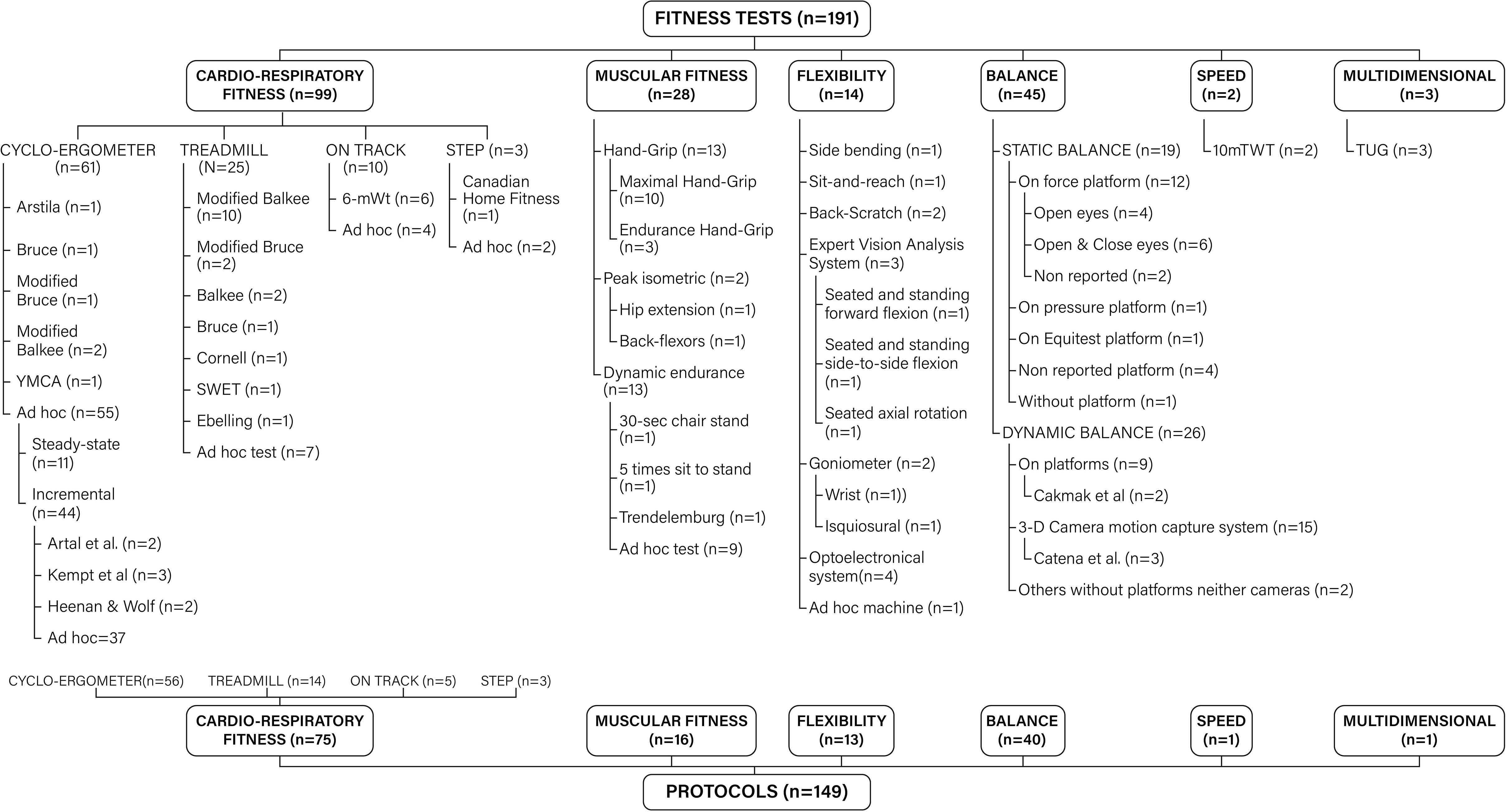
Scheme of the fitness tests and the different protocols divided by PF component.

Regarding aim 1, 99 tests (including 75 different protocols) were used to assess cardiorespiratory fitness, 28 (including 16 different protocols) to assess muscular fitness, 14 (including 13 different protocols) to assess flexibility, 45 tests (including 40 different protocols) to assess balance, 2 tests using the same protocol to assess speed, and 3 tests using the same protocol were multidimensional. No results were found for other PF components such as agility or coordination.

Regarding aim 2, a total of 19 articles (13% of the total number of articles included) assessed at least validity (n=3), reliability (n=4) of fitness tests, or the relationship of PF with maternal and neonatal health-related outcomes (n=16). Of these 16 articles, 11 were classified as very low quality [13,28–36] and 5 were classified as low quality [8,33,37–39] (table 2). Of the 3 articles [40–42] that assessed validity, 2 articles were classified as low quality [40, 41] and 1 as high quality [42]. Of the 4 articles that assessed reliability criteria, 3 were considered high quality [40,43,44] and 1 low quality [45] (table 2).

**Table 2.**
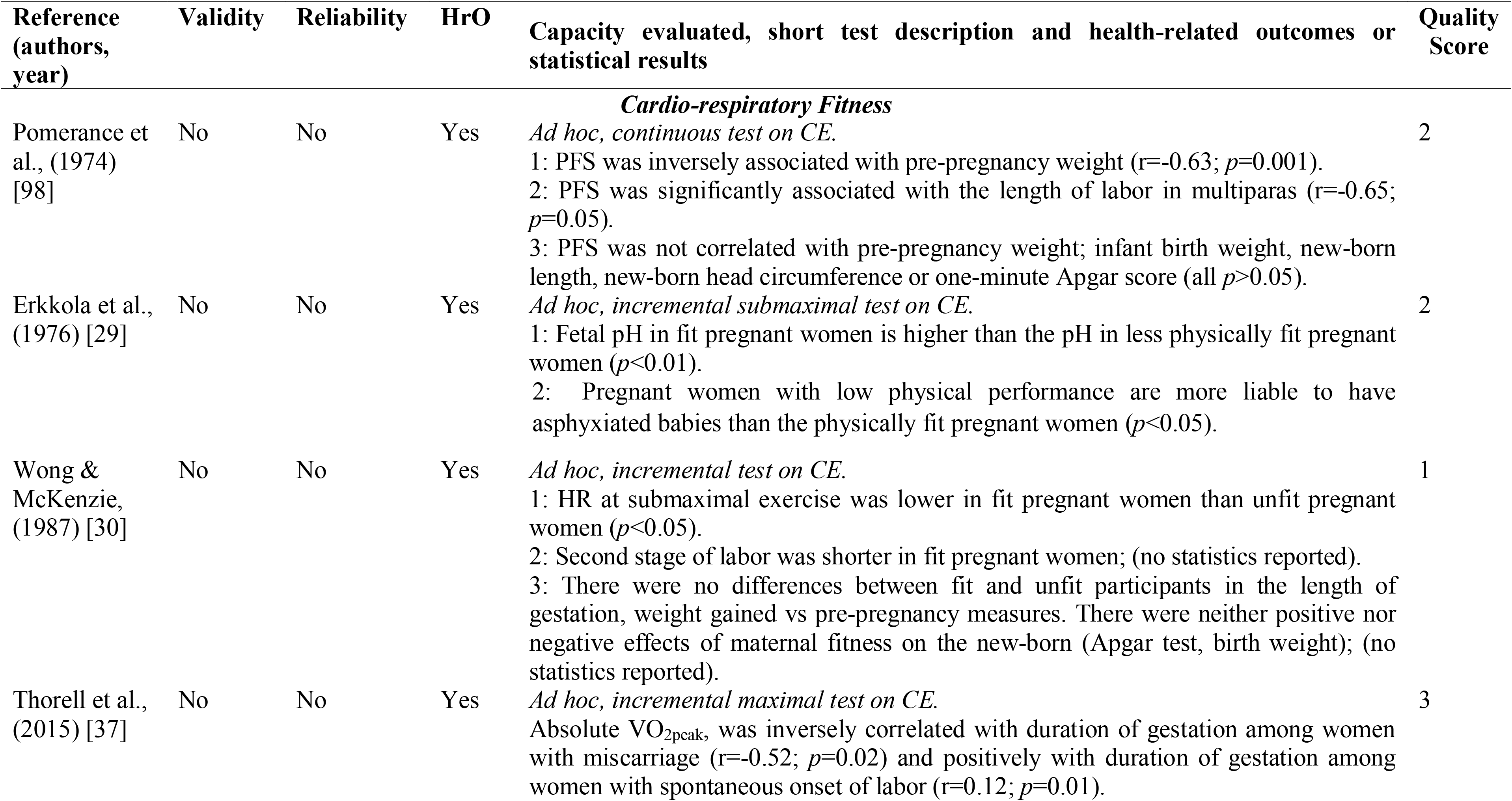
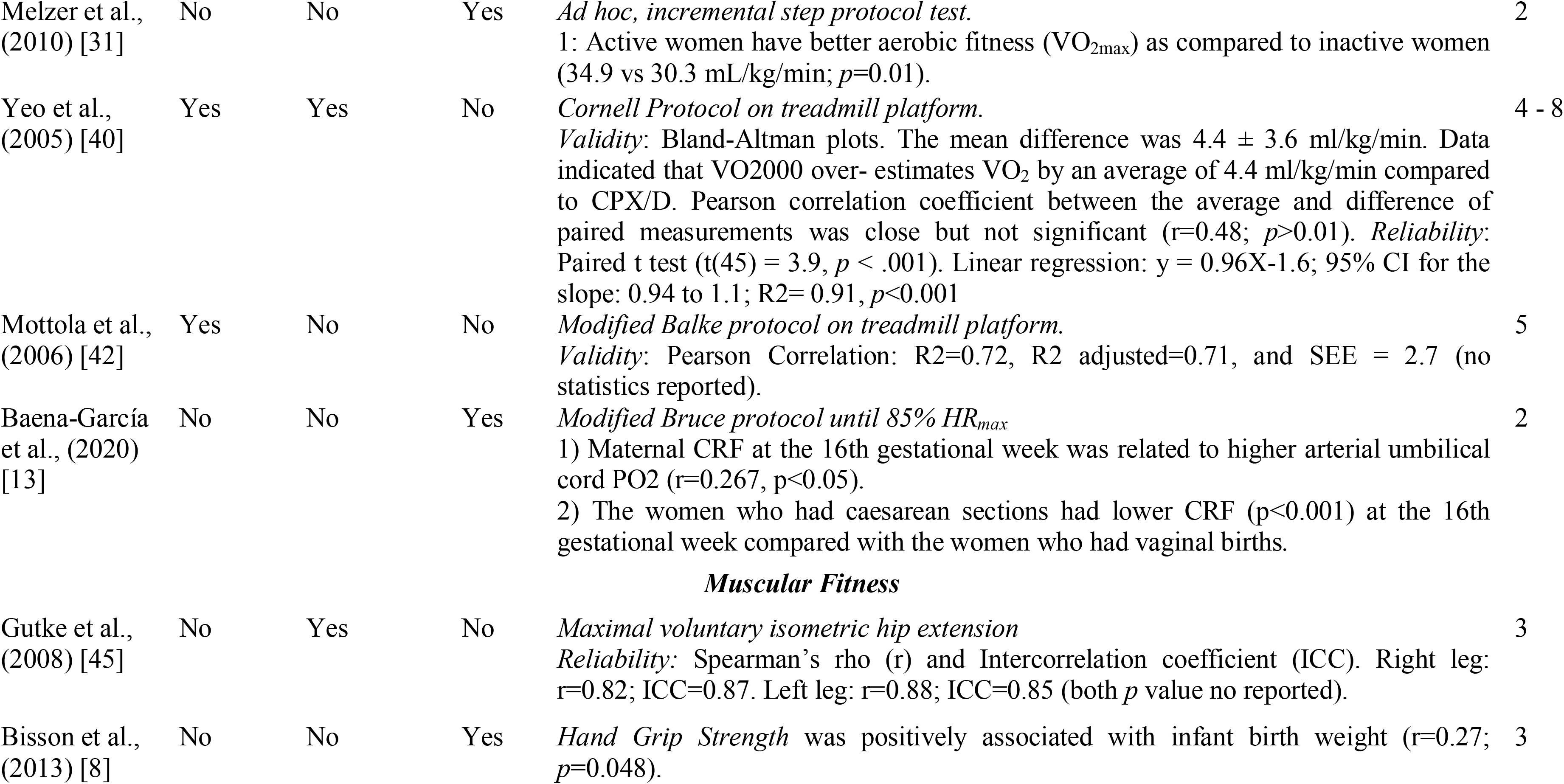
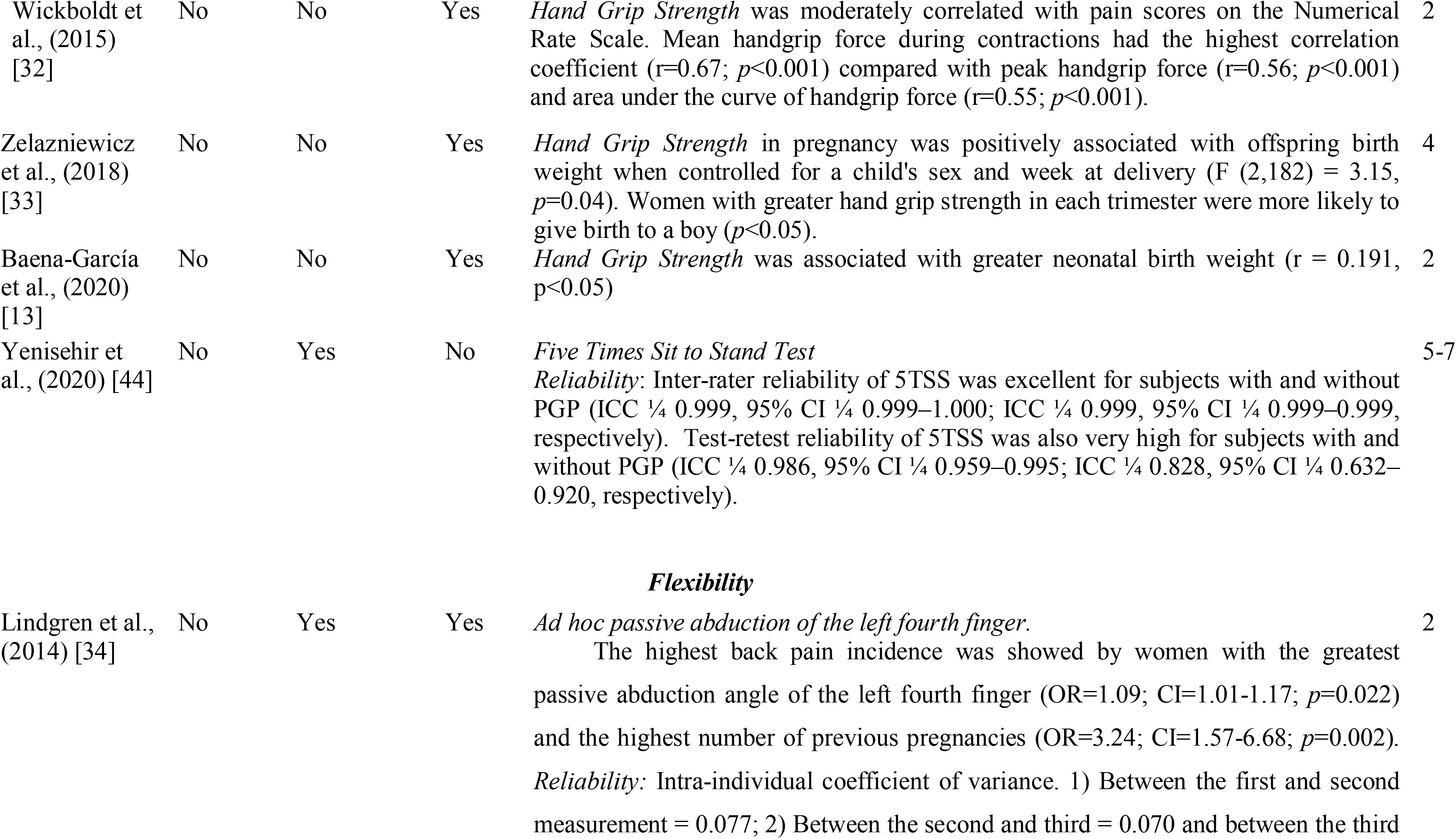
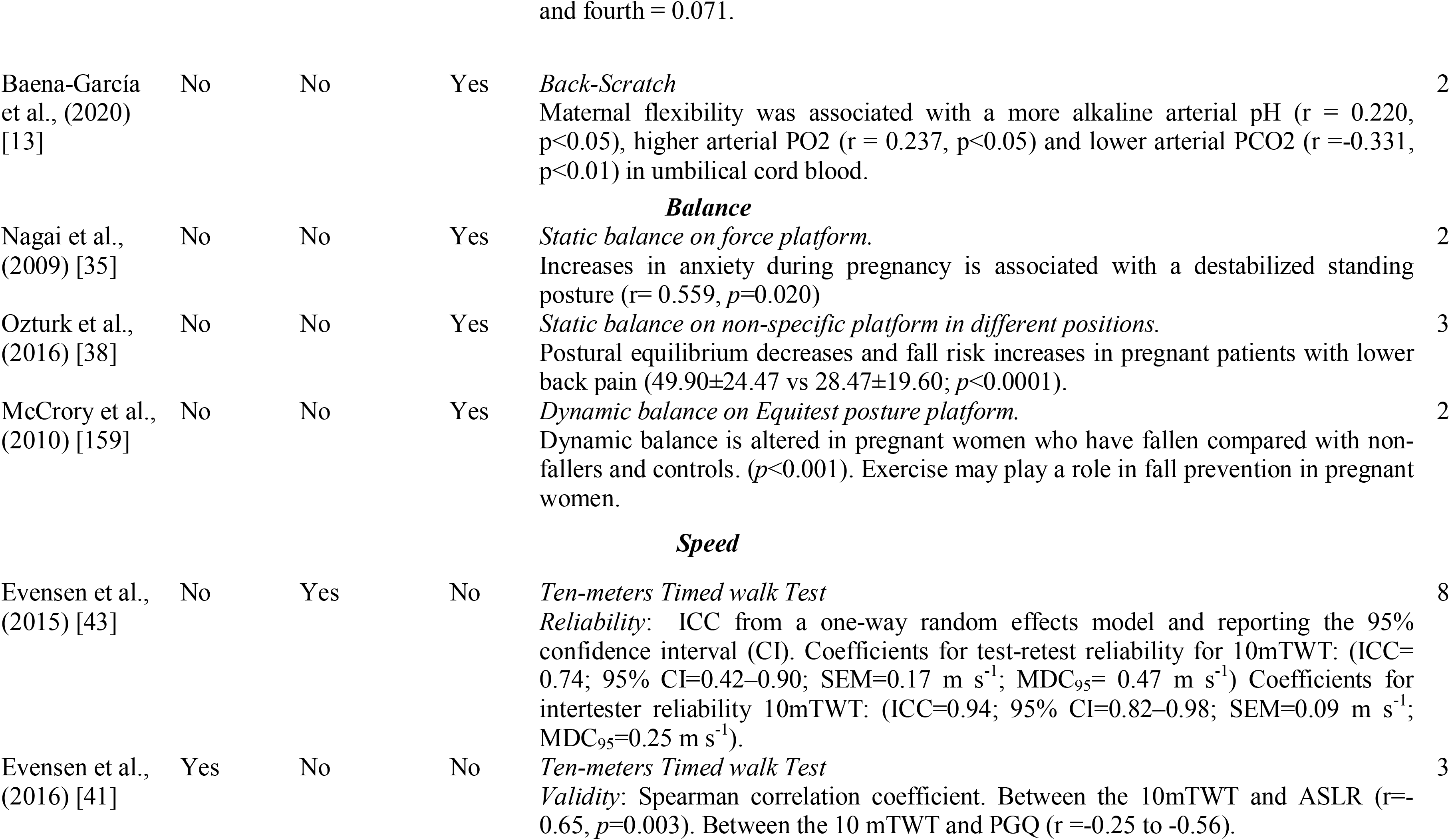
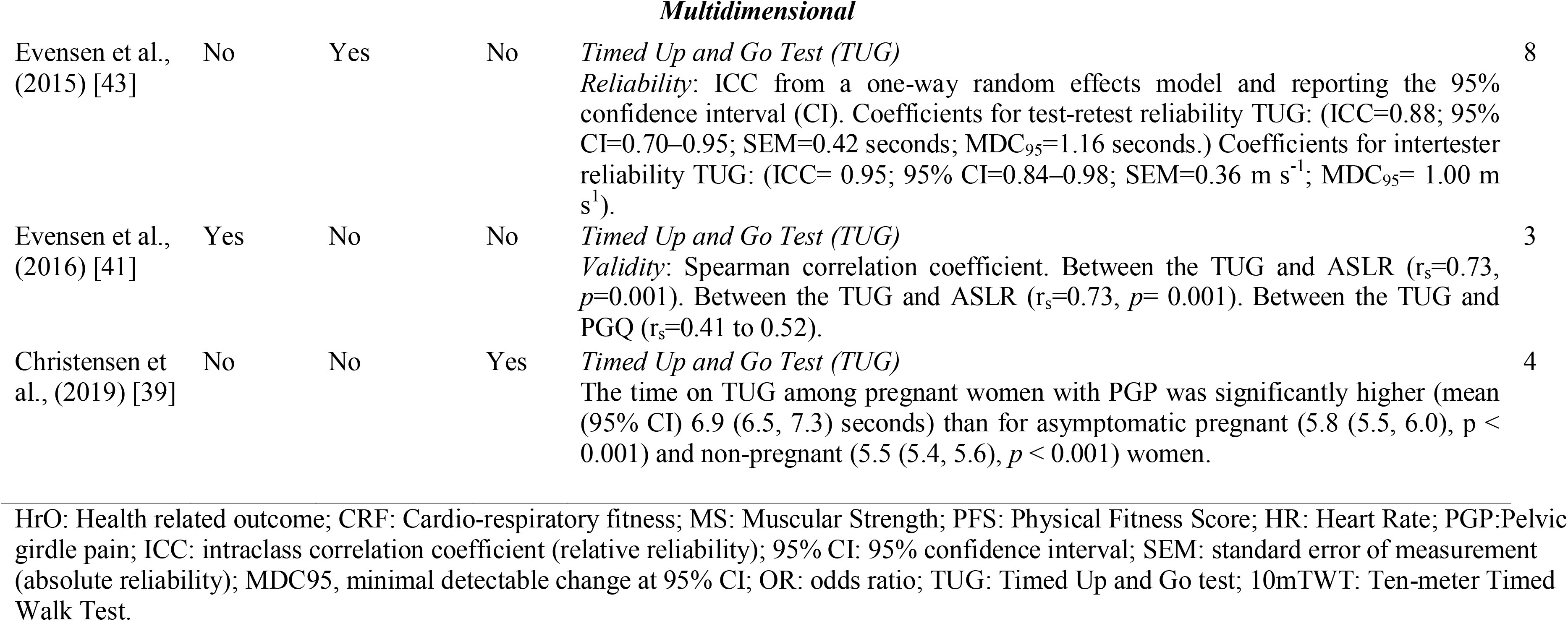
Overview of studies that assessed the validity and/or reliability of fitness tests during pregnancy or the association of physical fitness with health-related outcomes (HrO) in pregnant women.

The gestational week at PF assessment ranged from 8 to 41 across articles. Some articles assessed PF at different time points throughout pregnancy; therefore, we divided pregnancy into two stages. Early pregnancy (i.e., from week 0 to week 20 of gestation) and late pregnancy (i.e., from week 21 to week 40). Using this approach, 11 articles (7%) were performed in early pregnancy; 57 articles (38%) were performed in late pregnancy; 55 articles (37%) were performed several times (i.e., range 2 to 5 times) throughout pregnancy; 7 (5%) articles specified a range of weeks that included early to late pregnancy; 14 articles (9%) reported only the trimester without specifying gestational week; 4 articles (3%) provided no information, and 1 article (1%) assessed PF on the day of labor.

### 3.2. Cardiorespiratory fitness

#### 3.2.1. Tests used

We identified 99 tests assessing CRF, of which 61 (62%) were performed on cycle-ergometer, 25 (25%) on treadmill, 10 (10%) on track and there were 3 (3%) step protocols (figure 2). Of the 99 tests, a total of 75 corresponded to different protocols. For instance, there were 56 different protocols using *cycle-ergometer*, distributed as follows: only one article used the Arstila test [29]; one used the Bruce Protocol at 75% HR_max_ [46]; one applied the Modified Bruce ramp protocol at anaerobic threshold [47]; two employed the Modified Balke protocol at 70% HR_max_ [48, 49]; one used YMCA protocol [50]; and the remaining articles (n=55) used ad-hoc tests (i.e. specifically designed for the purpose of the investigation); 11 of which [28,49,51–57] used steady-state tests and 44 [29,30,62–71,37,72–81,47,82–91,49,92–96,50,58–61] used incremental tests. When analyzing the type of test based on intensity, we found that 13 articles used maximal tests [47,63,93,94,97,69,70,72,73,75,84,85,90], 37 submaximal tests [29,30,59,61,62,65–68,71,76,78,37,79–83,87–89,91,95,50,96,98,99,51,53,54,56–58] and 3 used mixed tests [49,60,64] containing submaximal and maximal stages within the same protocol.

There were 25 *treadmill* tests that used 14 different protocols, distributed as follows: the Modified Balke protocol was used in 10 articles [8,42,63,100–106]; the Modified Bruce protocol in 2 articles [13, 107]; and the traditional Balke protocol -twice in the same article- [108]; the traditional Bruce protocol [109], the Cornell protocol [40], the SWET protocol and the Ebbelling single-stage protocol [18] were each used in 1 article. There were other 7 *ad hoc* tests of which 2 were steady-state [54, 110], and 5 were incremental tests [58,111–114]. According to intensity, 3 were maximal tests [110,112,113] and 4 submaximal tests [54,58,111,114].

Of the 10 tests *on track*, 6 articles performed the 6-minute walk test protocol [115–120], and 4 were *ad hoc* tests (i.e. 1 maximal and 4 were submaximal). In regards to the 3 *step tests*, 1 Canadian Home Fitness test [121] was used and 2 *ad hoc* incremental submaximal tests were used [31, 122].

### 3.2.2. Validity and reliability

We identified 2 articles examining validity [40, 42]. Yeo et al. [40] aimed to validate a portable metabolic testing system (VO2000) on healthy sedentary pregnant women. The VO2000 consistently overestimated VO_2_ measurements, compared to the same manufacturer’s reference system, by 4.4±3.6 standard deviation (SD) ml/kg/min although the Pearson correlation was significant (r=0.48; *p*=0.01). When VO2000 was used twice, the mean difference was statistically significant (1.0±1.8 ml/kg/min; t(45)=3.9, *p*<0.001). Mottola et al. [42] provided a prediction equation for VO_2peak_ in pregnant women between 16 and 22 weeks of gestation, using a modified Balke protocol. The results of this equation revealed an adjusted R^2^ of 0.71 (*p* value not reported) When the authors used this equation to predict VO_2peak_ in a cross-validation group (n=39), they found a predicted value of 23.38±4.03 mL×kg ^-1^×min^-1^, while the actual value was 23.54 ± 5.9 mL×kg ^-1^×min^-1^ (*p* value not reported).

### 3.2.3. Relationship with maternal and neonatal health-related outcomes

A total of 6 articles analyzed the association of CRF with maternal and neonatal health-related outcomes. Pomerance et al. [28] observed that VO_2max_ was inversely associated with the length of labor in multiparas (r=-0.65; *p*=0.001) and pre-pregnancy weight (r=-0.63; *p*=0.001). However, VO_2max_ was neither correlated with newborn weight, length, or head circumference, nor with the one-minute Apgar score (all *p*>0.05). In the same line, Wong & McKenzie [30], observed that fit mothers showed lower HR at submaximal exercise intensity (p<0.05) and the second stage of labor was shorter (no statistics reported) compared to unfit pregnant mothers. However, there was no difference between fit and unfit in the length of gestation or weight gained (no statistics reported). In the same article, the authors showed neither positive nor negative effect of maternal fitness on newborn weight or Apgar scores.

In addition, Erkkola et al. [29] found that newborn from fit pregnant women had higher pH than fetuses of less physically fit women (*p*<0.01). In this article, participants with low physical performance were more likely to have asphyxiated neonates than neonates of physically fit women (*p*<0.05). In the same line, Baena-García et al. [13] observed that maternal CRF at the 16th gestational week was related to higher arterial umbilical cord PO_2_ (r=0.267, p<0.05), and women who had caesarean sections had significantly lower CRF compared with the women who had vaginal births (p<0.001).

On the other hand, Bisson et al. [8] studied the association of CRF in early pregnancy with physical activity before and during early pregnancy. The authors found that a higher VO_2 peak_ in early pregnancy was positively associated with physical activity spent at sports and exercise before and during early pregnancy (p<0.001).

### 3.3. Muscular fitness

#### 3.3.1. Tests used

A total of 28 tests (i.e., 14% of all included articles) that included 16 different protocols assessed muscular fitness, of which 10 performed maximal hand-grip strength tests [8,12,128,13,32,33,123–127], 3 performed endurance hand-grip test, 2 for 3-min [129, 130] and 1 for 2-min period [131] (figure 2). In 2 of the articles conducting an endurance hand- grip test [129, 131], a hand grip sphygmomanometer was used instead of dynamometry. On the other hand, 1 used hand-held dynamometer fixed to a chair to assess quadriceps strength [132] and 1 used toe-grip dynamometer [132]. Moreover, 2 *ad hoc* isometric tests were used to assess maximal voluntary hip extension and back flexors endurance in the same article [133]. Finally, 13 dynamic endurance tests were found, 9 were listed as *ad hoc* tests [12,126,134] and other 3 (30-sec Chair Stand Test, 5 Times Sit to Stand test, Trendelenburg’s test) were classified as others dynamic tests [13,44,126].

#### 3.3.2. Validity and reliability

Only 2 muscular fitness tests assessed reliability [44, 45]. Yenisehir et al. [44] analyzed reliability and validity of Five Times Sit-to-Stand. Inter-rater reliability was excellent for subjects with and without pelvic girdle pain (ICC = 0.999, 95% CI = 0.999–1.000; ICC = 0.999, 95% CI = 0.999–0.999, respectively). Test-retest reliability was also very high for subjects with and without pelvic girdle pain (PGP) (ICC = 0.986, 95% CI = 0.959–0.995; ICC = 0.828, 95% CI = 0.632–0.920, respectively).

On the one hand, Gutke et al.[45] analyzed the reliability for an *ad hoc* test. This test consisted in a maximal voluntary isometric hip extension with a fixed sensor holding a sling around the thigh and pulling for 5 seconds during 3 reps with 5-10 seconds of rest (r=0.82 for the right leg and r=0.88 for the left leg; ICC=0.87 for the right leg and 0.85 for the left leg; with *p* value no reported).

#### 3.3.3. Relationship with maternal and neonatal health-related outcomes

Bisson et al.[8] observed that hand-grip strength was positively associated with infant birth weight (r=0.34, *p*=0.0068) even after adjustment for confounders (r=0.27, *p*=0.0480). Zelazniewicz et al. [33] observed that hand-grip strength was associated with offspring birth weight when controlled for the newborn sex and gestational age at delivery (F(2.182)=3.15; *p*=0.04). Baena-García et al. [13] found greater hand-grip strength weakly associated with greater neonatal birth weight (r = 0.191, p<0.05). Wickboldt [32] found that hand-grip strength was moderately correlated with pain scores, where the mean hand-grip strength during contractions had the highest correlation coefficient (r=0.67; *p*<0.001) compared with peak handgrip strength (r=0.56; *p*<0.001) and the area under the curve of handgrip force (r=0.55; *p*<0.001).

### 3.4. Flexibility

#### 3.4.1. Tests used

Our search identified 14 (7%) tests that assessed flexibility using 13 different protocols, including the side bending test [135], the sit-and-reach test [12], the back-scratch test (twice) [13, 125], the Motion Analysis (i.e. including 3 different tests such as the seated and standing forward flexion, seated and standing side to side flexion and seated axial rotation [136]) and optoelectrical system (i.e. performing 4 different tests) [137]. Goniometry was used in two different articles to measure isquiosural flexibility, [138] wrist flexion-extension and medial lateral deviation [139]. Only one article used an *ad hoc* machine to test passive abduction of the left fourth finger [34].

#### 3.4.2. Validity and reliability

Lindgren et al.[34] designed an *ad hoc* machine to test passive abduction of the left fourth finger and its relationship with low-back pain during pregnancy and early postpartum. Abduction angle was measured at three different times throughout the pregnancy and once in the postnatal period. Reliability of the abduction angle was analyzed by the intra-individual coefficient of variance. The coefficients of variance between the first and second measurement was 0.077, between the second and third 0.070 and between the third and fourth 0.071.

#### 3.4.3. Relationship with maternal and neonatal health-related outcomes

Only 2 flexibility tests evaluated associations with maternal and neonatal health-related outcomes. Lindgren et al. [34] found that women with greater passive abduction angle of the left fourth finger was associated with highest back pain incidence (OR=1.09; CI=1.01-1.17; *p*=0.022) and the highest number of previous pregnancies (OR=3.24; CI=1.57-6.68; *p*=0.002). Baena-Garcia et al. [13] found increased flexibility associated with a more alkaline arterial pH (r = 0.220, p<0.05), higher arterial PO_2_ (r = 0.237, p<0.05) and lower arterial PCO_2_ (r =- 0.331, p<0.01) in the umbilical cord blood.

### 3.5. Balance

#### 3.5.1. Tests used

We identified 45 (24%) articles assessing balance of which 19 analyzed static balance and 26 dynamic balance trough 40 different protocols. With regard to *static balance*, 18 were laboratory tests of which 12 assessed balance through stabilometry tests on force platform [35,140,149,150,141–148], one on pressures platform [143] and another on Equitest® platform [151]. Four articles did not mention the type of platform used (40,150,176,177). Regarding protocols, all articles conducted the tests with participants standing with bipedal support. However, standing position varied between articles. Ten articles maintained a standing posture with feet separated [35,132,145,147–153], 1 with feet together [146], 2 used mixed protocols [38, 142], 1 with medial malleoli separated [154] and 4 did not mention the standing posture [140,141,143,144]. Moreover, 3 articles used protocols with eyes open [141,148,150] exclusively, 8 articles used mixed protocols with eyes open and closed, 1 used visual target and visual tasks [144] and 6 did not specify whether participants kept their eyes closed or opened. Only 1 article used a field test, the one-legged standing protocol [125]. On the other hand, 1 test was field-test without platform.

In relation to the 26 articles measuring *dynamic balance*, 9 of them assessed balance using platforms. Each of these articles used different testing tool such as balance master platform [155], pressure platform [143], force platform [156], Equitest® platform [36] and movable platform, which was used in two articles [157, 158]. Two of these articles were walking protocols [143, 156], 1 with translational perturbations [159], one was standing with one knee flexed and arms across the chest [157, 158]. Another 15 articles used 3-D camera motion capture systems using 13 different protocols. Twelve of the 15 articles were walking protocols [160,161,170–172,162–169] and 2 used a stand to sit motion protocol [173, 174]. Moreover, 1 article used a triaxial accelerometer[175]; another article assessed balance through recording (without specification of camera type) [176], and another using instrumented insoles [177]. All three articles used walking protocols.

#### 3.5.2. Validity and reliability

No validity or reliability assessments were performed regarding balance tests.

#### 3.5.3. Relationship with maternal and neonatal health-related outcomes

3 articles associated balance with neonatal and maternal health-related outcomes. Ozturk [38] observed that static balance decreased and fall risk increased in pregnant women with lower back pain (49.90±24.47 vs 28.47±19.60; *p*<0.0001). In relation to exercise, McCrory et al. [159] showed that exercise may play a role in fall prevention in pregnant women (*p*=0.005); they also found that dynamic balance is altered in pregnant women who have fallen compared with non-fallers and non-pregnant women (*p*<0.001). Nagai et al. [35] studied the relationship between anxiety and balance. They concluded that when anxiety increases during pregnancy, the standing posture is destabilized (r=0.559, *p*=0.020), which may increase the chance of falling.

### 3.6. Speed

#### 3.6.1. Test used

The only protocol that was used to assess speed during pregnancy was the ten-meters timed walk test (10mTWT). However, the same test was identified in two different articles. [41, 43]. In the 10mTWT, the participants commenced standing at a chair. When told to start, subjects walk as fast as possible along 14 meters marked with white blank tapes placed at 0 m, 2 m, 12 m and 14 m. The time (100th of a second) required to walk between the 2 m and 12 m markers was recorded and converted into speed in meters per second (m/sec).

#### 3.6.2. Validity and reliability

Validity and reliability for 10mTWT was studied by Evensen et al. in two different articles [41, 43]. In 2015, Evensen al. [43] analyzed the test-retest reliability of 10mTWT showing an intraclass correlation coefficient (ICC) of (0.74). Intertester reliability was determined in the first 13 participants with strong correlation (ICC =0.94). In 2016 [41] the same authors analyzed the convergent validity of 10mTWT by comparing performances with scores achieved on the Active Straight Leg Raise (ASLR) test and observed moderate positive correlations between 10mTWT and ASLR (r=0.65, *p*=0.003).

#### 3.6.3. Relationship with maternal and neonatal health-related outcomes

This systematic review did not find any articles that analyzed the association of speed with maternal and neonatal health-related outcomes.

### 3.7. Agility and coordination

No articles of agility and coordination were identified.

### 3.8. Multidimensional

#### 3.8.1. Test used

Our search identified a walking multidimensional test that was used in three studies [39,41,43]. In Timed Up and Go Test (TUG) the participant begins seated in a chair with their arms on armrests and their toes against a start line. The purpose is to cross the front white line at three meters away, turn around, and walk back to the chair and sit down as fast as possible. The performance is measured in time (100th of a second).

#### 3.8.2. Validity and reliability

Validity and reliability for TUG was analyzed by Evensen et al. in two different studies [41, 43]. The TUG showed good test-retest reliability (ICC=0.88) and intertester reliability (ICC=0.95). Regarding reliability, strong correlations were found between the TUG and Active Straight Leg Raise (r= 0.73, *p*= 0.001).

#### 3.8.3. Relationship with maternal and neonatal health-related outcomes

The time on TUG among pregnant women with pelvic girdle pain was significantly higher (mean (95% CI) 6.9 (6.5, 7.3) seconds) than for asymptomatic pregnant (5.8 (5.5, 6.0), p < 0.001) and non-pregnant (5.5 (5.4, 5.6), *p* < 0.001) women.

## 4. Discussion

### 4.1. Summary of the evidence

This systematic review revealed that PF has been assessed through a wide variety of tests during pregnancy. However, very little is known on the validity and reliability of the tests performed. These findings have important research and clinical implications. First, until a specific battery of fitness assessments for pregnant women is developed and validated, the confidence in PF data during pregnancy is limited and potentially unreliable and may prove harmful if unreliable values are used for exercise recommendation during pregnancy. Second, the large variety of tests used makes is challenging to compare results from different studies. Third, provided the lack of rigorous information on validity and reliability of PF tests, it is also difficult to evaluate the association of PF with maternal and neonatal health-related outcomes, which is of wide clinical and public health interest. However, some studies have attempted to present associations of PF with maternal and neonatal health-related outcomes, which undoubtedly needs to be replicated once a PF test battery is released. Before that, exhaustive research must be performed in validating such battery of tests.

#### 4.1.1. Cardiorespiratory Fitness

This systematic review identified that cycle-ergometer has been the equipment most frequently used to assess CRF followed by treadmill and field tests; although step tests have also been conducted. There is a large disparity of protocols and wide variety of *ad hoc* tests used, which makes comparing results between studies difficult. However, the Modified Balke treadmill Protocol validated by Mottola et al. [42] for pregnant women has been the most frequently used test. There have been more incremental tests used for CRF tests during pregnancy compared to steady-state tests and more submaximal compared to maximal tests. There is no consensus regarding test termination criteria for submaximal tests, which undoubtedly needs further research. Some articles used relative intensity using physiological variables such as %HR_max_ or %VO_2max,_ and other used absolute intensity, such as specific HR (beats per minute). Among the studies that used %HR_max_ as a test termination criterion, there was a variety of percentages such as 70% [48,58,65,96], 75% [46,61,91,107] or 85% [13,40,81]. Among the studies that used %VO_2max_, there were different percentages such as 40% [54], 50% [53, 59], 60% [52, 54], or 70% [178]. Among the studies that used absolute HR as a test termination criterion, the HR for finalizing the tests were set either at 125 [86], 150 [30,56,66,87], 155 [122], 160 [89], or 170 [77,78,82,83] beats per minute. Some studies even used the rate perceived exertion as complementary criteria [50,71,78] or peak aerobic power [67]. These complementary criteria have been recommended and studied in pregnant women by authors like Hesse et al. [109] since the physical and emotional changes during pregnancy limit performance. It must be noted that the same equation was not used to estimate HR_max_. Some articles used the traditional 220-age formula [61,65,81,91,107] while others used the Karvonen [40] or Tanaka [96] formulas. Some articles did not specify how HR_max_ was estimated [46,48,58]. This heterogeneity could be due to the physiological complexity of the pregnant woman, in terms of cardiac changes and response to exercise and the lack of scientific information in this regard. Moreover, the gestational week could be a determinant for physiological response since Bijl et al. [96] observed a slower hemodynamic recovery and an increased ventilatory response to exercise in early pregnancy compared to non-pregnant women. With regards to the maximal tests, different terms have been used like maximal criteria such as volitional fatigue [64,69,70,72,73,93,94,109,178], exhaustion [63], anaerobic threshold [47,75,112,113], and point of symptom limitation [84,85,95].

This lack of consensus has many drawbacks that need to be resolved in view of the need to accurately assess CRF during pregnancy. We advocate for an expert consensus to be developed in the following years to achieve the goal of appropriate and effective CRF assessment during pregnancy. In particular, it seems essential to develop a treadmill and a cycle ergometer submaximal test that reveals sufficient validity to confidently estimate VO_2max_ throughout gestation.

#### 4.1.2. Muscular fitness

Muscular fitness tests included muscular strength, endurance and power [2]. The studies included in this systematic review show that muscular strength was the most frequently assessed component of muscular fitness, since only 6 studies [12,13,44,126,134,179] assessed endurance and none of them assessed power in pregnant women. In most studies, muscular strength was evaluated through handgrip maximal strength using a dynamometer. However, 2 studies used a handgrip sphygmomanometer test [129, 131]. Some of the handgrip tests were performed in standing position [8, 123], while others used sitting position [125] or supine position [127], and others did not reveal the position used for the assessment [33,124,126,138]. Some tests were completed 3 times [126], others twice [8,33,124], and others only once [125,127,138]. This clearly reveals a large methodological variability that might influence the results and make comparing results between studies difficult. Another limitation is the fact that the main strength outcome was handgrip strength. While handgrip strength is a good marker of health [180], it is unclear whether handgrip responds to changes following exercise interventions. Therefore, validating other muscular strength tests, including lower limb strength tests, is needed in order for researchers and practitioners to confidently assess muscular strength during pregnancy.

There were no validity studies and the reliability was assessed only in one maximal isometric hip extension test [45]. This test has limitations since the pregnant abdomen must be on a bed and, as acknowledged by the authors, it cannot be performed during the third trimester. It must be noted that higher handgrip strength was associated with higher birth weight [8, 33]. Moreover, increased hand-grip strength was produced during uterine contraction [32]. The advantage of using handgrip is that it represents an inexpensive, rapid, and easy-to-use assessment with minimal training needed to appropriately administer. However, assessing the performance of pregnant athletes with this test seems clearly insufficient. More quality in tests employed is necessary since the association of muscular strength with maternal and neonatal health-related outcomes is of clinical importance. Moreover, other studies are needed to understand the extent to which preserving strength throughout pregnancy and postpartum relates to clinical outcomes.

#### 4.1.3. Flexibility

Although there were 7 studies assessing flexibility, none of them used the same protocol. Once again, this reflects a lack of agreement when assessing the same component of PF. Moreover, Lindgren et al. [34] found that a higher flexibility showed a higher low back pain. Despite the limitation of a finger laxity test, we considered these findings an interesting association that warrants further article since passive stretching is one of the most common practical prescriptions for exercise professionals instead of mobility and breathing exercises. On the other hand, the results of Baena-García et al. [13] are very relevant to foetal health since the flexibility was associated with a better pH, PO_2_ and PCO_2_ in umbilical cord blood. Hence, more research about flexibility tests, their outcomes and their prescription are needed.

#### 4.1.4. Balance

We identified that balance was the second PF component most frequently evaluated during pregnancy, following CRF. This makes sense since the center of gravity changes during pregnancy as a result of expansion of the uterus and the risk of falls increases. However, there is a great heterogeneity between the protocols employed in different studies. For *static balance*, the protocol most frequently used was stabilometry on force platform with bipedal support and eyes open and eyes closed within the same test [35,38,154,181,182,140– 144,146,147,151]. For *dynamic balance*, there was a greater heterogeneity across protocols both in the platform used and in the movements over the platforms. Regarding the assessment tool, the 3-D camera was the device most frequently used [151,160–162,164,173]. Likewise, we observed differences between the number of platform pieces, trials and Hz utilized. Some protocols were performed on 2 piece-platforms [141,147,154], others on 1 piece-platforms [35,140,142,145,146,148,152] and others did not specify the type of platform [143,144,151]. Although the number of trials and the frequency of recording (i.e. Hz) are important parameters that should be carefully described about the protocols, only 5 (out of 13) articles described the number of trials [140,145,147,152,183] and 1 described frequency of recording [141]. The usefulness of these tests are restricted to the research area and all of them use expensive technological tools; therefore, it is difficult to extrapolate these tests to fitness centers or clinical settings. We could prevent falls during pregnancy if we could assess balance easily. For this reason, it is necessary to develop an inexpensive and easy-to-use balance field-test.

### 4.2. Validity, reliability and maternal and neonatal health-related outcomes

Unfortunately, studies that examine validity and reliability of PF tests are scarce. The physical fitness component most frequently studied was CRF, however we have only found two studies that analyzed the validity of the CRF tests and no studies examined the reliability of these tests. On treadmill platform, Mottola et al. [42], validated a special equation for modified Balke protocol that has been used by numerous other authors. In contrast, Yeo tried to validate a portable metabolic testing system (mod. VO2000) but it overestimated VO_2_ measurements for pregnant women compared to non-pregnant women and males.

Regarding muscular fitness, hand grip test was most commonly used; this test was used as the gold standard for muscular fitness during pregnancy. Only Gutke et al. [45], studied the reliability of a test for hip extension. However, the *p* value was not reported, and the position adopted in the test could be uncomfortable for pregnant participants. Finally, the studies evaluating validity and reliability of speed tests and multidimensional components of PF have been researched by Evensen et al. [41, 43]. They demonstrated that TUG and 10mTWT are reliable and valid tests for pregnant women.

The validity and reliability of balance (without tests), agility and/or coordination tests has not been investigated to date.

We suggest that specific tests to be performed in pregnant women are needed and their validity and reliability must be assessed to understand the extent to which one might rely on such measures when prescribing exercise, or making clinical recommendations.

Regarding maternal and neonatal health-related outcomes, we can conclude that more research is also necessary. Nevertheless, from this review we can highlight some interesting associations with different fitness components. A better CRF was associated with a shorter labor [30, 98] and a lower risk of cesarean section [13]. However, no association was found regarding other fetal outcomes such as Apgar scores or the newborn anthropometrics [30, 98]. By contrast, muscular strength was associated with optimum infant birth weight [8,13,33]. Other neonatal outcomes like fetal umbilical cord pH were positively associated with maternal CRF [29]. On the other hand, better balance scores are associated with decreased fall risk [35,184,185]. These results are very useful for exercise professionals, as it implies that protocols during pregnancy must be implemented with balance exercises. Finally, Evensen et al.[39] found that pelvic girdle pain could be a limiting factor to assess physical fitness in pregnant women since the time of TUG was significantly higher in women with pain than in asymptomatic pregnant and non-pregnant women.

None of the studies reviewed in this article have described adverse events during PF assessment. Moreover, official bodies such as the American College of Obstetrician and Gynaecology or the Canadian Society of Exercise Physiology have highlighted the benefits of an adequate PF assessment, and assert the need of consensus in the PF assessment during pregnancy [186].

### 4.3. Limitations and Strengths

A limitation of this article is that, although PubMed and WOS are among the most relevant databases in the medical literature, the possibility that a small number of studies have been overlooked cannot be discarded. Nevertheless, these two databases are the biggest databases in sports medicine and sports sciences and, therefore, include the vast majority of studies.

A strength of this systematic review is the fact that, to the best of our knowledge, this is the first article to comprehensively analyze PF assessments, the validity and reliability of fitness tests, and their relationship with maternal and neonatal health-related outcomes during pregnancy. The results from this systematic review provide an overall picture of how PF is being assessed in this population, what type of tests are being performed, their specific characteristics, whether these tests have been tested for validity and/or reliability; and whether PF is associated with maternal and neonatal health-related outcomes. All this information is of wide and undoubted clinical interest.

## 5. Conclusions

The main finding of this systematic review is that PF has been assessed through a wide variety of protocols, mostly lacking validity and reliability data, and that no consensus exists on the most suitable fitness tests to be performed during pregnancy. In addition, information regarding the association of PF with maternal and neonatal health-related outcomes is scarce and should further evaluated as well. Provided the need to assess PF during pregnancy and the importance not only to understand the physical state of the pregnant women but also to precisely prescribe exercise in this population, extensive research is needed to design and validate a battery of fitness tests to be used for the safe and effective assessment of PF during pregnancy. We advocate for an expert consensus to be developed in the following years to achieve the goal of appropriate and effective PF assessment during pregnancy.

## Supporting information

ESM1_PRISMA_Checklist

ESM2_TableS1_TableS2

ESM3_Description_quality_score

ESM4_TableS3_S4_S5

ESM5_TableS6

## Data Availability

This article is a systematic review and their results can be consulted in the electronic supplementary material at the same article

## AUTHORS CONTRUBITIONS

Conceptualization: Romero-Gallardo, L.; Aparicio, V.; Castro-Piñero, J.; Soriano-Maldonado, A.

Literature search and data analysis: Romero-Gallardo, L.; Roldán-Reoyo, O. Methodology: Romero-Gallardo, L.; Roldán-Reoyo, O.; Castro-Piñero,J.; Soriano-Maldonado, A.

Formal analysis and investigation: Romero-Gallardo, L.; Roldán-Reoyo, O.; Soriano-Maldonado, A. Writing - original draft preparation: Romero-Gallardo, L.

Writing - review and editing: Romero-Gallardo, L.; Roldan-Reoyo, O; Castro-Piñero, J.; Aparicio, A.; May, L; Ocón, O., Soriano-Maldonado, A.

Resources: Aparicio, V., Castro-Piñero, J.; Soriano-Maldonado, A. Supervision: Aparicio, V.; Ocón O.; Castro-Piñedo, J.; Soriano-Maldonado, A.

## FUNDING

This study has been partially funded by the University of Granada, Plan Propio de Investigación 2016, Excellence actions: Units of Excellence: Unit of Excellence on Exercise and Health (UCEES), and by the Junta de Andalucía, Consejería de Conocimiento, Investigación y Universidades and European Regional Development Fund (ERDF), ref. SOMM17/6107/UGR. Current research activities of Dr. Alberto Soriano-Maldonado are supported by a grant from the Spanish Ministry of Science, Innovation and Universities (ref. RTI2018-093302-A-I00). This study is of the Doctoral Thesis of Lidia Romero Gallardo, within the Biomedicine Doctoral Program at the University of Granada.

## CONFLICT OF INTEREST

Authors declare that they have no conflict of interest.

## Appendix

### Electronic Supplementary Material (ESM)

**ESM 1:** PRISMA Checklist

**ESM 2:** Electronic Supplementary Material 2

**- Table S1.** Search strategy used and number of articles found in Pubmed.

**- Table S2.** Search strategy used and number of articles found in Web of Science.

**ESM 3: Comprehensive description of the three quality assessment scores used in the present systematic review.**

**ESM 4:** Electronic Supplementary Material 4

**- Table S3.** Quality assessment criteria to evaluate validity and reliability studies.

**- Table S4.** Quality assessment criteria to evaluate reliability studies.

**- Table S5.** Quality assessment criteria to evaluate health-related outcomes studies.

**ESM 5:** Electronic Supplementary Material 5

**- Table S6.** Overview of studies included in the systematic review and description of physical fitness tests.

## Notes

### Competing Interest Statement

The authors have declared no competing interest.

### Clinical Trial

PROSPERO (CRD42018117554; available at http://www.t.ly/fS6a)

### Clinical Protocols

http://www.t.ly/fS6a

### Funding Statement

This study has been partially funded by the University of Granada, Plan Propio de Investigacion 2016, Excellence actions: Units of Excellence: Unit of Excellence on Exercise and Health (UCEES), and by the Junta de Andalucia, Consejeria de Conocimiento, Investigacion y Universidades and European Regional Development Fund (ERDF), ref. SOMM17/6107/UGR. Current research activities of Dr. Alberto Soriano-Maldonado are supported by a grant from the Spanish Ministry of Science, Innovation and Universities (ref. RTI2018-093302-A-I00). This study is of the Doctoral Thesis of Lidia Romero Gallardo, within the Biomedicine Doctoral Program at the University of Granada.

### Author Declarations

This article is a systematic review.

## REFERENCES

1. Caspersen CJ, Christenson GM. Physical Activity, Exercise, and Physical Fitness: Definitions and Distinctions for Health-Related Research. Public Health Rep [Internet]. 1985 [cited 2020 Oct 30];100(April):126–31. Available from: /pmc/articles/PMC1424733/?report=abstract

2. American College of Sports Medicine, In Riebe D, In Ehrman JK, In Liguori G, In Magal M. ACSM’s guidelines for exercise testing and prescription.10th edition. 2018.

3. Blair S, Kohl H, Barlow C. Changes in physical fitness and allcause mortality. A prospective study of healthy and unhealthy men. JAMA. 1995;273:1093–8.

4. Kodama S, Saito K, Tanaka S, Miko M, Yoko Y, Asumi M, et al. Cardiorespiratory Fitness as a Quantitative Predictor of All-Cause Mortality and Cardiovascular Events. JAMA. 2009;301(19):2024–35.

5. Gibbons LW, Blair SN, Cooper KH, Smith M. Association between coronary heart disease risk factors and physical fitness in healthy adult women. Circulation [Internet]. 1983 May 1 [cited 2018 May 4];67(5):977–83. Available from: http://circ.ahajournals.org/cgi/doi/10.1161/01.CIR.67.5.977

6. Ortega F, Cadenas-Sanchez C, Lee D, Ruiz J, Blair S, Sui X. Fitness and Fatness as Health Markers through the Lifespan: An Overview of Current Knowledge. Prog prev med [Internet]. 2018;3(2):e0013. Available from: http://ovidsp.ovid.com/ovidweb.cgi?T=JS&PAGE=reference&D=ovftt&NEWS=N&AN=01960908-201804000-00001

7. Ortega FB, Ruiz JR, Castillo MJ, Sjöström M. Physical fitness in childhood and adolescence: A powerful marker of health. Int J Obes. 2008;32(1):1–11.

8. Bisson M, Alméras N, Plaisance J, Rhéaume C, Bujold E, Tremblay A, et al. Maternal fitness at the onset of the second trimester of pregnancy: Correlates and relationship with infant birth weight. Pediatr Obes [Internet]. 2013 Dec 1 [cited 2018 Nov 12];8(6):464–74. Available from: http://doi.wiley.com/10.1111/j.2047-6310.2012.00129.x

9. Gar C, Rottenkolber M, Grallert H, Banning F, Freibothe I, Sacco V, et al. Physical fitness and plasma leptin in women with recent gestational diabetes. Hu C, editor. PLoS One [Internet]. 2017 Jun 13 [cited 2018 Nov 10];12(6):e0179128. Available from: https://dx.plos.org/10.1371/journal.pone.0179128

10. Engberg E, Tikkanen HO, Koponen A, Hägglund H, Kukkonen-Harjula K, Tiitinen A, et al. Cardiorespiratory fitness and health-related quality of life in women at risk for gestational diabetes. Scand J Med Sci Sports [Internet]. 2017;(April):1–9. Available from: http://doi.wiley.com/10.1111/sms.12896

11. Weissgerber TL, Wolfe LA, Davies GAL, Mottola MF. Exercise in the prevention and treatment of maternal – fetal disease : a review of the literature. 2006;674:661–74.

12. Price BB, Amini SB, Kappeler K. Exercise in Pregnancy: Effect on Fitness and Obstetric Outcomes-A Randomized Trial. Med Sci Sports Exerc [Internet]. 2012 [cited 2018 Nov 12];44(12):2263–9. Available from: http://www.acsm-msse.org

13. Baena-García L, Coll-Risco I, Ocón-Hernández O, Romero-Gallardo L, Acosta- Manzano P, May L, et al. Association of objectively measured physical fitness during pregnancy with maternal and neonatal outcomes. The GESTAFIT Project. Luo Z-C, editor. PLoS One [Internet]. 2020 Feb 18 [cited 2020 Jul 23];15(2):e0229079. Available from: https://dx.plos.org/10.1371/journal.pone.0229079

14. Marín-Jiménez N, Acosta-Manzano P, Borges-Cosic M, Baena-García L, Coll-Risco I, Romero-Gallardo L, et al. Association of self-reported physical fitness with pain during pregnancy: The GESTAFIT Project. Scand J Med Sci Sport. 2019;29(7):1022–30.

15. Romero-Gallardo L, Soriano-Maldonado A, Ocón-Hernández O, Acosta-Manzano P, Coll-Risco I, Borges-Cosic M, et al. International Fitness Scale—IFIS: Validity and association with health-related quality of life in pregnant women. Scand J Med Sci Sport. 2019;(October):1–10.

16. Miller MJ, Kutcher J, Adams KL. Effect of Pregnancy on Performance of a Standardized Physical Fitness Test. Mil Med [Internet]. 2017;182(11):e1859–63. Available from: http://militarymedicine.amsus.org/doi/10.7205/MILMED-D-17-00093%0Ahttp://www.ncbi.nlm.nih.gov/pubmed/29087853

17. Treuth MS, Butte NF, Puyau M. Pregnancy-Related Changes in Physical Activity, Fitness, and Strength. Med Sci Sport Exerc [Internet]. 2005 [cited 2018 Nov 13];37(5):832–7. Available from: http://www.acsm-msse.org

18. LeMoyne EL, Curnier D, Ellemberg D. Pregnancy and cognition: Deficits in inhibition are unrelated to changes in fitness. J Clin Exp Neuropsychol [Internet]. 2014 Feb 7 [cited 2018 Nov 10];36(2):178–85. Available from: http://www.ncbi.nlm.nih.gov/pubmed/24479656

19. Meah VL, Backx K, Davenport MH, International Working Group on Maternal Hemodynamics. Functional hemodynamic testing in pregnancy: recommendations of the International Working Group on Maternal Hemodynamics. Ultrasound Obstet Gynecol Off J Int Soc Ultrasound Obstet Gynecol [Internet]. 2018 Mar [cited 2018 Nov 10];51(3):331–40. Available from: http://www.ncbi.nlm.nih.gov/pubmed/28857365

20. May LE, Allen JJB, Gustafson KM. Fetal and maternal cardiac responses to physical activity and exercise during pregnancy. Early Hum Dev [Internet]. 2016;94:49–52. Available from: http://dx.doi.org/10.1016/j.earlhumdev.2016.01.005

21. Wolfe LA, Weissgerber TTL. Clinical physiology of exercise in pregnancy: a literature review. J Obstet Gynaecol Can [Internet]. 2003 Jun [cited 2018 Nov 13];25(6):473–83. Available from: http://www.ncbi.nlm.nih.gov/pubmed/12806449

22. Liberati A, Altman DG, Tetzlaff J, Mulrow C, Gøtzsche PC, Ioannidis JPA, et al. The PRISMA Statement for Reporting Systematic Reviews and Meta-Analyses of Studies That Evaluate Health Care Interventions: Explanation and Elaboration. PLoS Med [Internet]. 2009 Jul 21;6(7):e1000100. Available from: https://dx.plos.org/10.1371/journal.pmed.1000100

23. Moher D, Liberati A, Tetzlaff J, Altman DG. Preferred Reporting Items for Systematic Reviews and Meta-Analyses: The PRISMA Statement. PLoS Med [Internet]. 2009 Jul 21;6(7):e1000097. Available from: https://dx.plos.org/10.1371/journal.pmed.1000097

24. Castro-Piñero J, Artero EG, España-Romero V, Ortega FB, Sjöström M, Suni J, et al. Criterion-related validity of field-based fitness tests in youth: A systematic review [Internet]. Vol. 44, British Journal of Sports Medicine. Br J Sports Med; 2010 [cited 2020 Oct 30]. p. 934–43. Available from: https://pubmed.ncbi.nlm.nih.gov/19364756/

25. Artero EG, Espaa-Romero V, Castro-Piero J, Ortega FB, Suni J, Castillo-Garzon MJ, et al. Reliability of field-based fitness tests in youth [Internet]. Vol. 32, International Journal of Sports Medicine. Int J Sports Med; 2011 [cited 2020 Oct 30]. p. 159–69. Available from: https://pubmed.ncbi.nlm.nih.gov/21165805/

26. Armijo-Olivo S, Stiles CR, Hagen NA, Biondo PD, Cummings GG. Assessment of study quality for systematic reviews: A comparison of the Cochrane Collaboration Risk of Bias Tool and the Effective Public Health Practice Project Quality Assessment Tool: Methodological research. J Eval Clin Pract [Internet]. 2012 Feb [cited 2020 Oct 30];18(1):12–8. Available from: https://pubmed.ncbi.nlm.nih.gov/20698919/

27. Rodriguez-Ayllon M, Cadenas-Sánchez C, Estévez-López F, Muñoz NE, Mora-Gonzalez J, Migueles JH, et al. Role of Physical Activity and Sedentary Behavior in the Mental Health of Preschoolers, Children and Adolescents: A Systematic Review and Meta-Analysis. Sport Med [Internet]. 2019;49(9):1383–410. Available from: https://doi.org/10.1007/s40279-019-01099-5

28. Pomerance JJ, Gluck L, Lynch VA. Maternal exercise as a screening test for uteroplacental insufficiency. Obstet Gynecol [Internet]. 1974 Sep [cited 2018 Nov 14];44(3):383–7. Available from: http://www.ncbi.nlm.nih.gov/pubmed/4853105

29. Erkkola R, Rauramo L. Correlation of Maternal Physical Fitness During Pregnancy with Maternal and Fetal Ph and Lactic Acid at Delivery. Acta Obstet Gynecol Scand [Internet]. 1976 Jan 1 [cited 2018 Nov 14];55(5):441–6. Available from: http://doi.wiley.com/10.3109/00016347609158527

30. Wong SC, McKenzie DC. Cardiorespiratory fitness during pregnancy and its effect on outcome. Int J Sports Med [Internet]. 1987 Apr 14 [cited 2018 Nov 14];8(2):79–83. Available from: http://www.thieme-connect.de/DOI/DOI?10.1055/s-2008-1025645

31. Melzer K, Schutz Y, Soehnchen N, Othenin-Girard V, Martinez de Tejada B, Irion O, et al. Effects of recommended levels of physical activity on pregnancy outcomes. Am J Obstet Gynecol [Internet]. 2010 Mar 1 [cited 2018 Nov 12];202(3):266.e1-266.e6. Available from: https://www.sciencedirect.com/science/article/pii/S0002937809020924?via%3Dihub

32. Wickboldt N, Savoldelli G, Rehberg-Klug B. Continuous assessment of labour pain using handgrip force. Pain Res Manag [Internet]. 2015 [cited 2018 Nov 10];20(3):159–63. Available from: http://www.ncbi.nlm.nih.gov/pubmed/25996768

33. Żelaźniewicz A, Pawłowski B. Maternal hand grip strength in pregnancy, newborn sex and birth weight. Early Hum Dev [Internet]. 2018 Apr 1 [cited 2018 Nov 26];119:51–5. Available from: https://www.sciencedirect.com/science/article/pii/S0378378218300847?via%3Dihub

34. Lindgren A, Kristiansson P. Finger joint laxity, number of previous pregnancies and pregnancy induced back pain in a cohort study. BMC Pregnancy Childbirth [Internet]. 2014 Feb 6 [cited 2018 Nov 10];14(1):61. Available from: http://www.ncbi.nlm.nih.gov/pubmed/24507564

35. Nagai M, Isida M, Saitoh J, Hirata Y, Natori H, Wada M. Characteristics of the control of standing posture during pregnancy. Neurosci Lett [Internet]. 2009 Sep 22 [cited 2018 Nov 12];462(2):130–4. Available from: https://www.sciencedirect.com/science/article/pii/S0304394009008908?via%3Dihub

36. McCrory JLL, Chambers AJJ, Daftary A, Redfern MSS. Dynamic postural stability during advancing pregnancy. J Biomech [Internet]. 2010 Aug 26 [cited 2018 Nov 12];43(12):2434–9. Available from: http://www.ncbi.nlm.nih.gov/pubmed/20537334

37. Thorell E, Goldsmith L, Weiss G, Kristiansson P. Physical fitness, serum relaxin and duration of gestation. BMC Pregnancy Childbirth [Internet]. 2015;15(1):1–7. Available from: http://dx.doi.org/10.1186/s12884-015-0607-z

38. Öztürk G, Geler Külcü D, Aydoğ E, Kaspar Ç, Uğurel B. Effects of lower back pain on postural equilibrium and fall risk during the third trimester of pregnancy. J Matern Neonatal Med [Internet]. 2016 Apr 17 [cited 2018 Nov 10];29(8):1358–62. Available from: http://www.tandfonline.com/doi/full/10.3109/14767058.2015.1049148

39. Christensen L, Vollestad NK, Veierod MB, Stuge B, Cabri J, Robinson HS. The Timed Up & Go test in pregnant women with pelvic girdle pain compared to asymptomatic pregnant and non-pregnant women. Musculoskelet Sci Pract. 2019 Oct;43:110–6.

40. Yeo S, Ronis DLDL, Antonakos CLCL, Roberts K, Hayashi R. Need for population specific validation of a portable metabolic testing system: A case of sedentary pregnant women. J Nurs Meas [Internet]. 2005 [cited 2018 Nov 13];13(3):207–18. Available from: 10.1139/y03-072

41. Evensen NM, Kvåle A, Brækken IH. Convergent validity of the Timed Up and Go Test and Ten-metre Timed Walk Test in pregnant women with pelvic girdle pain. Man Ther [Internet]. 2016 Feb 1 [cited 2018 Nov 10];21:94–9. Available from: https://www.sciencedirect.com/science/article/pii/S1356689X15001319

42. Mottola MF, Davenport MH, Brun CR, Inglis SD, Charlesworth S, Sopper MM. VO2peak prediction and exercise prescription for pregnant women. Med Sci Sports Exerc. 2006;38(8):1389–95.

43. Evensen NM, Kvale A, Braekken IH, Kvåle A, Braekken IH. Reliability of the Timed Up and Go test and Ten-Metre Timed Walk Test in Pregnant Women with Pelvic Girdle Pain. Physiother Res Int [Internet]. 2015 Sep [cited 2018 Nov 26];20(3):158–65. Available from: http://doi.wiley.com/10.1002/pri.1609

44. Yeni ehir S, Çıtak Karakaya İ, Sivaslıoğlu AA, Özen Oruk D, Karakaya MG. Reliability and validity of Five Times Sit to Stand Test in pregnancy-related pelvic girdle pain. Musculoskelet Sci Pract [Internet]. 2020 Aug;48:102157. Available from: https://linkinghub.elsevier.com/retrieve/pii/S2468781219304047

45. Gutke A, Östgaard H, Öberg B. Association between muscle function and low back pain in relation to pregnancy. J Rehabil Med [Internet]. 2008 [cited 2018 Nov 13];40(4):304–11. Available from: https://medicaljournals.se/jrm/content/abstract/10.2340/16501977-0170

46. Kulpa PJ, White BM, Visscher R. Aerobic exercise in pregnancy. Am J Obstet Gynecol [Internet]. 1987 Jun 1 [cited 2018 Nov 16];156(6):1395–403. Available from: https://www.sciencedirect.com/science/article/pii/0002937887900068?via%3Dihub

47. Bilodeau J-F, Bisson M, Larose J, Pronovost E, Brien M, Greffard K, et al. Physical fitness is associated with prostaglandin F-2 alpha isomers during pregnancy. PROSTAGLANDINS Leukot Essent Fat ACIDS. 2019 Jun;145:7–14.

48. Field SK, Bell SG, Cenaiko DF, Whitelaw WA. Relationship between inspiratory effort and breathlessness in pregnancy. J Appl Physiol [Internet]. 1991 Nov [cited 2018 Nov 27];71(5):1897–902. Available from: http://www.physiology.org/doi/10.1152/jappl.1991.71.5.1897

49. Soultanakis HN, Artal R, Wiswell RA. Prolonged exercise in pregnancy: glucose homeostasis, ventilatory and cardiovascular responses. Semin Perinatol [Internet]. 1996 Aug 1 [cited 2018 Nov 14];20(4):315–27. Available from: https://www.sciencedirect.com/science/article/pii/S0146000596800243?via%3Dihub

50. Sussman D, Saini BS, Schneiderman JE, Spitzer R, Seed M, Lye SJ, et al. Uterine artery and umbilical vein blood flow are unaffected by moderate habitual physical activity during pregnancy. Prenat Diagn. 2019 Oct;39(11):976–85.

51. Morton MJ, Paul MS, Campos GR, Hart M V., Metcalfe J. Exercise dynamics in late gestation: Effects of physical training. Am J Obstet Gynecol [Internet]. 1985 May 1 [cited 2018 Nov 14];152(1):91–7. Available from: https://www.sciencedirect.com/science/article/pii/S0002937885801873?via%3Dihub

52. Hume RFFJ, Bowie JDD, McCoy C, Magarelli PCC, Gall M, Hertzberg BSS, et al. Fetal umbilical artery Doppler response to graded maternal aerobic exercise and subsequent maternal mean arterial blood pressure: predictive value for pregnancy-induced hypertension. Am J Obstet Gynecol [Internet]. 1990 Sep 1 [cited 2018 Nov 14];163(3):826–9. Available from: https://www.sciencedirect.com/science/article/pii/000293789091077P?via%3Dihub

53. Young JC, Treadway JL. The effect of prior exercise on oral glucose tolerance in late gestational women. Eur J Appl Physiol Occup Physiol [Internet]. 1992;64(5):130–3. Available from: http://link.springer.com/10.1007/BF00625062

54. Clapp JF, Little KD, Capeless EL. Fetal heart rate response to sustained recreational exercise. Am J Obstet Gynecol [Internet]. 1993 Jan 1 [cited 2018 Nov 14];168(1):198–206. Available from: https://linkinghub.elsevier.com/retrieve/pii/S000293781290914X

55. O’Neill ME, Cooper KA, Hunyor SN, Boyce S. Cardiorespiratory response to walking in trained and sedentary pregnant women. J Sports Med Phys Fitness [Internet]. 1993 Mar [cited 2018 Nov 27];33(1):40–3. Available from: http://www.ncbi.nlm.nih.gov/pubmed/8350606

56. Mcgrath MJ, Wolfe LA, Preston RJ, Burggraf GW, Mcgrath MJ. Effects of pregnancy and chronic exercise on maternal cardiac structure and function. Artic Can J Physiol Pharmacol [Internet]. 1999 Nov [cited 2018 Nov 13];77(11):909–17. Available from: https://www.researchgate.net/publication/12707078

57. Kim J-H, Roberge RJ, Powell JB. Effect of external airflow resistive load on postural and exercise-associated cardiovascular and pulmonary responses in pregnancy: a case control study. BMC Pregnancy Childbirth [Internet]. 2015 Feb 22 [cited 2018 Nov 10];15(1):45. Available from: http://www.ncbi.nlm.nih.gov/pubmed/25886031

58. Veille J-CC, Hohimer ARR, Burry K, Speroff L. The effect of exercise on uterine activity in the last eight weeks of pregnancy. Am J Obstet Gynecol [Internet]. 1985 Mar 15 [cited 2018 Nov 14];151(6):727–30. Available from: https://www.sciencedirect.com/science/article/pii/0002937885905058?via%3Dihub

59. Jovanovic L, Kessler A, Peterson CM. Human maternal and fetal response to graded exercise. J Appl Physiol [Internet]. 1985 May [cited 2018 Nov 14];58(5):1719–22. Available from: http://www.ncbi.nlm.nih.gov/pubmed/3888949

60. Carpenter MW, Sady SP, Hoegsberg B, Sady MA, Haydon B, Cullinane EM, et al. Fetal heart rate response to maternal exertion. JAMA J Am Med Assoc [Internet]. 1988 May 27 [cited 2018 Nov 14];259(20):3006–9. Available from: http://jama.jamanetwork.com/article.aspx?doi=10.1001/jama.1988.03720200028028

61. Moore DH, Jarrett JC 2nd, Bendick PJ. Exercise-induced changes in uterine artery blood flow, as measured by Doppler ultrasound, in pregnant subjects. Am J Perinatol [Internet]. 1988 Apr 4 [cited 2018 Nov 16];5(2):94–7. Available from: http://www.ncbi.nlm.nih.gov/pubmed/3279974

62. Sady SP, Carpenter MW, Sady MA, Haydon B, Hoegsberg B, Cullinane EM, et al. Prediction of VO2max during cycle exercise in pregnant women Prediction of VO 2 max during cycle exercise in pregnant women. J Appl Physiol. 1988;65(2):657–61.

63. Artal R, Masaki DI, Khodiguian N, Romem Y, Rutherford SE, Wiswell RA. Exercise prescription in pregnancy: weight-bearing versus non-weight-bearing exercise. Am J Obstet Gynecol [Internet]. 1989 Dec 1 [cited 2018 Nov 14];161(6 Pt 1):1464–9. Available from: https://www.sciencedirect.com/science/article/pii/0002937889909058

64. Sady MA, Haydon BB, Sady SP, Carpenter MW, Thompson PD, Coustan DR. Cardiovascular response to maximal cycle exercise duringpregnancy and at two and seven months postpartum. Am J Obstet Gynecol [Internet]. 1990 May 1 [cited 2018 Nov 14];162(5):1181–5. Available from: https://www.sciencedirect.com/science/article/pii/000293789090012V?via%3Dihub

65. Rafla NM, Beazely JM. The effect of maternal exercise on fetal umbilical artery waveforms. Eur J Obstet Gynecol Reprod Biol [Internet]. 1991 Jul 1 [cited 2018 Nov 14];40(2):119–22. Available from: https://www.sciencedirect.com/science/article/abs/pii/002822439190102Q

66. Bung P, Huch R, Huch A. Maternal and fetal heart rate patterns: a pregnant athlete during training and laboratory exercise tests; a case report [Internet]. Vol. 39, European Journal of Obstetrrcs & Gynecoiogv and Reproductive Biology. 1991 [cited 2018 Nov 27]. Available from: https://www.ejog.org/article/0028-2243(91)90143-9/pdf

67. Lotgering FK, Struijk PC, Doorn MBVAN, Spinnewijn WEM, Wallenburg HCS. Anaerobic threshold and respiratory compensation in pregnant women. J Appl Physiol [Internet]. 1995;85(6):1772–7. Available from: http://www.ncbi.nlm.nih.gov/pubmed/7649911

68. Artal R, Fortunato V, Welton A, Constantino N, Khodiguian N, Villalobos L, et al. A comparison of cardiopulmonary adaptations to exercise in pregnancy at sea level and altitude. Am J Obstet Gynecol [Internet]. 1995 Apr 1 [cited 2018 Nov 14];172(4):1170–80. Available from: https://www.sciencedirect.com/science/article/pii/0002937895914757?via%3Dihub

69. Manders MA., Sonder GJ., Mulder EJ., Visser GH. The effects of maternal exercise on fetal heart rate and movement patterns. Early Hum Dev [Internet]. 1997 May 28 [cited 2018 Nov 14];48(3):237–47. Available from: https://www.sciencedirect.com/science/article/pii/S0378378296018580?via%3Dihub

70. Kemp JG, Greer FA, Wolfe LA. Acid-base regulation after maximal exercise testing in late gestation. J Appl Physiol [Internet]. 1997 Aug [cited 2018 Nov 14];83(2):644–51. Available from: http://www.physiology.org/doi/10.1152/jappl.1997.83.2.644

71. Brenner IKM, Wolfe LA, Monga M, McGrath MJ. Physical conditioning effects on fetal heart rate responses to graded maternal exercise. Med Sci Sports Exerc [Internet]. 1999 Jun;31(6):792–9. Available from: https://journals.lww.com/acsm-msse/Fulltext/1999/06000/Physical_conditioning_effects_on_fetal_heart_rate.6.aspx

72. MacPhail A, Davies GA., Victory R, Wolfe LA. Maximal exercise testing in late gestation: fetal responses. Obstet Gynecol [Internet]. 2000 Oct 1 [cited 2018 Nov 13];96(4):565–70. Available from: http://www.ncbi.nlm.nih.gov/pubmed/11004360

73. Heenan AP, Wolfe LA, Davies GA. Maximal exercise testing in late gestation: maternal responses. Obstet Gynecol [Internet]. 2001 Jan 1 [cited 2018 Nov 13];97(1):127–34. Available from: https://www.sciencedirect.com/science/article/pii/S0029784400010899?via%3Dihub

74. Kennelly MM, Geary M, McCaffrey N, McLoughlin P, Staines A, McKenna P. Exercise-related changes in umbilical and uterine artery waveforms as assessed by Doppler ultrasound scans. Am J Obstet Gynecol. 2002 Sep;187(3):661–6.

75. Kennelly MM, McCaffrey N, McLoughlin P, Lyons S, McKenna P. Fetal heart rate response to strenuous maternal exercise: not a predictor of fetal distress. Am J Obstet Gynecol [Internet]. 2002 Sep 1 [cited 2018 Nov 13];187(3):811–6. Available from: https://www.sciencedirect.com/science/article/pii/S0002937802002077?via%3Dihub

76. Heenan AP, Wolfe LA, Davies GAL, McGrath MJ. Effects of human pregnancy on fluid regulation responses to short-term exercise. J Appl Physiol [Internet]. 2003 Dec [cited 2018 Nov 13];95(6):2321–7. Available from: http://www.physiology.org/doi/10.1152/japplphysiol.00984.2002

77. Heenan AP, Wolfe LA. Plasma osmolality and the strong ion difference predict respiratory adaptations in pregnant and nonpregnant women. Can J Physiol Pharmacol [Internet]. 2003 Sep [cited 2018 Nov 13];81(9):839–47. Available from: http://www.nrcresearchpress.com/doi/10.1139/y03-072

78. Wolfe LA, Heenan AP, Bonen A. Aerobic conditioning effects on substrate responses during graded cycling in pregnancy. Can J Physiol Pharmacol [Internet]. 2003 Jul [cited 2018 Nov 13];81(7):696–703. Available from: http://www.nrcresearchpress.com/doi/10.1139/y03-059

79. Lindqvist PG, Marsal K, Merlo J, Pirhonen JP. Thermal response to submaximal exercise before, during and after pregnancy: a longitudinal study. J Matern neonatal Med Off J Eur Assoc Perinat Med Fed Asia Ocean Perinat Soc Int Soc Perinat Obstet [Internet]. 2003 Mar 7 [cited 2018 Nov 13];13(3):152–6. Available from: http://www.tandfonline.com/doi/full/10.1080/jmf.13.3.152.156

80. Lynch A-MM, McDonald S, Magann EF, Evans SF, Choy PL, Dawson B, et al. Effectiveness and safety of a structured swimming program in previously sedentary women during pregnancy. J Matern Neonatal Med [Internet]. 2003 Jan 7 [cited 2018 Nov 13];14(3):163–9. Available from: http://www.ncbi.nlm.nih.gov/pubmed/14694971

81. Pirhonen JP, Lindqvist PG, Marsal K. A longitudinat study of maternal oxygen saturation during short-term submaximal exercise. Clin Physiol Funct Imaging [Internet]. 2003 Jan 1 [cited 2018 Nov 13];23(1):37–41. Available from: http://www.ncbi.nlm.nih.gov/pubmed/12558612

82. McAuley SE, Jensen D, McGrath MJ, Wolfe LA. Effects of human pregnancy and aerobic conditioning on alveolar gas exchange during exercise. Can J Physiol Pharmacol [Internet]. 2005 Jul [cited 2018 Nov 13];83(7):625–33. Available from: http://www.nrcresearchpress.com/doi/10.1139/y05-054

83. Weissgerber TL, Wolfe LA, Hopkins WG, Davies GALL. Serial respiratory adaptations and an alternate hypothesis of respiratory control in human pregnancy. Respir Physiol Neurobiol [Internet]. 2006 Aug 1 [cited 2018 Nov 13];153(1):39–53. Available from: https://www.sciencedirect.com/science/article/pii/S1569904805002521?via%3Dihub

84. Jensen D, Webb KA, Davies GAL, O’Donnell DE. Mechanical ventilatory constraints during incremental cycle exercise in human pregnancy: implications for respiratory sensation. J Physiol [Internet]. 2008 Oct 1 [cited 2018 Nov 13];586(19):4735–50. Available from: http://www.ncbi.nlm.nih.gov/pubmed/18687714

85. Kardel KR, Johansen BBBB, Voldner N, Iversen PO, Henriksen T. Association between aerobic fitness in late pregnancy and duration of labor in nulliparous women. ACTA Obstet Gynecol Scand [Internet]. 2009 Jan 1 [cited 2018 Nov 12];88(8):948–52. Available from: http://doi.wiley.com/10.1080/00016340903093583

86. Thorell E, Svärdsudd K, Andersson K, Kristiansson P. Moderate impact of full-term pregnancy on estimated peak oxygen uptake, physical activity and perceived health. Acta Obstet Gynecol Scand [Internet]. 2010 Sep 1 [cited 2018 Nov 12];89(9):1140–8. Available from: http://doi.wiley.com/10.3109/00016349.2010.487894

87. Rojas Vega S, Kleinert J, Sulprizio M, Hollmann W, Bloch W, Strüder HK. Responses of serum neurotrophic factors to exercise in pregnant and postpartum women. Psychoneuroendocrinology [Internet]. 2011 Feb 1 [cited 2018 Nov 12];36(2):220–7. Available from: https://www.sciencedirect.com/science/article/pii/S0306453010001812?via%3Dihub

88. Jędrzejko M, Nowosielski K, Poręba R, Ulman-Włodarz I, Bobiński R. Physical efficiency and activity energy expenditure in term pregnancy females measured during cardiopulmonary exercise tests with a supine cycle ergometer. J Matern Neonatal Med [Internet]. 2016 Dec 18 [cited 2018 Nov 10];29(23):3800–5. Available from: http://www.ncbi.nlm.nih.gov/pubmed/26866763

89. Nakagaki A, Inami T, Minoura T, Baba R, Iwase S, Sato M. Differences in autonomic neural activity during exercise between the second and third trimesters of pregnancy. J Obstet Gynaecol Res [Internet]. 2016 Aug 1 [cited 2018 Nov 10];42(8):951–9. Available from: http://www.ncbi.nlm.nih.gov/pubmed/27121772

90. Kardel KR. Effects of intense training during and after pregnancy in top-level athletes. Scand J Med Sci Sport [Internet]. 2005 Apr 1 [cited 2018 Nov 13];15(2):79–86. Available from: http://doi.wiley.com/10.1111/j.1600-0838.2004.00426.x

91. Ong MJ, Guelfi KJ, Hunter T, Wallman KE, Fournier PA, Newnham JP. Supervised home-based exercise may attenuate the decline of glucose tolerance in obese pregnant women. Diabetes Metab [Internet]. 2009 Nov 1 [cited 2018 Nov 12];35(5):418–21. Available from: https://www.sciencedirect.com/science/article/pii/S1262363609001426?via%3Dihub

92. Jensen D, Webb KA, O’Donnell DE. Chemical and mechanical adaptations of the respiratory system at rest and during exercise in human pregnancy. Appl Physiol Nutr Metab [Internet]. 2007 Dec [cited 2018 Nov 13];32(6):1239–50. Available from: http://www.nrcresearchpress.com/doi/10.1139/H07-120

93. Purdy GM, James MA, Wakefield PK, Skow RJ, Van Diepen S, May LE, et al. Maternal cardioautonomic responses during and following exercise throughout pregnancy. Appl Physiol Nutr Metab. 2019 Mar;44(3):263–70.

94. Matenchuk BA, James M, Skow RJ, Wakefield P, MacKay C, Steinback CD, et al. Longitudinal study of cerebral blood flow regulation during exercise in pregnancy. J Cereb BLOOD FLOW Metab. 2020 Nov;40(11):2278–88.

95. Correa M da S, Catai AM, Milan-Mattos JC, Porta A, Driusso P. Is pelvic floor muscle training able to alter the response of cardiovascular autonomic modulation and provide a possible cardiovascular benefit to pregnant women? Neurourol Urodyn. 2020 Nov;39(8):2272–83.

96. Bijl RC, Cornette JMJ, van Der Ham K, de Zwart ML, Dos Reis Miranda D, Steegers-Theunissen RPM, et al. The physiological effect of early pregnancy on a woman’s response to a submaximal cardiopulmonary exercise test. Physiol Rep. 2020 Nov;8(21).

97. Kennelly MM, Geary M, McCaffrey N, McLoughlin P, Staines A, McKenna P. Exercise-related changes in umbilical and uterine artery waveforms as assessed by Doppler ultrasound scans. Am J Obstet Gynecol [Internet]. 2002 Sep 1 [cited 2018 Nov 14];187(3):661–6. Available from: https://www.sciencedirect.com/science/article/pii/S0002937802001849?via%3Dihub

98. Pomerance JJ, Gluck L, Lynch VA. Physical fitness in pregnancy: its effect on pregnancy outcome. Am J Obstet Gynecol [Internet]. 1974 Aug 1 [cited 2018 Nov 14];119(7):867–76. Available from: https://www.sciencedirect.com/science/article/pii/0002937874900015?via%3Dihub

99. ONeill ME, O’Neill ME. Maternal rectal temperature and fetal heart rate responses to upright cycling in late pregnancy. Br J Sports Med [Internet]. 1996 Mar 1 [cited 2018 Nov 14];30(1):32–5. Available from: http://www.ncbi.nlm.nih.gov/pubmed/8665115

100. Lewis RD, Yates CY, Driskell JA. Riboflavin and thiamin status and birth outcome as a function of maternal aerobic exercise. Am J Clin Nutr [Internet]. 1988 Jul 1 [cited 2018 Nov 14];48(1):110–6. Available from: https://academic.oup.com/ajcn/article/48/1/110/4694812

101. Davenport MH, Charlesworth S, Vanderspank D, Sopper MM, Mottola MF. Development and validation of exercise target heart rate zones for overweight and obese pregnant women. Appl Physiol Nutr Metab [Internet]. 2008;33(5):984–9. Available from: http://www.nrcresearchpress.com/doi/abs/10.1139/H08-086

102. de Oliveria Melo AS, Silva JLP, Tavares JS, Barros VO, Leite DFBB, Amorim MMRR. Effect of a physical exercise program during pregnancy on uteroplacental and fetal blood flow and fetal growth: A randomized controlled trial. Obstet Gynecol [Internet]. 2012 Aug [cited 2018 Nov 12];120(2):302–10. Available from: http://insights.ovid.com/crossref?an=00006250-201208000-00017

103. Ruchat S-M, Davenport M, Giroux I, Hillier M, Batada A, Sopper M, et al. Walking Program of Low or Vigorous Intensity During Pregnancy Confers an Aerobic Benefit. Int J Sports Med [Internet]. 2012 Aug 17 [cited 2018 Nov 12];33(08):661–6. Available from: http://www.thieme-connect.de/DOI/DOI?10.1055/s-0032-1304635

104. Szymanski LM, Satin AJ. Strenuous exercise during pregnancy: Is there a limit? Am J Obstet Gynecol [Internet]. 2012;207(3):179.e1-179.e6. Available from: http://dx.doi.org/10.1016/j.ajog.2012.07.021

105. Bisson M, Rhéaume C, Bujold E, Tremblay A, Marc I. Modulation of blood pressure response to exercise by physical activity and relationshipwith resting blood pressure during pregnancy. J Hypertens [Internet]. 2014 Jul [cited 2018 Nov 10];32(7):1450–7. Available from: http://www.ncbi.nlm.nih.gov/pubmed/24721929

106. Santos CM dos, Santos WM dos, Gallarreta FMP, Pigatto C, Portela LOC, Morais EN de, et al. Effect of maternal exercises on biophysical fetal and maternal parameters: a transversal study. Einstein (São Paulo) [Internet]. 2016 Dec [cited 2018 Nov 26];14(4):455–60. Available from: http://www.scielo.br/scielo.php?script=sci_arttext&pid=S1679-45082016000400455&lng=en&tlng=en

107. Winn HN, HESS O, Goldstein I, Wackers F, HOBBINS JC. FETAL RESPONSES TO MATERNAL EXERCISE - EFFECT ON FETAL BREATHING AND BODY MOVEMENT. Am J Perinatol [Internet]. 1994 Jul 4 [cited 2018 Nov 14];11(4):263–6. Available from: http://www.thieme-connect.de/DOI/DOI?10.1055/s-2007-994588

108. SIBLEY L, RUHLING RO, Cameron-Foster J, CHRISTENSEN C, BOLEN T, CAMERONFOSTER J, et al. Swimming and physical fitness during pregnancy. J Nurse Midwifery [Internet]. 1981 Nov [cited 2018 Nov 14];26(6):3–12. Available from: http://linkinghub.elsevier.com/retrieve/pii/0091218281901695

109. Hesse CM, Tinius RA, Pitts BC, Olenick AA, Blankenship MM, Hoover DL, et al. Assessment of endpoint criteria and perceived barriers during maximal cardiorespiratory fitness testing among pregnant women. J Sports Med Phys Fitness. 2018;58(12):1844–51.

110. Mottola MF, Inglis S, Brun CR, Hammond J-A. Physiological and metabolic responses of late pregnant women to 40 min of steady-state exercise followed by an oral glucose tolerance perturbation. J Appl Physiol [Internet]. 2013 Sep 1 [cited 2018 Nov 10];115(5):597–604. Available from: http://www.ncbi.nlm.nih.gov/pubmed/23813524

111. Marquez-Sterling S, Perry AC, Kaplan TA, Halberstein RA, Signorile JF. Physical and psychological changes with vigorous exercise in sedentary primigravidae. Med Sci Sports Exerc [Internet]. 2000 Jan [cited 2018 Nov 13];32(1):58–62. Available from: http://www.msse.org

112. Santos IA, Stein R, Fuchs SC, Duncan BB, Ribeiro JP, Kroeff LR, et al. Aerobic Exercise and Submaximal Functional Capacity in Overweight Pregnant Women. Obstet Gynecol [Internet]. 2005 Aug [cited 2018 Nov 27];106(2):243–9. Available from: http://content.wkhealth.com/linkback/openurl?sid=WKPTLP:landingpage&an=00006250-200508000-00007

113. Salvesen K a. Å, Hem E, Sundgot-Borgen J. Fetal wellbeing may be compromised during strenuous exercise among pregnant elite athletes. Br J Sports Med [Internet]. 2012 Mar [cited 2018 Nov 12];46(4):279–83. Available from: http://www.ncbi.nlm.nih.gov/pubmed/21393257

114. Marshall MR, Pivarnik JM. Perceived Exertion of Physical Activity During Pregnancy. J Phys Act Heal [Internet]. 2015 Jul [cited 2018 Nov 10];12(7):1039–43. Available from: http://www.ncbi.nlm.nih.gov/pubmed/26462301

115. Da Silva EG, De Godoy I, De Oliveira Antunes LC, Da Silva EG, Peraçoli JC. Respiratory parameters and exercise functional capacity in preeclampsia. Hypertens Pregnancy. 2010;29(3):301–9.

116. Ramírez-Vélez R, Aguilar de Plata AC, Escudero MM, Echeverry I, Ortega JG, Salazar B, et al. Influence of regular aerobic exercise on endothelium-dependent vasodilation and cardiorespiratory fitness in pregnant women. J Obstet Gynaecol Res [Internet]. 2011 Nov [cited 2018 Nov 12];37(11):1601–8. Available from: http://www.ncbi.nlm.nih.gov/pubmed/21733037

117. Radzikowska E, Wiatr E, Franczuk M, Bestry I, Roszkowski-Śliż K. Lung Function in Pregnancy in Langerhans Cell Histiocytosis. In: Advances in experimental medicine and biology [Internet]. 2017 [cited 2018 Nov 10]. p. 73–83. Available from: http://www.ncbi.nlm.nih.gov/pubmed/28744781

118. Angel Oviedo-Caro M, Bueno-Antequera J, Munguia-Izquierdo D. Associations of 24-hours activity composition with adiposity and cardiorespiratory fitness: The PregnActive project. Scand J Med Sci Sports. 2020 Feb;30(2):295–302.

119. Dennis AT, Salman M, Paxton E, Flint M, Leeton L, Roodt F, et al. Resting Hemodynamics and Response to Exercise Using the 6-Minute Walk Test in Late Pregnancy: An International Prospective Multicentre Study. Anesth Analg. 2019;129(2):450–7.

120. Amola M, Pawara S, Kalra S. Effect of Inspiratory Muscle Training and Diaphragmatic Breathing Exercises on Dyspnea, Pulmonary Functions, Fatigue and Functional Capacity in Pregnancy during Third Trimester. J Clin DIAGNOSTIC Res. 2019;13(8):YC1–4.

121. Dibblee L, Graham TEE. A longitudinal study of changes in aerobic fitness, body composition, and energy intake in primigravid patients. Am J Obstet Gynecol [Internet]. 1983 Dec 15 [cited 2018 Nov 14];147(8):908–14. Available from: https://www.sciencedirect.com/science/article/pii/0002937883902442?via%3Dihub

122. Williams A, Reilly T, Campbell I, Sutherst J. Investigation of changes in responses to exercise and in mood during pregnancy. Ergonomics [Internet]. 1988 Nov [cited 2018 Nov 14];31(11):1539–49. Available from: http://www.tandfonline.com/doi/abs/10.1080/00140138808966803

123. Petrov Fieril K, Glantz A, Fagevik Olsen M. The efficacy of moderate-to-vigorous resistance exercise during pregnancy: A randomized controlled trial. Acta Obstet Gynecol Scand. 2015;94(1):35–42.

124. Hjorth MF, Kloster S, Girma T, Faurholt-Jepsen D, Andersen G, Kæstel P, et al. Level and intensity of objectively assessed physical activity among pregnant women from urban Ethiopia. BMC Pregnancy Childbirth [Internet]. 2012 Dec 17 [cited 2018 Nov 11];12(1):154. Available from: http://www.ncbi.nlm.nih.gov/pubmed/23244057

125. Atay E, Başalan İz F. Investigation of the effect of changes in muscle strength in gestational age upon fear of falling and quality of life. Turkish J Med Sci [Internet]. 2015 [cited 2018 Nov 26];45(4):977–83. Available from: http://journals.tubitak.gov.tr/medical/

126. Kalliokoski P, Rodhe N, Bergqvist Y, Lofvander M, Löfvander M. Long-term adherence and effects on grip strength and upper leg performance of prescribed supplemental vitamin D in pregnant and recently pregnant women of Somali and Swedish birth with 25- hydroxyvitamin D deficiency: a before-and-after treatment study. BMC Pregnancy Childbirth [Internet]. 2016 Nov 15 [cited 2018 Nov 10];16(1):353. Available from: http://bmcpregnancychildbirth.biomedcentral.com/articles/10.1186/s12884-016-1117-3

127. Ngaka TCC, Coetzee JFF, Dyer RAA. The Influence of Body Mass Index on Sensorimotor Block and Vasopressor Requirement During Spinal Anesthesia for Elective Cesarean Delivery. Anesth Analg [Internet]. 2016 Dec [cited 2018 Nov 10];123(6):1527–34. Available from: http://www.ncbi.nlm.nih.gov/pubmed/27870737

128. Rodriguez-Diaz L, Ruiz-Frutos C, Maria Vazquez-Lara J, Ramirez-Rodrigo J, Villaverde-Gutierrez C, Torres-Luque G. Effectiveness of a physical activity programme based on the Pilates method in pregnancy and labour. Enferm Clin. 2017;27(5):271–7.

129. Baker PN, Johnson IR. The use of the hand-grip test for predicting pregnancy-induced hypertension. Eur J Obstet Gynecol Reprod Biol [Internet]. 1994 Sep 1 [cited 2018 Nov 27];56(3):169–72. Available from: https://www.sciencedirect.com/science/article/abs/pii/0028224394901651?via%3Dihub

130. Feiner B, Weksler R, Ohel G, Degani S. The influence of maternal exercise on placental blood flow measured by Simultaneous Multigate Spectral Doppler Imaging (SM- SDI). Ultrasound Obstet Gynecol [Internet]. 2000 Jun 1 [cited 2018 Nov 13];15(6):498–501. Available from: http://doi.wiley.com/10.1046/j.1469-0705.2000.00146.x

131. Rogers MS, Tomlinson B. Change in cardiovascular indices with position and isometric exercise throughout pregnancy: Assessment by impedance cardiography and oscillometric sphygmomanometry. Hypertens Pregnancy [Internet]. 1998 Jan 7 [cited 2018 Nov 27];17(2):191–202. Available from: http://www.tandfonline.com/doi/full/10.3109/10641959809006075

132. Takeda K, Yoshikata H, Imura M. Do Squat Exercises With Weight Shift During Pregnancy Improve Postural Control? Int J WOMENS Heal Reprod Sci. 2019;7(1):10–6.

133. Gutke A, Östgaard HC, Öberg B. Predicting Persistent Pregnancy-Related Low Back Pain. Spine (Phila Pa 1976) [Internet]. 2008 May [cited 2018 Nov 13];33(12):E386–93. Available from: https://insights.ovid.com/crossref?an=00007632-200805200-00021

134. O’Connor PJ, Poudevigne MS, Cress ME, Motl RW, Clapp JF. Safety and Efficacy of Supervised Strength Training Adopted in Pregnancy. J Phys Act Heal [Internet]. 2011 Mar 1 [cited 2018 Nov 27];8(3):309–20. Available from: http://journals.humankinetics.com/doi/10.1123/jpah.8.3.309

135. Garshasbi A, Faghih Zadeh S. The effect of exercise on the intensity of low back pain in pregnant women. Int J Gynecol Obstet. 2005;88(3):271–5.

136. Gilleard W, Crosbie J, Smith R. Effect of pregnancy on trunk range of motion when sitting and standing. Acta Obstet Gynecol Scand [Internet]. 2002 Nov 1 [cited 2018 Nov 27];81(11):1011–20. Available from: http://www.blackwell-synergy.com/links/doi/10.1034%2Fj.1600-0412.2002.811104.x

137. Cherni Y, Desseauve D, Decatoire A, Veit-Rubinc N, Begon M, Pierre F, et al. Evaluation of ligament laxity during pregnancy. J Gynecol Obstet Hum Reprod. 2019 May;48(5):351–7.

138. Rodríguez-Díaz L, Ruiz-Frutos C, Vázquez-Lara JM, Ramírez-Rodrigo J, Villaverde-Gutiérrez C, Torres-Luque G. Efectividad de un programa de actividad física mediante el método Pilates en el embarazo y en el proceso del parto. Enfermería Clínica [Internet]. 2017 Sep 1 [cited 2018 Nov 26];27(5):271–7. Available from: https://www.sciencedirect.com/science/article/pii/S1130862117300815?via%3Dihub

139. Marnach ML, Ramin KD, Ramsey PS, Song S-W, Stensland JJ, An K-N. Characterization of the relationship between joint laxity and maternal hormones in pregnancy. Obstet Gynecol [Internet]. 2003 Feb [cited 2018 Nov 13];101(2):331–5. Available from: http://www.ncbi.nlm.nih.gov/pubmed/12576258

140. Butler EE, Colon I, Druzin ML, Rose J, Colón I, Druzin ML, et al. Postural equilibrium during pregnancy: Decreased stability with an increased reliance on visual cues. Am J Obstet Gynecol [Internet]. 2006 Oct 1 [cited 2018 Nov 13];195(4):1104–8. Available from: https://www.sciencedirect.com/science/article/pii/S0002937806007393?via%3Dihub

141. Ribas I S, Guirro ECO. Analysis of plantar pressure and postural balance during different phases of pregnancy. BRAZILIAN J Phys Ther. 2007;11(5):391–6.

142. Oliveira LF, Vieira TMM, Macedo AR, Simpson DM, Nadal J. Postural sway changes during pregnancy: A descriptive study using stabilometry. Eur J Obstet Gynecol Reprod Biol [Internet]. 2009 Nov 1 [cited 2018 Nov 12];147(1):25–8. Available from: https://www.sciencedirect.com/science/article/pii/S0301211509004333?via%3Dihub

143. Karadag-Saygi E, Unlu-Ozkan F, Basgul A. Plantar Pressure and Foot Pain in the Last Trimester of Pregnancy. Foot Ankle Int [Internet]. 2010 Feb 1 [cited 2018 Nov 12];31(2):153–7. Available from: http://journals.sagepub.com/doi/10.3113/FAI.2010.0153

144. Yu Y, Chung HC, Hemingway L, Stoffregen TA. Standing body sway in women with and without morning sickness in pregnancy. Gait Posture [Internet]. 2013 Jan 1 [cited 2018 Nov 12];37(1):103–7. Available from: https://www.sciencedirect.com/science/article/pii/S096663621200238X?via%3Dihub

145. Opala-Berdzik A, Błaszczyk JW, Bacik B, Cieślińska-Świder J, Świder D, Sobota G, et al. Static Postural Stability in Women during and after Pregnancy: A Prospective Longitudinal Study. McCrory JL, editor. PLoS One [Internet]. 2015 Jun 8 [cited 2018 Nov 10];10(6):e0124207. Available from: http://www.ncbi.nlm.nih.gov/pubmed/26053046

146. Shibayama Y, Kuwata T, Yamaguchi J, Matsumoto M, Watanabe M, Nakano R, et al. Changes in standing body sway of pregnant women after long-term bed rest. J Obstet Gynaecol (Lahore) [Internet]. 2016 May 18 [cited 2018 Nov 10];36(4):479–82. Available from: http://www.ncbi.nlm.nih.gov/pubmed/26471310

147. Moreira LS, Elias LA, Gomide AB, Vieira MF, Do Amaral WN. A longitudinal assessment of myoelectric activity, postural sway, and low-back pain during pregnancy. Acta Bioeng Biomech [Internet]. 2017 [cited 2018 Nov 24];Vol. 19(nr 3). Available from: http://yadda.icm.edu.pl/yadda/element/bwmeta1.element.baztech-b0cc7f75-aa2d-4716-8aaf-d037c8b9d580;jsessionid=3A62F7482C92C62BEB4A73EC922F11CD

148. Opala-Berdzik A, Błaszczyk JW, Świder D, Cieślińska-Świder J. Trunk forward flexion mobility in reference to postural sway in women after delivery: A prospective longitudinal comparison between early pregnancy and 2- and 6-month postpartum follow-ups. Clin Biomech [Internet]. 2018 Jul 1 [cited 2018 Nov 26];56:70–4. Available from: https://www.sciencedirect.com/science/article/pii/S0268003318300445?via%3Dihub

149. Catena RD, Campbell N, Wolcott WC, Rothwell SA. Anthropometry, standing posture, and body center of mass changes up to 28 weeks postpartum in Caucasians in the United States. Gait Posture. 2019 May;70:196–202.

150. Fontana Carvalho AP, Dufresne SS, de Oliveira M, Couto Furlanetto K, Dubois M, Dallaire M, et al. Effects of lumbar stabilization and muscular stretching on pain, disabilities, postural control and muscle activation in pregnant woman with low back pain. Eur J Phys Rehabil Med. 2020 Jun;56(3):297–306.

151. Ersal, T., McCrory, J. L., & Sienko KH, Ersal T, McCrory JL, Sienko KH. Theoretical and experimental indicators of falls during pregnancy as assessed by postural perturbations. Gait Posture [Internet]. 2014 Jan [cited 2018 Nov 10];39(1):218–23. Available from: http://www.ncbi.nlm.nih.gov/pubmed/23953273

152. Opala-Berdzik A, Bacik B, Markiewicz A, Cieślińska-Świder J, Swider D, Sobota G, et al. Comparison of static postural stability in exercising and non-exercising women during the perinatal period. Med Sci Monit [Internet]. 2014 Oct 8 [cited 2018 Nov 26];20:1865–70. Available from: http://www.ncbi.nlm.nih.gov/pubmed/25293983

153. Valerio PM, Goncalves VE, Zordao CC, Rezende MS, Dantas Moises EC, de Olveira Guirro EC. Influence of type 1 diabetes on the postural control of women in the third gestational trimester. Clin Biomech. 2020 Jul;77.

154. Takeda K, Yoshikata H, Imura M. Changes in Posture Control of Women That Fall During Pregnancy. 2018 [cited 2018 Nov 26]; Available from: http://www.ijwhr.net

155. Davies J, Fernando R, McLeod A, Verma S, Found P. Postural stability following ambulatory regional analgesia for labor. Anesthesiology [Internet]. 2002 Dec [cited 2018 Nov 13];97(6):1576–81. Available from: http://www.ncbi.nlm.nih.gov/pubmed/12459687

156. Branco M, Santos-Rocha R, Aguiar L, Vieira F, Veloso A. Kinematic analysis of gait in the second and third trimesters of pregnancy. J Pregnancy [Internet]. 2013 [cited 2018 Nov 12];2013:718095. Available from: http://www.ncbi.nlm.nih.gov/pubmed/23431450

157. Cakmak B, Inanir A, Nacar MC, Filiz B. The Effect of Maternity Support Belts on Postural Balance in Pregnancy. PM&R [Internet]. 2014 Jul 1 [cited 2018 Nov 10];6(7):624–8. Available from: http://www.ncbi.nlm.nih.gov/pubmed/24412671

158. Inanir A, Cakmak B, Hisim Y, Demirturk F. Evaluation of postural equilibrium and fall risk during pregnancy. Gait Posture [Internet]. 2014 Apr 1 [cited 2018 Nov 10];39(4):1122–5. Available from: http://www.ncbi.nlm.nih.gov/pubmed/24630464

159. McCrory J, Chambers A, Daftary A, Redfern M. Dynamic postural stability in pregnant fallers and non-fallers. BJOG An Int J Obstet Gynaecol [Internet]. 2010 Jul 1 [cited 2018 Nov 12];117(8):954–62. Available from: http://doi.wiley.com/10.1111/j.1471-0528.2010.02589.x

160. Wu W, Meijer OG, Lamoth CJCC, Uegaki K, Van Dieën JH, Wuisman PIJMJM, et al. Gait coordination in pregnancy: Transverse pelvic and thoracic rotations and their relative phase. Clin Biomech [Internet]. 2004 Jun 1 [cited 2018 Nov 13];19(5):480–8. Available from: https://www.sciencedirect.com/science/article/pii/S0268003304000427?via%3Dihub

161. Forczek W, Staszkiewicz R. Changes of kinematic gait parameters due to pregnancy. Acta Bioeng Biomech [Internet]. 2012 [cited 2018 Nov 27];14(4):113–9. Available from: http://www.ncbi.nlm.nih.gov/pubmed/23394129

162. Gottschall JS, Sheehan RC, Downs DS. Pregnant women exaggerate cautious gait patterns during the transition between level and hill surfaces. J Electromyogr Kinesiol Off J Int Soc Electrophysiol Kinesiol [Internet]. 2013 Oct [cited 2018 Nov 10];23(5):1237–42. Available from: http://www.ncbi.nlm.nih.gov/pubmed/23770002

163. McCrory JL, Chambers AJ, Daftary A, Redfern MS. The pregnant “waddle”: An evaluation of torso kinematics in pregnancy. J Biomech [Internet]. 2014 Sep 22 [cited 2018 Nov 10];47(12):2964–8. Available from: http://www.ncbi.nlm.nih.gov/pubmed/25108664

164. Krkeljas Z. Changes in gait and posture as factors of dynamic stability during walking in pregnancy. Hum Mov Sci [Internet]. 2018 Apr 1 [cited 2018 Nov 10];58:315–20. Available from: https://www.sciencedirect.com/science/article/pii/S0167945717307509?via%3Dihub

165. Catena RD, Campbell N, Werner AL, Iverson KM. Anthropometric Changes During Pregnancy Provide Little Explanation of Dynamic Balance Changes. J Appl Biomech. 2019 Jun;35(3):232–9.

166. Forczek W, Ivanenko Y, Curyło M, Frączek B, Masłoń A, Salamaga M, et al. Progressive changes in walking kinematics throughout pregnancy-A follow up study. Gait Posture. 2019 Feb;68:518–24.

167. Forczek W, Masłoń A, Frączek B, Curyło M, Salamaga M, Suder A. Does the first trimester of pregnancy induce alterations in the walking pattern? PLoS One. 2019;14(1):e0209766.

168. Forczek W, Ivanenko Y, Salamaga M, Sylos-Labini F, Fraczek B, Maslon A, et al. Pelvic movements during walking throughout gestation - the relationship between morphology and kinematic parameters. Clin Biomech. 2020 Jan 1;71:146–51.

169. Catena RD, Bailey JP, Campbell N, Stewart BC, Marion SJ. Correlations between joint kinematics and dynamic balance control during gait in pregnancy. Gait Posture. 2020 Jul;80:106–12.

170. Gimunova M, Zvonar M, Sebera M, Turcinek P, Kolarova K. Special footwear designed for pregnant women and its effect on kinematic gait parameters during pregnancy and postpartum period. PLoS One. 2020 May;15(5).

171. McCrory JL, Chambers AJ, Daftary A, Redfern MS. Torso kinematics during gait and trunk anthropometry in pregnant fallers and non-fallers. Gait Posture. 2020 Feb;76:204– 9.

172. Rothwell SA, Eckland CB, Campbell N, Connolly CP, Catena RD. An analysis of postpartum walking balance and the correlations to anthropometry. Gait Posture. 2020 Feb;76:270–6.

173. Takeda K. A Kinesiological Analysis of the Stand-to-Sit during the Third Trimester. J Phys Ther Sci [Internet]. 2012 [cited 2018 Nov 27];24(7):621–4. Available from: http://jlc.jst.go.jp/DN/JST.JSTAGE/jpts/24.621?lang=en&from=CrossRef&type=abstract

174. Catena RD, Bailey JP, Campbell N, Music HE. Stand-to-sit kinematic changes during pregnancy correspond with reduced sagittal plane hip motion. Clin Biomech. 2019 Jul;67:107–14.

175. Sawa R, Doi T, Asai T, Watanabe K, Taniguchi T, Ono R. Differences in trunk control between early and late pregnancy during gait. Gait Posture [Internet]. 2015 Oct [cited 2018 Nov 10];42(4):455–9. Available from: https://linkinghub.elsevier.com/retrieve/pii/S0966636215007870

176. Błaszczyk JW, Opala-Berdzik A, Plewa M. Adaptive changes in spatiotemporal gait characteristics in women during pregnancy. Gait Posture [Internet]. 2016 Jan [cited 2018 Nov 10];43:160–4. Available from: http://www.ncbi.nlm.nih.gov/pubmed/26480840

177. Martinez-Marti F, Martinez-Garcia MS, Carvajal MA, Palma AJ, Anguiano M, Lallena AM. Fractal behavior of the trajectories of the foot centers of pressure during pregnancy. Biomed Phys Eng EXPRESS. 2019 Feb;5(2).

178. Sady SP, Carpenter MW, Sady MA, Haydon B, Hoegsberg B, Cullinane EM, et al. Prediction of VO2max during cycle exercise in pregnant women. J Appl Physiol [Internet]. 1988 Aug [cited 2018 Nov 14];65(2):657–61. Available from: http://www.ncbi.nlm.nih.gov/pubmed/3170418

179. Gutke A, Kjellby-Wendt G, Oberg B. The inter-rater reliability of a standardised classification system for pregnancy-related lumbopelvic pain. Man Ther. 2010 Feb;15(1):13– 8.

180. García-Hermoso A, Cavero-Redondo I, Ramírez-Vélez R, Ruiz JR, Ortega FB, Lee D-CC, et al. Muscular Strength as a Predictor of All-Cause Mortality in an Apparently Healthy Population: A Systematic Review and Meta-Analysis of Data From Approximately 2 Million Men and Women [Internet]. Archives of Physical Medicine and Rehabilitation W.B. Saunders; Oct 1, 2018 p. 2100–2113.e5. Available from: https://www.sciencedirect.com/science/article/pii/S0003999318300790?via%3Dihub

181. Opala-Berdzik A, Bacik B, Markiewicz A, Cieślińska-Świder J, Swider D, Sobota G, et al. Comparison of Static Postural Stability in Exercising and Non-Exercising Women During the Perinatal Period. Med Sci Monit [Internet]. 2014 Oct 8 [cited 2018 Nov 26];20:1865–70. Available from: http://www.ncbi.nlm.nih.gov/pubmed/25293983

182. Opala-Berdzik A, Blaszczyk JW, Swider D, Cieslinska-Swider J. Trunk forward flexion mobility in reference to postural sway in women after delivery: A prospective longitudinal comparison between early pregnancy and 2-and 6-month postpartum follow-ups. Clin Biomech. 2018 Jul;56:70–4.

183. Cakmak B, Ribeiro AP, Inanir A. Postural balance and the risk of falling during pregnancy. J Matern Neonatal Med [Internet]. 2016 May [cited 2018 Nov 10];29(10):1623–5. Available from: http://www.ncbi.nlm.nih.gov/pubmed/26212584

184. Ozturk G, Kulcu DG, Aydog E, Kaspar C, Ugurel B. Effects of lower back pain on postural equilibrium and fall risk during the third trimester of pregnancy. J Matern NEONATAL Med. 2016;29(8):1358–62.

185. McCrory JL, Chambers AJ, Daftary A, Redfern MS. Ground reaction forces during gait in pregnant fallers and non-fallers. Gait Posture [Internet]. 2011 Oct 1 [cited 2018 Nov 12];34(4):524–8. Available from: https://www.sciencedirect.com/science/article/pii/S096663621100230X?via%3Dihub

186. Birsner ML, Gyamfi-Bannerman C. Physical Activity and Exercise During Pregnancy and the Postpartum Period ACOG Committee Opinion Summary, Number 804. Obstet Gynecol. 2020;135(4):E178–88.

