## Supplementary material for "Assessing physical fitness during pregnancy: validity and reliability of fitness tests, and relationship with maternal and neonatal health-related outcomes. A systematic review": ESM2_TableS1_TableS2

**Electronic Supplementary Material 2.** Search strategy in **PubMed** and **Web of Science.**

For PubMed, we used Medical Subject Heading (MeSH) terms. This is a powerful method to enhance the quality of the search. In addition, all MeSH terms were included without the command MeSH attached, to consolidate our results and avoid losing those papers not included in MeSH database. This is because some MeSH terms were introduced in a specific date (e.g., ‘physical fitness’ was included in 1996). Hence papers published in a previous date would be lost. The same process was developed with terms not available in the MeSH database such as agility, aerobic capacity, etc. (see ESM 2-Table-S1) for search criteria and related terms.

All terms were combined using the connector OR for similar criteria. The connector ‘AND’ was used to combine population group (i.e., pregnant women), to delimit date of publication ("0001/01/01"[PDat]: "2021/01/15"[PDat]), to include full text papers, and to include studies performed in humans. A similar search strategy and terms combination was undertaken in WoS (ESM 2-Table-S2), although MeSH terms and its appropriate terms connection were not used as they are exclusive for PubMed.

The first step of the search was to look for systematic reviews and meta-analysis within the field of this systematic review. Since there was no such article published regarding our topic, the research team agreed on starting the search with no limit on the publication date. Then, an initial search was undertaken in both databases following the strategy explained in ESM 2-Table-S1 and ESM 2-Table-S2 for PubMed and WoS database respectively. The results from both, were merged.

**Electronic Supplementary Material 2, Table S1.** Search strategy used and number of articles found in **Pubmed**.

| ***Search Strategy*** | | | | | |
| --- | --- | --- | --- | --- | --- |
| ("Pregnant Women"[Mesh] OR “Pregnant Women” OR "Pregnancy"[Mesh] OR “Pregnancy”) AND ("Physical Fitness"[Mesh] OR "Physical Fitness" OR “Physical Conditioning” OR "Exercise Test"[Mesh] OR "Exercise Test" OR "Fitness Trackers"[Mesh] OR “Fitness Trackers" OR “Muscle Strength”[MeSH] or “Muscle Strength” OR “Muscular fitness” OR “Range of motion, articular”[Mesh] OR “Range of motion, articular” OR “Postural Balance”[MeSH] OR “Postural Balance” OR “Walk Test”[Mesh] OR “Walk Test” OR “Cardiorespiratory Fitness” [Mesh] OR “Cardiorespiratory Fitness” OR “Agility” OR “running speed” OR “aerobic fitness” OR “aerobic capacity” OR “maximal oxygen consumption” OR “V02max” OR “Physical function”) AND full text[sb] AND ( "0001/01/01"[PDat] : "2021/01/15"[PDat] ) AND Humans[Mesh] | | | | | |
| **Search criteria 1** | | **MeSH**  **Entry Terms for**  **Criteria 1** |  | **Search criteria 2** | **MeSH**  **Entry Terms for**  **Criteria 2** |
| Pregnant Women (MeSH)  Pregnancy (MeSH) | | Women, Pregnant  Pregnant Woman  Woman, Pregnant |  | Physical fitness (MeSH) | Fitness, Physical |
|  |  |  |  | Exercise Test (MeSH) | Exercise Tests  Test, Exercise  Tests, Exercise  Arm Ergometry Test  Arm Ergometry Tests  Ergometry Test, Arm  Ergometry Tests, Arm  Test, Arm Ergometry  Tests, Arm Ergometry  Bicycle Ergometry Test  Bicycle Ergometry Tests  Ergometry Test, Bicycle  Ergometry Tests, Bicycle  Test, Bicycle Ergometry  Tests, Bicycle Ergometry  Fitness Testing  Fitness Testings  Testing, Fitness  Testings, Fitness  Step Test  Step Tests  Test, Step  Tests, Step  Stress Test  Stress Tests  Test, Stress  Tests, Stress  Treadmill Test  Test, Treadmill  Tests, Treadmill  Treadmill Tests  Physical Fitness Testing  Fitness Testing, Physical  Fitness Testings, Physical  Physical Fitness Testings  Testing, Physical Fitness  Testings, Physical Fitness  Cardiopulmonary Exercise Test  Cardiopulmonary Exercise Tests  Exercise Test, Cardiopulmonary  Exercise Tests, Cardiopulmonary  Test, Cardiopulmonary Exercise |
|  |  |  |  | Fitness Trackers (MeSH) | Fitness Tracker  Tracker, Fitness  Trackers, Fitness  Physical Fitness Trackers  Fitness Tracker, Physical  Fitness Trackers, Physical  Physical Fitness Tracker  Tracker, Physical Fitness  Trackers, Physical Fitness  Activity Trackers  Activity Tracker  Tracker, Activity  Trackers, Activity  Personal Fitness Trackers  Fitness Tracker, Personal  Fitness Trackers, Personal  Personal Fitness Tracker  Tracker, Personal Fitness  Trackers, Personal Fitness |
|  |  |  |  | Muscle Strength (MeSH) | Strength, Muscle |
|  |  |  |  | Muscle strength dynamometer (MeSH) | Dynamometer, Muscle Strength  Dynamometers, Muscle Strength  Muscle Strength Dynamometers |
|  |  |  |  | Range of motion, articular (MeSH) | Joint Range of Motion  Joint Flexibility  Flexibility, Joint  Range of Motion  Passive Range of Motion |
|  |  |  |  | Postural Balance (MeSH) | Musculoskeletal Equilibrium  Equilibrium, Musculoskeletal  Postural Equilibrium  Equilibrium, Postural  Balance, Postural |
|  |  |  |  | Walk Test  (MeSH) | Test, Walk  Tests, Walk  Walk Tests  6-Minute Walk Test  6 Minute Walk Test  6-Minute Walk Tests  Test, 6-Minute Walk  Tests, 6-Minute Walk  Walk Test, 6-Minute  Walk Tests, 6-Minute  Incremental Shuttle Walk Test  Endurance Shuttle Walk Test |
|  |  |  |  | Cardiorespiratory fitness (MeSH) | Fitness, Cardiorespiratory |
| **Total items found** | **Without filters:** 1657 | | | | |
|  | **With Humans filter:** 1135 | | | | |
|  | **With Full Text filter:** 1388 | | | | |
|  | **With Humans & Full Text Filter:** 930 | | | | |

The search recruited articles published until 15.01.21: no starting date limit was set for the search.

MeSH (Medical Subject Headings) is the National Library of Medicine controlled vocabulary thesaurus used for indexing articles for PubMed

**Electronic Supplementary Material 2, Table S2.** Search strategy used and number of articles found in **Web of Science**

| ***Search Strategy*** | |
| --- | --- |
| TS=("Pregnant women" OR "pregnancy" OR "pregnan*") AND (("Physical Conditioning" OR "Physical fitness" OR "Exercise Test*" OR "Arm Ergometry Test*" OR "Bicycle Ergometry Test*" OR "Step Test*" OR "Treadmill Test*" OR "Physical Fitness Test*" OR "Cardiopulmonary Exercise Test*" OR "Fitness Tracker*" OR "Physical Fitness Tracker*" OR "Activity Tracker*" OR "Personal Fitness Tracker*") OR ("Muscle Strength" OR "Muscular Fitness" OR "Muscle strength dynamometer*") OR ("Joint Range of motion" OR "Joint flexibility" OR "Flexibility" OR "Range of motion" OR "Passive Range of Motion") OR ("Postural Balance" OR "Musculoskeletal Equilibrium" OR "Equilibrium" OR "Postural Equilibrium") OR ("Walk Test*" OR "6-Minute Walk Test*" OR "Incremental Shuttle Walk Test*" OR "Endurance Shuttle Walk Test") OR ("Cardiorrespiratory Fitness" OR "Cardiovascular Fitness OR “Aerobic Fitness” OR “Aerobic Capacity” OR “Maximal Oxygen Consumption” OR “V02max") OR (“Agility” OR “running speed” OR “aerobic fitness” )) | |
| **Total items found** | 1687 |

The search recruited articles published until 15.01.21 no starting date limit was set for the search.
