## Supplementary material for "Assessing physical fitness during pregnancy: validity and reliability of fitness tests, and relationship with maternal and neonatal health-related outcomes. A systematic review": ESM3_Description_quality_score

**Electronic Supplementary Material 3. Comprehensive description of the three quality assessment scores used in the present systematic review.**

The first quality score [24], was used to evaluate the quality of the articles that assessed validity. This list included three items based on sample size, description of the article population and statistical analysis to assess validity of each article. The validity quality score ranged from 0 to 6 (Table-S3). A score of 0-2 defined a very low-quality article; a score of 3-4 defined a low-quality article; and a score of 5-6 defined a high-quality article.

The second quality score [25] was employed to rate the studies that measured reliability (ESM 4 – Table-S4). This ranking was formed by four items based on description of the participants, the time interval, the results and appropriateness of statistical analyses. Each item in both, was rated from 0 (the lowest quality) to 2 (the highest quality). The reliability quality score ranged from 0 to 8 (ESM 4– Table-S4). A score of 0-1 defined a very low-quality article; a score of 2-5 defined a low-quality article; and a score of 6-8 defined a high-quality article.

The third quality score (ESM 4– Table-S5) was created to evaluate those studies that assessed association of PF with health-related outcomes. We adapted a score previously used in the Effective Public Health Practice Project (EPHPP) [26] which has been used in similar reviews [27]. The health-related outcomes quality score ranged from 0 to 5 (ESM 4– Table S5). A score of 0-2 defined a very low-quality article, a score of 3-4 defined a low-quality article, and a score of 5 defined a high-quality score. Three quality scores were calculated by counting the number of positive items.
