## Supplementary material for "Assessing physical fitness during pregnancy: validity and reliability of fitness tests, and relationship with maternal and neonatal health-related outcomes. A systematic review": ESM4_TableS3_S4_S5

**Electronic Supplementary Material 4 Table S3. Quality assessment criteria to evaluate validity and reliability studies.**

| **Grading system parameter** | **Grade** | **Criterion** |
| --- | --- | --- |
| Number of study subjects | 0 | n < 10 |
|  | 1 | n= 11-50 |
|  | 2 | n>51 |
| Description of the study population regarding to age, sex, health status, fitness levels, etc | 0 | Less items than required for grade 1 |
|  | 1 | At least age and week of gestation. |
|  | 2 | Age, week of gestation, health status and fitness levels and more. |
| Statistical analysis included in the study | 0 | Those not included in grade 1 |
|  | 1 | Error indexes or regression analysis |
|  | 2 | ≥3 items of Bland-Altamn plot and or ANOVA for repeated measurements |
| **Electronic Supplementary Material 4 Table S4. Quality assessment criteria to evaluate reliability studies.** | | |
| **Grading system parameter** | **Grade** | **Criterion** |
| Description of the participants | 0 | Less items than required for grade 1. |
|  | 1 | At least age and week of gestation. |
|  | 2 | Age, week of gestation, health status and fitness levels and more. |
| Description of the time interval | 0 | Interval unknown. |
|  | 1 | Vague and imprecise information about interval. |
|  | 2 | Precise and complete description about interval. |
| Description of the results | 0 | Less results presented than required for grade. |
|  | 1 | Description of test-retest results or description of the differences. |
|  | 2 | Description of test-retest results and description of the differences. |
| Appropriateness of statistic | 0 | Only coefficient of variation |
|  | 1 | Everything between grades 0 and 2 (normally – but not always – correlation plus an additional statistic). |
|  | 2 | At least paired statistics, ANOVA for repeated measures (or non-parametrical corresponding tests) or Bland- Altman method. |

**Electronic Supplementary Material 4 Table S5. Quality assessment criteria to evaluate health-related outcomes studies.**

| **Grading system parameter** | **Grade** | **Criterion** |
| --- | --- | --- |
| Description of the study sample regarding to number of participants, age, sex, health status, fitness levels, etc | 0 | n ≤ 25 and including less item than required for grade 1. |
|  | 1 | N ≥26 and at least age and gestational week. |
| Adequate assessment and report of physical fitness test. | 0 | Items for grade 1 are not included within the article. |
|  | 1 | Validity and/or reliability reported of test and detailed description of testing protocol. |
| Adequate assessment of health-related outcomes | 0 | Items for grade 1 are not included within the article. |
|  | 1 | Validity or reliability of the outcome measure reported and/or measurement procedure adequately described. |
| Adequate adjustment of confounders | 0 | No adjustment was done. |
|  | 1 | Adjustment of confounders such as age and sex were done. |
| Description of both number and reasons to withdrawal and dropout. | 0 | No description included. |
|  | 1 | Description included. |
