## Supplementary material for "Assessing physical fitness during pregnancy: validity and reliability of fitness tests, and relationship with maternal and neonatal health-related outcomes. A systematic review": ESM5_TableS6

**Electronic Supplementary Material 5 Table S6. Overview of studies included in the systematic review and description of physical fitness tests.**

| **Reference**  **(authors, year)** | **Sample Size (n)** | | **Gestation Weeks (SD) or range in weeks** | | | | **Mean age (SD), or range, in years** | | | **Fitness Test and Short Description** | |
| --- | --- | --- | --- | --- | --- | --- | --- | --- | --- | --- | --- |
| ***Cardiorespiratory Fitness*** | | | | | | | | | | | |
| ***Cycle-ergometer protocol*** | | | | | | | | | | | |
| Pomerance et al., (1974)^1^ | 54 | | 17.5-27 | | | | 35-37 | | | Ad hoc, steady-state test at 60 rpm at 450, 600 and 300 kpm. | |
| Erkkola, (1976)^2^ | 120 | | (2 weeks before term) | | | | 20-26 | | | 1) Ad hoc, incremental submaximal test at 150, 300 and 450 kpm/min. 2) Arstila ECG test. | |
| Morton et al, (1985)^3^ | 23 | | 40.15 (1.5) | | | | 28.5 (2.1) | | | Ad hoc, steady-state test at 40 to 50 rpm and at 300 kpm . min^-1^ for 6 min. | |
| Veille et al., (1985)^4^ | 17 | | 35 (2) | | | | 31 (1) | | | Ad hoc, incremental submaximal test at 50 and 60 rpm at 50W for 10-15 min to 70% HR max (no formula). | |
| Jovanovic et al., (1985)^5^ | 6 | | 37.1 (0.9) | | | | 28.5 (1.7) | | | Ad hoc, incremental submaximal self-administered test to 50% VO_2_ max or exertion equivalent to usual training. | |
| Wong & McKenzie, (1987)^6^ | 20 | | 3 time-points, (10-14; 22-24; 34-36) | | | | 29.13 | | | Ad hoc, incremental submaximal test at 50 rpm at 25, 50, 75 and 100 W for 5-6 min to 150 bpm. | |
| Kulpa et al., (1987)^7^ | 141 | | First trimester | | | | 18-34 | | | Bruce protocol to 75% of HR max. | |
| Carpenter et al., (1988)^8^ | 45 | | 29 (3.7) | | | | 25.2 (3) | | | Ad hoc, incremental test. 2 phases: a) *submaximal*: at 0, 30 and 60W for 6 min. b) *maximal*: at 60W to volitional fatigue. | |
| Moore et al.,, (1988)^9^ | 11 | | 21.3 | | | | 26.6 | | | Ad hoc, incremental submaximal test at free pace for 20 min to 60 to 75% HR max. (220-age). | |
| Sady & Carpenter, (1988)^10^ | 40 | | 29.2 (3.9) | | | | 25.9 (3.3) | | | 2 incremental tests: 1) *Submaximal test* at 0 W, 30 W and 60 W at 30%, 50% and 70% of VO_2_ max. 2) *Maximal test* increasing 10 W every 2-min stage to volitional fatigue. | |
| Artal et al., (1989)^11^ | 37 | | 29.8 (0.5) | | | | 28.3 (1.8) | | | Ad hoc, incremental maximal test at 25, 50, 75W and increments of 25W every 2-min stage until exhaustion. | |
| Hume et al., (1990)^12^ | 30 | |  | | | | 28 | | | Ad hoc, steady-state submaximal test at 60% VO_2_ max for 20min. | |
| Sady et al., (1990)^13^ | 9 | | 25.6 (3.0); | | | | 29 (4.9) | | | Ad hoc, incremental test. 2 phases: a) *submaximal*: at 0, 30 and 60 W for 18 min. 6-min each stage. b) *maximal*: incremental continuous to volitional fatigue. | |
| Field et al., (1991)^14^ | 13 | | 33 ± 2 | | | | 30 (4) | | | Modified Balke protocol to 70% HR max (no formula) | |
| Rafla & Beazely, (1991)^15^ | 21 | | 28-37 | | | | - | | | Ad hoc, incremental submaximal test from 60 rpm to 70% HR max (220-age) | |
| Bung et al., (1991)^16^ | 1 | | 3 time-points, (24, 28, 37) | | | | 25 | | | Ad hoc, incremental submaximal test from 15 W to 150 bpm. | |
| Young & Treadway, (1992)^17^ | 5 | | 33 (1) | | | | 29 (1) | | | Ad hoc, steady-state submaximal test at 50% VO_2_ max for 30 min. | |
| Clapp et al., (1993)^18^ | 120 | | 16-39 | | | | - | | | Ad hoc, steady-state submaximal test at 60% ± 3% VO_2_ max for 30 min. | |
| Lotgering et al., (1995)^19^ | 33 | | 3 time-points, 16.1 (1); 25 (0.7); 35 (0.6) | | | | 30.9 (0.7) | | | Ad hoc, incremental submaximal test. After 3 min at 15W, to increase 10 W every 30 sec until peak aerobic power. | |
| Artal et al., (1995)^20^ | 7 | | 33.86±1.46 | | | | 24.9 (2.18) | | | Ad hoc, incremental submaximal test. After 5 min per stage at 25, 50 and 75W, to increase 25W every 2 min to volitional fatigue. | |
| O’Neill, (1996)^21^ | 11 | | 35.8 (1.1) | | | | 30.3 (3.3) | | | 1) Ad hoc, steady-state test at 62.5 W for 15 min. 2) Ad hoc, steady-state test at 87.5 W for 15 min. 3) Ad hoc, steady-state test at 62.5 W for 30 min. | |
| Soultanakis et al., (1996)^22^ | 20 | | 27.1 (1.3) | | | | 31.4 (1.5) | | | 1) Incremental maximal with modified Balke protocol, increasing 25 W every 2-min at 60 rpm to VO_2_max 2) Ad hoc, steady-state submaximal test during 1 hour at 50%-60% VO_2_max at 60 rpm. | |
| Manders et al., (1997)^23^ | 12 | | 29-32 | | | | 20-36 | | | Ad hoc, incremental maximal test. After 5-min per stage at 50W, to increase 25 W/min to volitional fatigue. | |
| Kemp et al., (1997)^24^ | 23 | | 33 (1) | | | | - | | | Ad hoc, incremental maximal test at 20 W for 4 min. Then, increasing 20 W/min until exhaustion. | |
| McGrath et al., (1999)^25^ | 41 | | 3 time-points: 17.45 (0.45); 26.5 (0.2) and 37.15 (0.15) | | | | 29.4 (0.85) | | | Ad hoc, steady-state test with three 6-min stages and exercise brief (<5-min) between them. 1) 20 W to 110 bpm, 2) 45 W to 130 bpm 3) 70 W to 150 bpm. | |
| Brenner et al., (1999)^26^ | 20 | | 27.0 (1.0) and 37.0 (1.0) | | | | 29 (3.35) | | | Ad hoc incremental submaximal test for 3 min without resistance, then, increased 30 W/min to 170 bpm or RPE of 18. | |
| MacPhail et al., (2000)^27^ | 23 | | 32 (4) | | | | 20-40 | | | Idem Kemp et al., (1997) | |
| Heenan et al., (2001)^28^ | 28 | | 34.7 (0.4) | | | | 30.8 (1.5 | | | Idem Kemp et al., (1997) | |
| Kennelly et al., (2002)^29^ | 22 | | 32.1 (1.4) | | | | 25.9 (4.9) | | | Ad hoc incremental maximal test. After 2-min at 30 W, increasing 10 W/min at 50-60 rpm to achieve AT. | |
| Heenan & Wolfe, (2003)^30^ | 22 | | 37.0 (0.2) | | | | 29 (1.1) | | | 1) Ad hoc, incremental submaximal test at 20 W for 4 min. Then, to increase 20 W/min until 170 bpm. 2) Ad hoc, incremental ramp test from 0 W increasing work rate in 30-sec periods to 70 or 110% of VT. | |
| Wolfe et al., (2003)^31^ | 18 | | 3 time-points: 19.2 (0.8) 27.8 (0.3) 37.0 (0.3) | | | | 28.3 (0.25) | | | Ad hoc incremental submaximal test for 3 min of no resistance. Then, to increase 30 W/min to 170 bpm or RPE of 18. | |
| Lindqvist et al., (2003)^32^ | 14 | | 5 time-points: 8, 15, 22, 29 and 36. | | | | 29 (5) | | | Ad hoc incremental submaximal test for 2 min of no resistances. Then, to increase 20 W every 2 min to HR max or pulse oximetry below 95%. | |
| Lynch et al., (2003)^33^ | 23 | | 16, 20, 24, 28, 32, 36 | | | | 28.7(4) | | | Ad hoc incremental submaximal test at 60 rpm no resistance. Then, to increase 0.5 or 1 kP during two 3-min stages to 130 ± 5 bpm and 1 stage more to 145 ± 5 beats/min. | |
| Heenan et al., (2003)^34^ | 39 | | 37.0 (0.2) | | | | 28.5 (1.4) | | | 1) Ad hoc incremental submaximal test at 20 W for 4 min. Then, to increase 20 W/min until 170 bpm. 2) Ad hoc, incremental ramp test from 0 W increasing work rate in 30-sec period. (70 or 110% of VT) | |
| Pirhonen et al., (2003)^35^ | 14 | | 5 time-points: (8, 15, 22, 29, 36) | | | | 29.2 (4.6) | | | Ad hoc incremental submaximal test at 0 W and 20 W for 2 min. Then, to increase at 40 W and thereafter 30 W/min to 85% HR max (220-age) or pulse oximetry below 95%. | |
| Kardel, (2005)^36^ | 41 | | 17, 30, 36 | | | | 27.7 (1.95) | | | Ad hoc, incremental maximal test for 3-min stages at 50 W, 100 W and 150 W. After a rest, (no longer than 3-min) work maximally (200-280 W) for the first 30 seconds of 3-min stages. | |
| McAuley et al., (2005)^37^ | 14 | | 17.05 (2.05) | | | | 29.9 (0.85) | | | Ad hoc incremental submaximal and maximal test for 4 min at 20 W at 60-80 rpm. Then, to increase 20 W/min to 170 bpm or volitional fatigue. | |
| Weissgerber et al., (2006)^38^ | 11 | | 7 - 22 | | | | 25-40 | | | Ad hoc incremental submaximal test at 20 W for 4 min. Then, increasing 5 W/min until volitional fatigue or 170 bpm. | |
| Jensen et al., (2007)^39^ | 22 | | 3 time-points: 19.7 (1.2), 28.2 (0.3), 36.3 (0.3) | | | | 30.9 (0.9) | | | Idem test 1 of Heenan & Wolfe (2003). | |
| Jensen et al., (2008)^40^ | 15 | | 34-38 | | | | 30.6 (1.0) | | | Ad hoc incremental maximal test from 6-min resting period. After 25 W/2 min at cadence of 60 and 70 rpm to the point of volitional fatigue. | |
| Kardel et al., (2009)^41^ | 40 | | 35-37 | | | | 20-40 | | | Ad hoc incremental maximal test at 20 W for 2 min. Then, to increase (8-12 min) to ramp up 10% of the predicted maximal load. | |
| Ong et al., (2009)^42^ | 12 | | 2 time-points: 18 and 28. | | | | 30 (4) | | | Ad hoc, incremental submaximal test increasing 25 W/min to 75 % HR Max (220-age). | |
| Thorell et al., (2010)^43^ | 520 | | 4 time-points: 10.9, 24.0, 29.7, 36.5. | | | | 29.0 (4.4) | | | Ad hoc incremental submaximal test at 50 or 75 W (based on previous level) increasing 25 W/min to ≥125 bpm. | |
| Rojas-Vega et al., (2011)^44^ | 20 | | 34±1.6 | | | | 35.2 (3.6) | | | Ad hoc incremental submaximal test free of cadence and speed for 2 min. Then, to increase 25 W/ 2 min at 60 rpm to 150 bpm. | |
| Thorell et al., (2015)^45^ | 520 | | 10.9 | | | | 29.6 | | | Idem Thorell et al. (2010). | |
| Kim et al., (2015)^46^ | 32 | | 13-35 | | | | 24.8 (2.5) | | | Ad hoc, steady-state test with three 20-min phases:1) standing 2) pedalling at 50 W for 20 min 3) sitting. | |
| Nakagaki et al., (2016)^47^ | 20 | | 25.1(6.3) | | | | 33.7(4.2) | | | Ad hoc, incremental submaximal test at 50 rpm to 160 bpm or impossibility to maintain the pedalling rate. | |
| Jedrzejko,et al., (2016)^48^ | 22 | | 37-41 | | | | 24.4 (3.92) | | | Ad hoc, incremental submaximal test on supine cycle divided into three 4-min constant stages increasing from 25 W to 75 W. | |
| Sussman et al., (2019)^49^ | 23 | | 2 time-points: 14-15 and 33-34 gw | | | | 30 (3) | | | YMCA protocol. Incremental test on semirecumbent to 60-80% HR_Max_ or RPE of 14 out 20. | |
| Purdy et al., (2019)^50^ | 63 | | 4 groups: 10-12, 20-27, 30-37 | | | | 30.5 (4.5) | | | Ad hoc, incremental maximal test on recumbent cycle at 25 W at 50 rpm for 5 min. Then, to increase 25 W/min at same speed to volitional fatigue. | |
| Bilodeau et al., (2019)^51^ | 58 | | 3 time-points: 16.5 (1.0), 35.6 (0.9); 39.8 (1.1) gw | | | | 30 (3.7) | | | Modified Bruce ramp protocol. | |
| Matenchuk et al., (2019)^52^ | 47 | | 4 groups: nonpregnant; 1^st^ trimester, 2^nd^ trimester, 3^rd^ trimester | | | |  | | | Ad hoc, incremental maximal test at 25 W at 50 rpm for 5 min. Then, to increase 25 W/min to volitional fatigue. | |
| Correa et al., (2020)^53^ | 48 | | 2 time-points: 18; 36 gw. | | | |  | | | Ad hoc, incremental ramp submaximal test at 4 W for 4 min. Then, to increase 20 W/min until symptom limitation or HR_Max_ (220-age). | |
| Bijl et al., (2020)^54^ | 40 | | 11 (1) | | | |  | | | Ad hoc, incremental submaximal on an upright cycle ergometer for 3-min at 40rpm. Then, to increase at 60-70 rpm at 25 W followed by a rise of 5 Watt in every 12-s to 70% HR_max_ (Tanaka formula). | |
| ***Treadmill protocol*** | | | | | | | | | | | |
| Sibley et al., (1981)^55^ | 13 | | 2 time-points: 21.9 (2.3); 33.9 (2.3) | | | | 24.3 (1.4) | | | Balke protocol to 140 bpm. | |
| Veille, (1985)^4^ | 17 | | 35 (2) | | | | 31 (1) | | | Ad hoc, incremental submaximal walking test to 70% HR_Max_ (no equation to calculate HR_max_ shown). | |
| Lewis et al., (1988)^56^ | 28 | | 2 time-points: 22 wg and 30 wg | | | | 27.8 (3.3) | | | Modified Balke protocol. | |
| Artal et al., (1989) ^11^ | 37 | | 30.3 (1.9) | | | | 25.9 (2.5) | | | Modified Balke protocol. | |
| Clapp, Little & Capeless, (1993)^18^ | 120 | | 16-39 | | | | NR | | | 1) Ad hoc, steady-state test at 40% ± 3% VO_2_ max for 30 min. 2) Idem at 60% ± 3% VO2max. | |
| Winn et al., (1994)^57^ | 12 | | 26-36 | | | | 32 (4) | | | Modified Bruce Protocol to 75% HR Max (220-age). | |
| Marquez-Sterling et al., (2000)^58^ | 15 | | 19.1 (2.15) | | | | 29.5 (3.1) | | | Ad hoc incremental test at 4 km/h and 0% grade for 2-min. Then, increasing 6 km/h and 2.5% every 2-min to 150 bpm. | |
| Santos et al., (2005)^59^ | 72 | | 17.9 (3.6) | | | | 27.3 (4.65) | | | Ad hoc, incremental ramp test from 2.4 km/h and 0% grade to AT. | |
| Yeo et al., (2005)^60^ | 9 | | 19 (5) | | | | 30 (3) | | | 2 Cornell Protocol (85%MHR; Karvonen formula) with 2 systems (VO2000 and CPX/D). | |
| Mottola et al., (2006)^61^ | 156 | | 16-22 | | | | 30.8 (3.7) | | | Modified Balke protocol with this equation VO_2_ peak (predicted) = (0.055*peak HR) + (0.381* incline) + (5.541* speed (mph)) + (-0.090*BMI) -6.846 : incremental walking test at 3 mph for 5 min, 0% grade. Then, increase 2% every 2 min. Max inclination permitted 12% grade. Then, increasing speed 0.2 mph every 2-min to volitional fatigue. | |
| Davenport, et al., (2008)^62^ | 106 | | 16-20 | | | | 20-39 | | | Modified Balke protocol. Idem Mottola et al., 2006. | |
| Oliveria et al., (2012)^63^ | 187 | | 3 time-points: 13, 20, 28. | | | | 24.7 (5.5) | | | Modified Balke protocol. Idem Mottola et al. (2006). | |
| Ruchat et al., (2012)^64^ | 44 | | 2 time-points: 16-20 and 34-36 | | | | 30.8 (4.2) | | | Modified Balke protocol. Idem Mottola et al. (2006). | |
| Szymanski, (2012)^65^ | 45 | | 30.4 (1) | | | | 33.36 | | | Modified Balke protocol. Idem Mottola et al. (2006). | |
| Salvesen et al., (2012)^66^ | 6 | | 25.5 | | | | 32 | | | Ad hoc, incremental maximal test at 6% grade increasing speed in periods of 1km/h every 5-min to volitional fatigue. | |
| Mottola et al., (2013)^67^ | 40 | | 35.7 (0.4) | | | | 33.5 (0.7) | | | Ad hoc, steady-state test for 40-min, preceded by a 5-min warm-up increasing speed and inclination to 95% VT. | |
| Bisson et al., (2013)^68^ | 65 | | 16 | | | | 29.9 (4.5) | | | Modified Balke protocol. | |
| LeMoyne et al., (2014)^69^ | 67 | | 1st trimester, 2nd trimester, 3rd trimester | | | | 29.6 (5.5) 30.1 (3.1) 32.3 (3.7) | | | Ebbeling single-stage submaximal treadmill walking test. | |
| Bisson et al., (2014)^70^ | 61 | | 16 (0.6) | | | | 30.0 (4.5) | | | Modified Balke protocol. | |
| Marshall et al., (2015)^71^ | 51 | | 3 time-points: 20, 32 | | | | 29.2(5.3) | | | Ad hoc, incremental submaximal test at 0% grade and 3.21 km/h for 5-min. Then, two 5-min stages with speed and grades self-administered to moderate (brisk walk) and vigorous (jog/run) respectively. | |
| Santos et al., (2016)^72^ | 28 | | 30.51 (3.3) | | | | 26 (6.9) | | | Modified Balke protocol. | |
| Hesse et al., (2018)^73^ | 25 | | 22.1 (1.4) | | | | 30 (3.6) | | | Bruce protocol until volitional fatigue. | |
| Baena-García et al., (2020)^74^ | 127 | | 16 | | | | 32.9 (4.6) | | | Modified Bruce protocol until 85% HR_Max_ | |
| Dobson et al., (2020)^75^ | 22 | | 3 time-points: Early- (13–18 gw), mid- (24–28 gw) and late-pregnancy (34–37 gw). | | | | 31.4 (3.7) | | | Submaximal incremental Walking Exercise Test (SWET) during 21-min on a treadmill. From 3.2 km/hr at 4 min at 2% grade, to increase 2% every 3 min over seven stages. | |
| ***On track*** |  | |  | | | |  | | |  | |
| Bung et al., (1991)^16^ | 1 | | 3 time-points(24, 28, 37) | | | | 25 | | | Ad hoc, maximal test. 3 sprints of 200 m and one of 100 m on track | |
| Da Silva et al., (2010)^76^ | 74 | | 37 | | | | 21.5 | | | 6-minute walk test. | |
| Ramírez-Vélez et al., (2011)^77^ | 64 | | 2 time-points: 18.6 (3.4) and 16 weeks later. | | | | 19.5 (2.3) | | | 6-minute walk test. | |
| Hjorth et al., (2012)^78^ | 304 | | 25.0 (7.3) | | | | 23.0 | | | Ad hoc, steady-state walking test for 250 m on ground level at their normal walking pace. | |
| Price et al., (2012)^79^ | 62 | | 5 time-points: 12–14, 18–20, 24–26 and 30–32 | | | | 29.05 | | | Ad hoc test walking or running as fast a as possible within comfort zone at a steady pace. Power = (weight x distance) / time. | |
| Radzikowska et al., (2017)^80^ | 45 | | 3-7 | | | | 24-36 | | | 6-minute walk test. | |
| Oviedo-Caro et al., (2018)^81^ | 134 | | 20 | | | | 32.5 (4.2) | | | 6-minute walk test. | |
| Dennis et al., (2019)^82^ | 300 | | 37 (1.3) | | | | 31 (4.2) | | | 6-minute walk test. | |
| Amola et al., (2019)^83^ | 34 | | 3rd trimestre | | | | 25.1 (7.5) | | | 6-minute walk test. | |
| Birnbaumer et al., (2020)^84^ | 39 | | 26 (7) | | | | 26 (3.4) | | | Ad hoc, incremental walking test on a 400 Walking speed was paced by audio every 10 m and started at 3 km/h. Then, to increase 0.5 km/h every 50 m to participants were unable to walk the given pacer speed. | |
| ***Step Protocol*** |  | |  | | | |  | | |  | |
| Dibblee & Graham (1983)^85^ | 16 | | 3 time-points: (the last month of each trimester) | | | | 23-31 | | | | Canadian Home Fitness Test. |
| Williams, Reilly et al. (1988)^86^ | 16 (10 pregnant and 6 non-pregnant) | | First, second and third trimester. | | | | 25.6 (3.6) | | | Ad hoc, incremental test at 115, 135, and 155 bpm for 5 min. | |
| Melzer et al., (2010)^87^ | 44 | | 38.27 | | | | 31 (5.6) | | | Ad hoc, incremental test at 15-32.5 body lifts per minute (rate of change: 2.5 body lifts/ min^2^). Mechanical power was calculated as: 9.81 m/s^2^ x step height (m) x lift frequency (number of body weight lifts/ min) and expressed in J/min/kg | |
| ***Muscular Fitness*** |  | |  | | | |  | | |  | |
| Baker & Johnson (1994)^88^ | 200 | | NR | | | | 28-32 | | | Hand Grip Sphygmomanometer Test: Pressing an inflated cuff of for 30-sec to MVCF over 3-min period. | |
| Rogers & Tomilson (1998)^89^ | 20 | | NR | | | | 5 times: 12, 18, 24, 30, 36 | | | Hand Grip Sphygmomanometer Test at 30% of MVCF for 2-min. | |
| Feiner et al. (2000)^90^ | 34 | | 22-36 | | | | 22-35 | | | Isometric Hand-Grip Test with dominant hand for 3 min at one-third of MVCF. | |
| Gutke et al., (2008)^91^ | 301 | | 12-18 | | | | 29 | | | 1) Maximal voluntary isometric hip extension test with a fixed sensor holding a sling around the thigh and pulling for 5 sec during 3 reps with 5-10-sec of rest. 2) Isometric back flexors endurance: Maintaining an abdominal crunch for a maximum of 120 sec. | |
| Thorell et al., (2010)^43^ | 520 | | 1 time-points: 10.9 | | | | 29.0 (4.4) | | | Sit-up test. Supine position with the knees at a 90º angle and the feet flat on the floor. 3 sets per 5 repetitions, without a rest or to stop when they were unable to perform of 15 repetitions of sit-ups. | |
| O’Connor et al., (2011)^92^ | 32 | | 21-25 | | | | 18-38 | | | Ad hoc 5 tests: 1) Seated leg press; 2) Leg curls; 3) Leg extension; (4) Lat pull; (5) Back extension. | |
| Hjorth et al., (2012)^78^ | 304 | | 25.0 (7.3) | | | | 23.0 | | | Hand-Grip maximal strength test twice on dominant and non-dominant side alternatively. | |
| Price et al. (2012)^79^ | 62 | | 5 time-points:  12–14, 18–20, 24–26 and 30–32 | | | | 29.1 | | | Ad hoc test. Lifting a 7-kg medicine ball from the floor to waist height as many times possible for 1 min. | |
| Bisson et al. (2013)^68^ | 65 | | 16 | | | | 29.9 (4.5) | | | Hand-Grip maximal strength test twice on dominant and non-dominant side alternatively. Adjusting the handle of dynamometer. | |
| Atay et al., (2015)^93^ | 37 | | 2 time-points: 20 and 32 | | | | 29.6 (5.9) | | | Hand-Grip maximal strength test in a sitting position. | |
| Petrov et al., (2015)^94^ | 92 | | 2 time-points: 13 and 35 | | | | 30.7 (3.5) | | | Hand-Grip isometric peak strength. | |
| Wickboldt (2015)^95^ | 43 | | 32 (4) | | | | 37-42 | | | Hand-Grip maximal strength test during the uterine contraction. | |
| Kalliokoski et al. (2016)^96^ | 51 | | NR | | | | 28.3(6.4) | | | 1) Hand-Grip maximal strength test for 10 sec 3-times in each hand.  2) Ad hoc upper leg performance test through 3 movements: a) To rise once after a squat b) to stand on one leg for 30 sec 3) Trendelenburg’s test. It was evaluated able or unable. | |
| Ngaka et al. (2016)^97^ | 50 | | >37 | | | | 28.8 (5.7) | | | Hand-Grip maximal strength test in a supine position. | |
| Rodriguez-Díaz et al., (2017)^98^ | 105 | | 24-30 | | | | 32.2 (4.7) | | | Hand-Grip maximal strength test for each hand. | |
| Zelazniewicz, (2018)^99^ | 95 | | 3 time-points (once in each trimester) | | | | 29.6 (3.4) | | | Hand-Grip maximal strength test twice on dominant and non-dominant side alternatively. | |
| Takeda et al., (2019)^100^ | 21 | | 22 and 23.25 gw. | | | | 32 (3.3) | | | 1) Toe grip dynamometer 2) Hand-held dynamometer fixed to the legs of the chair with a belt not stretchable to assess quadriceps strength . | |
| Baena-García et al., (2020)^74^ | 156 | | 16 | | | | 32.9 (4.6) | | | 1) Hand-grip maximal strength twice on dominant and non-dominant side alternatively with 30 sec rest between them.  2) 30-sec Chair Stand Test | |
| Yenisehir et al., (2020)^101^ | 167 | | Second and third trimester. | | | | 28.4 (4.6) | | | 5 Times Sit to Stand test, 5 repetitions of sit-to-stand maneuver as fast as possible with fold arms across the chest. | |
| ***Flexibility*** | | | | | | | | | | | |
| Gilleard et al. (2002)^102^ | 21 | | | | 4 time-points: 18 or less, 24, 32, 38 | | 21-40 | | | 3 tests measured with Expert Vision™ Motion Analysis System: 1) Seated and standing forward flexion 2) Seated and standing side-to-side flexion 3) Seated axial rotation | |
| Marnach et al. (2003)^103^ | 46 | | 3 time-points: 8-12, 16-22, 34-36. | | | | 28.8 (0.8) | | | Wrist flexion-extension and medial-lateral deviation using goniometer | |
| Garshasbi et al. (2005)^104^ | 212 | | 17-22 | | | | 26.4 (4.7) | | | Side bending test: Both sides. | |
| rice et al., (2012)^79^ | 62 | | 5 time-points: 12–14, 18–20, 24–26 and 30–32. | | | | 29.1 | | | Sit-and-reach test. | |
| Lindgren et al. (2014)^105^ | 200 | | 3 time-points: 11, 24 and 36. | | | | 28.4 (5.9) | | | Ad hoc machine to test passive abduction of the left fourth finger. | |
| Atay et al., (2015)^93^ | 37 | | 2 time-points: 20 and 32, | | | | 29.6 (5.9) | | | Back scratch test. | |
| Rodriguez-Díaz et al., (2017)^98^ | 105 | | 24-30 | | | | 32.2 (4.7) | | | Isquiosural flexibility test by goniometer. | |
| Cherni et al., (2019)^106^ | 17 | | 3 occassions: first, second and third trimester | | | | 36 (2) | | | 4 tests measured with optoelectronical system: 1) Extensometer of the metacarpophalangeal joint of the index. 2) Figertrip to floor test: from 20cm platform, to reach the floor with knees extended; 3) Sit-and-reach test adapted on delivery bed 4) Beighton score | |
| Baena-García et al., (2020)^74^ | 156 | | 16 | | | | 32.9 (4.6) | | | Back Scratch | |
| ***Balance*** | | | | | | | | | | | |
| ***Stabilometry – On force platform or pressures platform*** | | | | | | | | | | | |
| Butler et al., (2006)^107^ | | 12 | | 3 time-points: 11-14, 19-22, 36-39 | | | | 32.9 (5.5) | | Standing with eyes open and eyesclosed for 30 sec each. 3 trials. 1 piece. Force Platform. | |
| Ribas et al., (2007)^108^ | 60 | | 3 time-points: 1) Up to 12 week 2) 13-24 3) Upwards of 25 weeks | | | | 23.3 (4.8) | | | Standing with bipedal support and eyes open for 5 sec. 2 pieces at 40 Hz. | |
| Nagai et al., (2009)^109^ | 43 | | 30.3 (0.8) | | | | 33 (0.65) | | | Standing with feet parallel, gazing a black 12-cm circle fixed at a 1.5 m distance with eyes open and eyes closed for 1 min each. 1 piece. | |
| Oliveira et al., (2009)^110^ | 20 | | 3 time-points: 15.1 (1.8); 24.0 (2.4); 34.5 (2.5) | | | | 28.7 (6.2) | | | Standing with 4 protocols at 50Hz and 2-min rest periods between them: 1) Eyes open with feet comfortably apart; 2) Eyes closed with feet comfortably apart; 3) Eyes open with feet together; 4) Eyes closed with feet together. 1 piece. | |
| Karadag-Saygi et al., (2010)^111^ | 35 | | 33 (3) | | | | 29.8 (4.5) | | | Standing for 60 sec. | |
| Yu et al., (2013)^112^ | 21 | | NR | | | | 30.2 (3.05) | | | Standing with heels on a line at 1.0 m from visual target with visual tasks and inspection tasks. | |
| Ersal et al., (2014)^113^ | 69 | | 2 time-points: 20.9 (1.2) and 35.8 (1.5) | | | | 28.3 (5.0) | | | Standing with feet hip-width apart and staring straight ahead on Equitest platform. | |
| 7Opala-Berdzik et al., (2014)^114^ | 31 | | 36.2 (1.2) | | | | 28.2 (3.6) | | | Standing with arms at both sides and in a comfortable stance on a stable force platform with eyes open and eyes closed for 2 trials of 30-sec and 1-min rest between them. | |
| Opala-Berdzik et al., (2015)^115^ | 45 | | 2 time-points: 13.1 (2.5) and 36.2 (1.2) | | | | 28.2 (3.6) | | | Idem Opala-Berdzik et al., (2014) | |
| Ozturk, (2016)^116^ | 68 | | 31.5 (4.73) | | | | 30.3 (3.6) | | | Standing and arms extended in 6 different positions for 32-sec: 1) facingforward eyes open and eyes closed; 2) Eyes closed head rotated at 45º to the right 3) Idem 45º to the left; 4) Eyes closed, head tilted at 30º backward and 5) Idem 30º forward; 6) Standing on an unstable cushion, facing forward eyes open and eyes closed. 4 pieces. | |
| Shibayama et al., (2016)^117^ | 161 | | 28-33 | | | | 33.3 (4.7) | | | Standing and feet together for 30 sec on force platform. 1 piece. | |
| Takeda et al., (2018)^118^ | 100 | | 2^nd^ and 3^rd^ trimester | | | | 20-30 | | | Standing with the medial malleoli 100 mm apart for 10-sec. Then, moving forward, backward, right and left for 10-sec each. 2 pieces. | |
| Moreira et al., (2017)^119^ | 30 | | 1^st^ and 3^rd^ trimester | | | | 26.8 (5.1) | | | Standing with each foot positioned on each triaxial force plate (feet apart by ~20 cm) and arms along the body with eyes open focusing on a target located ~2 m in front and eyes closed for 3 trails of 60-sec each and 2-min rest. 2 pieces. | |
| Opala-Berdzik et al., (2018)^120^ | 70 | | 10.8 (1.6) | | | | 28.6 (4.4) | | | Standing with arms at both sides and in a comfortable stance on a stable force platform, with eyes open looking straight ahead at a wall 3m away for 2 trials of 30-sec and 1-min rest between them. | |
| Catena et al., (2019)^121^ | 17 | | 9 time-points: 16-20 gw, 36-40 gw and 1 time per month up to 7 months postpartum | | | | 28.9 (4.0) | | | 2 trials: 1) quiet static in anatomical position for 10 s on a force plate; 2) Idem 1 on a back-board spanning two force plates. | |
| Fontana et al., (2020)^122^ | 24 | | 23 (3) | | | | 30 (6) | | | Standing barefoot two-legged stance with arms at both sides with eyes open at 2 m from a cross placed on a wall at eye level during 3 x 30s trials with 30 s rest intervals. The mean was retained on force platform. | |
| Valerio et al., (2020)^123^ | 40 | | 30.8 (3.9) | | | | 28 (2.5) | | | Standing barefoot with freestanding supports inside the platform and arms by their sides. And staring at a mark on the opposite wall. 3 trials with the eyes open and three trials with eyes closed, with 30 s rest intervals. | |
| Takeda et al., (2019)^100^ | 21 | | 22 and 23.25 gw. | | | | 32 (3.3) | | | Standing barefoot on 2 stabilometers. 3 trials: 1) 10-sec standing position; 2) 10-sec moving in the anterior position; 3) 10-sec moving in the posterior position. | |
| ***Others*** | | | | | | | | | | | |
| Atay et al., (2015)^93^ | 37 | | | | 2 time-points: 20 gw and 32 gw | | 29.6 (5.9) | | | | One-legged stand test. |
| ***Dynamic Balance*** | | | | | | | | | | | |
| ***On platforms*** | | | | | | | | | | | |
| Davies et al., (2002)^124^ | 150 | | Day of labour | | | 30.2 (5.8) | | | | Balance Master Platform Tests: 1) Sit to Stand; 2) Walk Test, 3) Step and Quick Turn, 4) Step Up and Over. | |
| Karadag-Saygi et al., (2010)^111^ | 35 | | 33 (3) | | | | 29.75 (4.5) | | | Walking barefoot 4 m. | |
| McCrory et al, (2010)^125^ | 81 | | 2 time-points: 20.9 (1.2) and 35.8 (1.5) | | | | 28 (5.7) | | | The Motor Control Test protocol with translational perturbations. Equitest posture platform. | |
| Branco et al., (2013)^126^ | 22 | | 27 (1.3) | | | | 32.5 (2.6) | | | Walking barefoot for 10 m between 2 points in a straight line at a natural and comfortable speed for 3 min. | |
| Cakmak et al., (2014)^127^ | 41 | | 6-12 | | | | 26.5 (4.7) | | | Standing with knee flexed, arms placed across the chest and glare fixed ahead with open eyes on a movable platform provides up to 20º of surface tilt in a 360º range of motion for 3 trails of 20 sec each. | |
| Inanir et al., (2014)^128^ | 110 | | 3 groups: 1^st^ trimester, 2^nd^ trimester and 3^rd^ trimester. | | | | 24.7 (5.2) | | | Idem to Cakmak et al., (2014) | |
| ***3-D Camera motion capture system*** | | | | | | | | | | | |
| Wu et al., (2004)^129^ | | 25 | | 27 | | | | 33.1 | | Walking on a treadmill at different velocities (incrementing 0.11 m/s, from 0.17 up to 1.72 m/s; for 3 min at each level). | |
| Forczeck et al., (2012)^130^ | 13 | | NR | | | | 29.2 (3.5) | | | Walking barefoot at a self-selected speed across the room during 15 gait cycles.10 | |
| Takeda et al., (2012)^131^ | 16 | | 24.85 (1.95) | | | | 35 (1.4) | | | Stand-to-sit motion assessing the time taken to sit down; the leg joint moment; the antero-posterior and vertical floor reaction forces; and the range of motion of the lower limbs and trunk. | |
| Gottschall et al., (2013)^132^ | 13 | | 2 time-points: 20 and 32 | | | | 31.3 (4.5) | | | Walking along 25 m on a custom-built portable apparatus composed of a 2.4 m ramp inclined at 15° continuous with a 4.8 m plateau. | |
| McCrory et al., (2014)^133^ | 69 | | 28.35 (1.35) | | | | 28.0 (5.7) | | | Walking along the 8-m runway. | |
| Krkeljas, (2018)^134^ | 35 | | 3 time-points: 9-12 gw; 20-22 gw and 28-32 gw. | | | | 27 (6.1) | | | Walking on a straight line, at a self-selected pace along the 15-m walkway. | |
| Catena et al., (2019)^135^ | 15 | | 5 time-points: 16-20; 20-24, 24-28; 28-32; 32-36 gw. | | | | 29.3 (3.7) | | | 60-second trial of semi-continuous stand-to-sit motion. 54 reflective markers were adhered to body land. | |
| Catena et al., (2019)^136^ | 15 | | 7 time-points: 12-16; 16-20; 20-24, 24-28; 28-32; 32-36; 36-40 gw | | | | 28.1 (4.3) | | | Walking on a treadmill for 60 seconds at a self-selected comfortable speed. | |
| Forczek et al., (2019)^137^ | 30 | | 3 time-points: 12, 25, 36 gw. | | | | 30.3 (3.4) | | | Walking barefoot at a self-selected speed during 12m intervals. 10 gait cycles. | |
| Forczek et al., (2019)^138^ | 14 | | 2 time-points: pre-pregnancy; 1^st^ trimester | | | | 20-40 | | | Walking barefoot across room at a self-selected during 50 m with 1 min rest intervals. 10 gait cycles. | |
| Catena et al., (2020)^139^ | 23 | | 5 time-points: 18, 22, 26, 30, 34 gw. | | | |  | | | Walking on a treadmill for 60 seconds at a self-selected comfortable speed. | |
| Gimunova et al., (2020)^140^ | 41 | | 4 time-points: 14, 28, 37 gw | | | | 30.5 (4.1) | | | Walking barefoot along a 6-meter walkway at a self-selected. | |
| McCrory et al., (2020)^141^ | 95 | | 2 time-points: 2^nd^ and 3^rd^ trimester | | | | 28.4 (5.5) | | | Walking along the 8m laboratory runway until walking speed stabilized. | |
| Rothwell et al., (2020)^142^ | 17 | | 2 time-points: 16-20; 36-40 gw | | | | 22-37 | | | Walking on a treadmill for 60 seconds at a self-selected comfortable speed. | |
| Forczek et al., (2019)^143^ | 36 | | 3 time-points: 12; 25; 36 gw | | | | 30.3 (3.4) | | | Walking across the room 50 m with 1-min rest interval. 10 gait cycles. | |
| ***Others*** | | | | | | |  | | |  | |
| Sawa et al., (2015)^144^ | 27 | | 2 groups: early pregnancy (≤27 gw) or late pregnancy (≥27 gw) | | | | | | 30.9 (4.2) | Walking at self-pace speed along a 15-m smooth, horizontal corridor. It was recorded with 2 wireless motion-recording-sensor units and one piezo-resistive triaxial accelerometer. | |
| Błaszczyk et al., (2016)^145^ | 28 | | 1^st^ trimestrer and 3^rd^ trimester | | | | 28.2 (3.4) | | | Walking along 10-m long walkway (back and forth 10 times) at self-space speed. It was recorded by custom made; self-adhesive copper foil electrodes attached to the soles of their shoes. | |
| ***Speed*** | | | | | | | | | | | |
| Evensen et al., (2015)^146^ | 17 | | 28.7 (7.4) | | | | 31.1(2.3) | | | Ten-metres Timed walk Test (10mTWT) | |
| Evensen et al., (2016)^147^ | 18 | | 28.9 (7.3) | | | | 31.4 (2.7) | | | 10mTWT | |
| ***Multicomponent*** | | | | | | | | | | | |
| Evensen et al., (2015)^146^ | 17 | | 28.7 (7.4) | | | | 31.1(2.3) | | | Timed Up and Go Test (TUG) | |
| Evensen et al., (2016)^147^ | 18 | | 28.9 (7.3) | | | | 31.4 (2.7) | | | TUG | |
| Christensen et al., (2019)^148^ | 74 | | 23 | | | | 31.2 (3.7) | | | TUG | |

Ad hoc: test designed specifically for that study; NR: Not reported; PFS: Physical Fitness Score; kpm: kilopoundimeter; min: minutes; sec: seconds; HR_Max_: Heart Rate Maximum; VO_2_ max: oxygen consumption maximum; RPE: rate of perceived exertion; AT: anaerobic threshold; ICC: intraclass correlation coefficient; MVCF: maximal voluntary contraction force; HGS: hand-grip strength; m: meters; mm: millimeters; FP: force platform; PP: pressure platform.

1.	Pomerance JJ, Gluck L, Lynch VA. Physical fitness in pregnancy: its effect on pregnancy outcome. *Am J Obstet Gynecol*. 1974;119(7):867-876. Accessed November 14, 2018. https://www.sciencedirect.com/science/article/pii/0002937874900015?via%3Dihub

2.	Erkkola R. The influence of physical training during pregnancy on physical work capacity and circulatory parameters. *Scand J Clin Lab Invest*. 1976;36(8):747-754. doi:10.3109/00365517609081933

3.	Morton MJ, Paul MS, Campos GR, Hart M V., Metcalfe J. Exercise dynamics in late gestation: Effects of physical training. *Am J Obstet Gynecol*. 1985;152(1):91-97. doi:10.1016/S0002-9378(85)80187-3

4.	Veille J-CC, Hohimer ARR, Burry K, Speroff L. The effect of exercise on uterine activity in the last eight weeks of pregnancy. *Am J Obstet Gynecol*. 1985;151(6):727-730. doi:10.1016/0002-9378(85)90505-8

5.	Jovanovic L, Kessler A, Peterson CM. Human maternal and fetal response to graded exercise. *J Appl Physiol*. 1985;58(5):1719-1722. doi:10.1152/jappl.1985.58.5.1719

6.	Wong SC, McKenzie DC. Cardiorespiratory fitness during pregnancy and its effect on outcome. *Int J Sports Med*. 1987;8(2):79-83. doi:10.1055/s-2008-1025645

7.	Kulpa PJ, White BM, Visscher R. Aerobic exercise in pregnancy. *Am J Obstet Gynecol*. 1987;156(6):1395-1403. doi:10.1016/0002-9378(87)90006-8

8.	Carpenter MW, Sady SP, Hoegsberg B, et al. Fetal heart rate response to maternal exertion. *JAMA J Am Med Assoc*. 1988;259(20):3006-3009. doi:10.1001/jama.259.20.3006

9.	Moore DH, Jarrett JC 2nd, Bendick PJ. Exercise-induced changes in uterine artery blood flow, as measured by Doppler ultrasound, in pregnant subjects. *Am J Perinatol*. 1988;5(2):94-97. doi:10.1055/s-2007-999663

10.	Sady SP, Carpenter MW, Sady MA, et al. Prediction of VO2max during cycle exercise in pregnant women. *J Appl Physiol*. 1988;65(2):657-661. doi:10.1152/jappl.1988.65.2.657

11.	Artal R, Masaki DI, Khodiguian N, Romem Y, Rutherford SE, Wiswell RA. Exercise prescription in pregnancy: weight-bearing versus non-weight-bearing exercise. *Am J Obstet Gynecol*. 1989;161(6 Pt 1):1464-1469. doi:10.1016/0002-9378(89)90905-8

12.	Hume RFFJ, Bowie JDD, McCoy C, et al. Fetal umbilical artery Doppler response to graded maternal aerobic exercise and subsequent maternal mean arterial blood pressure: predictive value for pregnancy-induced hypertension. *Am J Obstet Gynecol*. 1990;163(3):826-829. doi:10.1016/0002-9378(90)91077-p

13.	Sady MA, Haydon BB, Sady SP, Carpenter MW, Thompson PD, Coustan DR. Cardiovascular response to maximal cycle exercise during pregnancy and at two and seven months post partum. *Am J Obstet Gynecol*. 1990;162(5):1181-1185. doi:10.1016/0002-9378(90)90012-v

14.	Field SK, Bell SG, Cenaiko DF, Whitelaw WA. Relationship between inspiratory effort and breathlessness in pregnancy. *J Appl Physiol*. 1991;71(5):1897-1902. doi:10.1152/jappl.1991.71.5.1897

15.	Rafla NM, Beazely JM. The effect of maternal exercise on fetal umbilical artery waveforms. *Eur J Obstet Gynecol Reprod Biol*. 1991;40(2):119-122. doi:10.1016/0028-2243(91)90102-Q

16.	Bung P, Huch R, Huch A. *Maternal and Fetal Heart Rate Patterns: A Pregnant Athlete during Training and Laboratory Exercise Tests; a Case Report*. Vol 39.; 1991. Accessed November 27, 2018. https://www.ejog.org/article/0028-2243(91)90143-9/pdf

17.	Young JC, Treadway JL. The effect of prior exercise on oral glucose tolerance in late gestational women. *Eur J Appl Physiol Occup Physiol*. 1992;64(5):130-133. doi:10.1136/bmj.39546.498796.34

18.	Clapp JF, Little KD, Capeless EL. Fetal heart rate response to sustained recreational exercise. *Am J Obstet Gynecol*. 1993;168(1):198-206. doi:10.1016/S0002-9378(12)90914-X

19.	Lotgering FK, Struijk PC, Doorn MBVAN, Spinnewijn WEM, Wallenburg HCS. Anaerobic threshold and respiratory compensation in pregnant women. *J Appl Physiol*. 1995;85(6):1772-1777. http://www.ncbi.nlm.nih.gov/pubmed/7649911

20.	Artal R, Fortunato V, Welton A, et al. A comparison of cardiopulmonary adaptations to exercise in pregnancy at sea level and altitude. *Am J Obstet Gynecol*. 1995;172(4):1170-1180. doi:10.1016/0002-9378(95)91475-7

21.	O’Neill ME. Maternal rectal temperature and fetal heart rate responses to upright cycling in late pregnancy. *Br J Sports Med*. 1996;30(1):32-35. doi:10.1136/bjsm.30.1.32

22.	Soultanakis HN, Artal R, Wiswell RA. Prolonged exercise in pregnancy: glucose homeostasis, ventilatory and cardiovascular responses. *Semin Perinatol*. 1996;20(4):315-327. doi:10.1016/s0146-0005(96)80024-3

23.	Manders MA. AM, Sonder GJ. JB, Mulder EJ. JH, Visser GH. HA. The effects of maternal exercise on fetal heart rate and movement patterns. *EARLY Hum Dev*. 1997;48(3):237-247. doi:10.1016/S0378-3782(96)01858-0

24.	Kemp JG, Greer FA, Wolfe LA. Acid-base regulation after maximal exercise testing in late gestation. *J Appl Physiol*. 1997;83(2):644-651.

25.	Mcgrath MJ, Wolfe LA, Preston RJ, Burggraf GW, Mcgrath MJ. Effects of pregnancy and chronic exercise on maternal cardiac structure and function. *Artic Can J Physiol Pharmacol*. Published online 1999. doi:10.1139/cjpp-77-11-909

26.	Brenner IKM, Wolfe LA, Monga M, McGrath MJ. Physical conditioning effects on fetal heart rate responses to graded maternal exercise. *Med Sci Sports Exerc*. 1999;31(6):792-799. doi:10.1097/00005768-199906000-00006

27.	MacPhail A, Davies GA., Victory R, Wolfe LA. Maximal exercise testing in late gestation: fetal responses. *Obstet Gynecol*. 2000;96(4):565-570. doi:10.1016/S0029-7844(00)00940-6

28.	Heenan AP, Wolfe LA, Davies GA. Maximal exercise testing in late gestation: maternal responses. *Obstet Gynecol*. 2001;97(1):127-134. doi:10.1016/S0029-7844(00)01089-9

29.	Kennelly MM, Geary M, McCaffrey N, McLoughlin P, Staines A, McKenna P. Exercise-related changes in umbilical and uterine artery waveforms as assessed by Doppler ultrasound scans. *Am J Obstet Gynecol*. 2002;187(3):661-666. doi:10.1067/MOB.2002.125741

30.	Heenan AP, Wolfe LA. Plasma osmolality and the strong ion difference predict respiratory adaptations in pregnant and nonpregnant women. *Can J Physiol Pharmacol*. 2003;81(9):839-847. doi:10.1139/y03-072

31.	Wolfe LA, Heenan AP, Bonen A. Aerobic conditioning effects on substrate responses during graded cycling in pregnancy. *Can J Physiol Pharmacol*. 2003;81(7):696-703. doi:10.1139/y03-059

32.	Lindqvist PG, Marsal K, Merlo J, Pirhonen JP. Thermal response to submaximal exercise before, during and after pregnancy: a longitudinal study. *J Matern neonatal Med Off J Eur Assoc Perinat Med Fed Asia Ocean Perinat Soc Int Soc Perinat Obstet*. 2003;13(3):152-156. doi:10.1080/jmf.13.3.152.156

33.	Lynch A-MM, McDonald S, Magann EF, et al. Effectiveness and safety of a structured swimming program in previously sedentary women during pregnancy. *J Matern Neonatal Med*. 2003;14(3):163-169. doi:10.1080/jmf.14.3.163.169

34.	Heenan AP, Wolfe LA, Davies GAL, McGrath MJ. Effects of human pregnancy on fluid regulation responses to short-term exercise. *J Appl Physiol*. 2003;95(6):2321-2327. doi:10.1152/japplphysiol.00984.2002

35.	Pirhonen JP, Lindqvist PG, Marsal K. A longitudinat study of maternal oxygen saturation during short-term submaximal exercise. *Clin Physiol Funct Imaging*. 2003;23(1):37-41. doi:10.1046/j.1475-097X.2003.00467.x

36.	Kardel KR. Effects of intense training during and after pregnancy in top-level athletes. *Scand J Med Sci Sport*. 2005;15(2):79-86. doi:10.1111/j.1600-0838.2004.00426.x

37.	McAuley SE, Jensen D, McGrath MJ, Wolfe LA. Effects of human pregnancy and aerobic conditioning on alveolar gas exchange during exercise. *Can J Physiol Pharmacol*. 2005;83(7):625-633. doi:10.1139/y05-054

38.	Weissgerber TL, Wolfe LA, Hopkins WG, Davies GALL. Serial respiratory adaptations and an alternate hypothesis of respiratory control in human pregnancy. *Respir Physiol Neurobiol*. 2006;153(1):39-53. doi:10.1016/j.resp.2005.09.004

39.	Jensen D, Webb KA, Wolfe LA, O’Donnell DE. Effects of human pregnancy and advancing gestation on respiratory discomfort during exercise. *Respir Physiol Neurobiol*. 2007;156(1):85-93. doi:10.1016/J.RESP.2006.08.004

40.	Jensen D, Webb KA, Davies GAL, O’Donnell DE. Mechanical ventilatory constraints during incremental cycle exercise in human pregnancy: implications for respiratory sensation. *J Physiol*. 2008;586(19):4735-4750. doi:10.1113/jphysiol.2008.158154

41.	Kardel KR, Johansen BBBB, Voldner N, Iversen PO, Henriksen T. Association between aerobic fitness in late pregnancy and duration of labor in nulliparous women. *ACTA Obstet Gynecol Scand*. 2009;88(8):948-952. doi:10.1080/00016340903093583

42.	Ong MJ, Guelfi KJ, Hunter T, Wallman KE, Fournier PA, Newnham JP. Supervised home-based exercise may attenuate the decline of glucose tolerance in obese pregnant women. *Diabetes Metab*. 2009;35(5):418-421. doi:10.1016/J.DIABET.2009.04.008

43.	Thorell E, Svärdsudd K, Andersson K, Kristiansson P. Moderate impact of full-term pregnancy on estimated peak oxygen uptake, physical activity and perceived health. *Acta Obstet Gynecol Scand*. 2010;89(9):1140-1148. doi:10.3109/00016349.2010.487894

44.	Vega SR, Kleinert J, Sulprizio M, Hollmann W, Bloch W, Strueder HK. Responses of serum neurotrophic factors to exercise in pregnant and postpartum women. *Psychoneuroendocrinology*. 2011;36(2):220-227. doi:10.1016/j.psyneuen.2010.07.012

45.	Thorell E, Goldsmith L, Weiss G, Kristiansson P. Physical fitness, serum relaxin and duration of gestation. *BMC Pregnancy Childbirth*. 2015;15(1):1-7. doi:10.1186/s12884-015-0607-z

46.	Kim J-H, Roberge RJ, Powell JB. Effect of external airflow resistive load on postural and exercise-associated cardiovascular and pulmonary responses in pregnancy: a case control study. *BMC Pregnancy Childbirth*. 2015;15(1):45. doi:10.1186/s12884-015-0474-7

47.	Nakagaki A, Inami T, Minoura T, Baba R, Iwase S, Sato M. Differences in autonomic neural activity during exercise between the second and third trimesters of pregnancy. *J Obstet Gynaecol Res*. 2016;42(8):951-959. doi:10.1111/jog.12990

48.	Jędrzejko M, Nowosielski K, Poręba R, Ulman-Włodarz I, Bobiński R. Physical efficiency and activity energy expenditure in term pregnancy females measured during cardiopulmonary exercise tests with a supine cycle ergometer. *J Matern Neonatal Med*. 2016;29(23):3800-3805. doi:10.3109/14767058.2016.1147550

49.	Sussman D, Saini BS, Schneiderman JE, et al. Uterine artery and umbilical vein blood flow are unaffected by moderate habitual physical activity during pregnancy. *Prenat Diagn*. 2019;39(11):976-985. doi:10.1002/pd.5517

50.	Purdy GM, James MA, Wakefield PK, et al. Maternal cardioautonomic responses during and following exercise throughout pregnancy. *Appl Physiol Nutr Metab*. 2019;44(3):263-270. doi:10.1139/apnm-2018-0397

51.	Bilodeau J-F, Bisson M, Larose J, et al. Physical fitness is associated with prostaglandin F-2 alpha isomers during pregnancy. *PROSTAGLANDINS Leukot Essent Fat ACIDS*. 2019;145:7-14. doi:10.1016/j.plefa.2019.05.001

52.	Matenchuk BA, James M, Skow RJ, et al. Longitudinal study of cerebral blood flow regulation during exercise in pregnancy. *J Cereb BLOOD FLOW Metab*. 2020;40(11):2278-2288. doi:10.1177/0271678X19889089

53.	Correa M da S, Catai AM, Milan-Mattos JC, Porta A, Driusso P. Is pelvic floor muscle training able to alter the response of cardiovascular autonomic modulation and provide a possible cardiovascular benefit to pregnant women? *Neurourol Urodyn*. 2020;39(8):2272-2283. doi:10.1002/nau.24481

54.	Bijl RC, Cornette JMJ, van Der Ham K, et al. The physiological effect of early pregnancy on a woman’s response to a submaximal cardiopulmonary exercise test. *Physiol Rep*. 2020;8(21). doi:10.14814/phy2.14624

55.	Sibley L, Ruhling R, Ph D, Christensen C, Bolen T. Swimming and Physicalfitness Duringpregnancy. 1981;26(6):3-12.

56.	Lewis RD, Yates CY, Driskell JA. Riboflavin and thiamin status and birth outcome as a function of maternal aerobic exercise. *Am J Clin Nutr*. 1988;48(1):110-116. doi:10.1093/ajcn/48.1.110

57.	WINN HN, HESS O, GOLDSTEIN I, WACKERS F, HOBBINS JC. FETAL RESPONSES TO MATERNAL EXERCISE - EFFECT ON FETAL BREATHING AND BODY MOVEMENT. *Am J Perinatol*. 1994;11(4):263-266. doi:10.1055/s-2007-994588

58.	Marquez-Sterling S, Perry AC, Kaplan TA, Halberstein RA, Signorile JF. Physical and psychological changes with vigorous exercise in sedentary primigravidae. *Med Sci Sports Exerc*. 2000;32(1):58-62. doi:10.1097/00005768-200001000-00010

59.	Santos IA, Stein R, Fuchs SC, et al. Aerobic exercise and submaximal functional capacity in overweight pregnant women - A randomized trial. *Obstet Gynecol*. 2005;106(2):243-249. doi:10.1097/01.AOG.0000171113.36624.86

60.	Yeo S, Ronis DL, Antonakos CL, Roberts K, Hayashi R. Need for population specific validation of a portable metabolic testing system: a case of sedentary pregnant women. *J Nurs Meas*. 2005;13(3):207-218. doi:10.1891/jnum.13.3.207

61.	Mottola MF, Davenport MH, Brun CR, Inglis SD, Charlesworth S, Sopper MM. V̇O2peak prediction and exercise prescription for pregnant women. *Med Sci Sports Exerc*. 2006;38(8):1389-1395. doi:10.1249/01.mss.0000228940.09411.9c

62.	Davenport MH, Charlesworth S, Vanderspank D, Sopper MM, Mottola MF. Development and validation of exercise target heart rate zones for overweight and obese pregnant women. *Appl Physiol Nutr Metab*. 2008;33(5):984-989. doi:10.1139/H08-086

63.	de Oliveria Melo AS, Silva JLPJLP, Tavares JS, Barros VO, Leite DFBB, Amorim MMRR. Effect of a Physical Exercise Program During Pregnancy on Uteroplacental and Fetal Blood Flow and Fetal Growth A Randomized Controlled Trial. *Obstet Gynecol*. 2012;120(2, 1):302-310. doi:10.1097/AOG.0b013e31825de592

64.	Ruchat S-M, Davenport M, Giroux I, et al. Walking Program of Low or Vigorous Intensity During Pregnancy Confers an Aerobic Benefit. *Int J Sports Med*. 2012;33(08):661-666. doi:10.1055/s-0032-1304635

65.	Szymanski LM, Satin AJ. Strenuous exercise during pregnancy: is there a limit? *Am J Obstet Gynecol*. 2012;207(3):179.e1-179.e6. doi:10.1016/j.ajog.2012.07.021

66.	Salvesen K a. Å, Hem E, Sundgot-Borgen J. Fetal wellbeing may be compromised during strenuous exercise among pregnant elite athletes. *Br J Sports Med*. 2012;46(4):279-283. doi:10.1136/bjsm.2010.080259

67.	Mottola MF, Inglis S, Brun CR, Hammond J-A. Physiological and metabolic responses of late pregnant women to 40 min of steady-state exercise followed by an oral glucose tolerance perturbation. *J Appl Physiol*. 2013;115(5):597-604. doi:10.1152/japplphysiol.00487.2013

68.	Bisson M, Alméras N, Plaisance J, et al. Maternal fitness at the onset of the second trimester of pregnancy: Correlates and relationship with infant birth weight. *Pediatr Obes*. 2013;8(6):464-474. doi:10.1111/j.2047-6310.2012.00129.x

69.	LeMoyne EL, Curnier D, Ellemberg D. Pregnancy and cognition: Deficits in inhibition are unrelated to changes in fitness. *J Clin Exp Neuropsychol*. 2014;36(2):178-185. doi:10.1080/13803395.2013.875520

70.	Bisson M, Rhéaume C, Bujold E, Tremblay A, Marc I. Modulation of blood pressure response to exercise by physical activity and relationshipwith resting blood pressure during pregnancy. *J Hypertens*. 2014;32(7):1450-1457. doi:10.1097/HJH.0000000000000185

71.	Marshall MR, Pivarnik JM. Perceived Exertion of Physical Activity During Pregnancy. *J Phys Act Heal*. 2015;12(7):1039-1043. doi:10.1123/jpah.2013-0458

72.	Perales M, Santos-Lozano A, Ruiz JR, Lucia A, Barakat R. Benefits of aerobic or resistance training during pregnancy on maternal health and perinatal outcomes: A systematic review. *EARLY Hum Dev*. 2016;94:43-48. doi:10.1016/j.earlhumdev.2016.01.004

73.	Hesse CM, Tinius RA, Pitts BC, et al. Assessment of endpoint criteria and perceived barriers during maximal cardiorespiratory fitness testing among pregnant women. *J Sports Med Phys Fitness*. 2018;58(12):1844-1851. doi:10.23736/S0022-4707.17.07750-7

74.	Baena-García L, Coll-Risco I, Ocón-Hernández O, et al. Association of objectively measured physical fitness during pregnancy with maternal and neonatal outcomes. The GESTAFIT Project. *PLoS One*. 2020;15(2):e0229079. doi:10.1371/journal.pone.0229079

75.	Dobson KL, da Silva DF, Dervis S, Mohammad S, Nagpal TS, Adamo KB. Physical activity and gestational weight gain predict physiological and perceptual responses to exercise during pregnancy. *Birth Defects Res*. Published online September 23, 2020:bdr2.1808. doi:10.1002/bdr2.1808

76.	Da Silva EG, De Godoy I, De Oliveira Antunes LC, Da Silva EG, Peraçoli JC. Respiratory parameters and exercise functional capacity in preeclampsia. *Hypertens Pregnancy*. 2010;29(3):301-309. doi:10.3109/10641950902779271

77.	Ramírez-Vélez R, Aguilar de Plata AC, Escudero MM, et al. Influence of regular aerobic exercise on endothelium-dependent vasodilation and cardiorespiratory fitness in pregnant women. *J Obstet Gynaecol Res*. 2011;37(11):1601-1608. doi:10.1111/j.1447-0756.2011.01582.x

78.	Hjorth MF, Kloster S, Girma T, et al. Level and intensity of objectively assessed physical activity among pregnant women from urban Ethiopia. *BMC Pregnancy Childbirth*. 2012;12(1):154. doi:10.1186/1471-2393-12-154

79.	Price BB, Amini SB, Kappeler K. Exercise in Pregnancy: Effect on Fitness and Obstetric Outcomes-A Randomized Trial. *Med Sci Sport Exerc*. 2012;44(12):2263-2269. doi:10.1249/MSS.0b013e318267ad67

80.	Radzikowska E, Wiatr E, Franczuk M, Bestry I, Roszkowski-Sliz K. Lung Function in Pregnancy in Langerhans Cell Histiocytosis. In: Pokorski, M, ed. *PULMONARY DISORDERS AND THERAPY*. Vol 1023. Advances in Experimental Medicine and Biology. ; 2018:73-83. doi:10.1007/5584_2017_72

81.	Oviedo-Caro MA, Bueno-Antequera J, Munguía-Izquierdo D. Explanatory factors and levels of health-related quality of life among healthy pregnant women at midpregnancy: A cross-sectional study of The PregnActive Project. *J Adv Nurs*. 2018;74(12):2766-2776. doi:10.1111/jan.13787

82.	Dennis AT, Salman M, Paxton E, et al. Resting Hemodynamics and Response to Exercise Using the 6-Minute Walk Test in Late Pregnancy: An International Prospective Multicentre Study. *Anesth Analg*. 2019;129(2):450-457. doi:10.1213/ANE.0000000000003818

83.	Amola M, Pawara S, Kalra S. Effect of Inspiratory Muscle Training and Diaphragmatic Breathing Exercises on Dyspnea, Pulmonary Functions, Fatigue and Functional Capacity in Pregnancy during Third Trimester. *J Clin DIAGNOSTIC Res*. 2019;13(8):YC1-YC4. doi:10.7860/JCDR/2019/41732.13037

84.	Birnbaumer P, Dietz P, Watson ED, et al. Absolute Accelerometer-Based Intensity Prescription Compared to Physiological Variables in Pregnant and Nonpregnant Women. *Int J Environ Res Public Health*. 2020;17(16). doi:10.3390/ijerph17165651

85.	Dibblee L, Graham TEE. A longitudinal study of changes in aerobic fitness, body composition, and energy intake in primigravid patients. *Am J Obstet Gynecol*. 1983;147(8):908-914. doi:10.1016/0002-9378(83)90244-2

86.	WILLIAMS A, Reilly T, CAMPBELL I, Sutherst J. Investigation of changes in responses to exercise and in mood during pregnancy. *Ergonomics*. 1988;31(11):1539-1549. doi:10.1080/00140138808966803

87.	Melzer K, Schutz Y, Soehnchen N, et al. Effects of recommended levels of physical activity on pregnancy outcomes. *Am J Obstet Gynecol*. 2010;202(3):266.e1-6. doi:10.1016/j.ajog.2009.10.876

88.	Baker PN, Johnson IR. The use of the hand-grip test for predicting pregnancy-induced hypertension. *Eur J Obstet Gynecol Reprod Biol*. 1994;56(3):169-172. doi:10.1016/0028-2243(94)90165-1

89.	Rogers MS, Tomlinson B. Change in cardiovascular indices with position and isometric exercise throughout pregnancy: Assessment by impedance cardiography and oscillometric sphygmomanometry. *Hypertens Pregnancy*. 1998;17(2):191-202. doi:10.3109/10641959809006075

90.	Feiner B, Weksler R, Ohel G, Degani S. The influence of maternal exercise on placental blood flow measured by Simultaneous Multigate Spectral Doppler Imaging (SM-SDI). *Ultrasound Obstet Gynecol*. 2000;15(6):498-501. doi:10.1046/j.1469-0705.2000.00146.x

91.	Gutke A, Östgaard HC, Öberg B. Association between muscle function and low back pain in relation to pregnancy. *J Rehabil Med*. 2008;40(4):304-311. doi:10.2340/16501977-0170

92.	O’Connor PJ, Poudevigne MS, Cress ME, Motl RW, Clapp JF. Safety and efficacy of supervised strength training adopted in pregnancy. *J Phys Act Health*. 2011;8(3):309-320. Accessed November 12, 2018. http://www.ncbi.nlm.nih.gov/pubmed/21487130

93.	Atay E, Başalan İz F. Investigation of the effect of changes in muscle strength in gestational age upon fear of falling and quality of life. *Turkish J Med Sci*. 2015;45(4):977-983. doi:10.3906/sag-1404-9

94.	Petrov Fieril K, Glantz A, Fagevik Olsen M. The efficacy of moderate-to-vigorous resistance exercise during pregnancy: A randomized controlled trial. *Acta Obstet Gynecol Scand*. 2015;94(1):35-42. doi:10.1111/aogs.12525

95.	Wickboldt N, Savoldelli G, Rehberg-Klug B. Continuous assessment of labour pain using handgrip force. *Pain Res Manag*. 2015;20(3):159-163. doi:10.1155/2015/281976

96.	Kalliokoski P, Rodhe N, Bergqvist Y, Löfvander M. Long-term adherence and effects on grip strength and upper leg performance of prescribed supplemental vitamin D in pregnant and recently pregnant women of Somali and Swedish birth with 25-hydroxyvitamin D deficiency: a before-and-after treatment study. *BMC Pregnancy Childbirth*. 2016;16(1):353. doi:10.1186/s12884-016-1117-3

97.	Ngaka TCC, Coetzee JFF, Dyer RAA. The Influence of Body Mass Index on Sensorimotor Block and Vasopressor Requirement During Spinal Anesthesia for Elective Cesarean Delivery. *Anesth Analg*. 2016;123(6):1527-1534. doi:10.1213/ANE.0000000000001568

98.	Rodriguez-Diaz L, Ruiz-Frutos C, Maria Vazquez-Lara J, Ramirez-Rodrigo J, Villaverde-Gutierrez C, Torres-Luque G. Effectiveness of a physical activity programme based on the Pilates method in pregnancy and labour. *Enferm Clin*. 2017;27(5):271-277. doi:10.1016/j.enfcle.2017.05.007

99.	Żelaźniewicz A, Pawłowski B. Maternal hand grip strength in pregnancy, newborn sex and birth weight. *Early Hum Dev*. 2018;119:51-55. doi:10.1016/j.earlhumdev.2018.03.004

100.	Takeda K, Yoshikata H, Imura M. Do Squat Exercises With Weight Shift During Pregnancy Improve Postural Control? *Int J WOMENS Heal Reprod Sci*. 2019;7(1):10-16. doi:10.15296/ijwhr.2019.02

101.	Yenişehir S, Çıtak Karakaya İ, Sivaslıoğlu AA, Özen Oruk D, Karakaya MG. Reliability and validity of Five Times Sit to Stand Test in pregnancy-related pelvic girdle pain. *Musculoskelet Sci Pract*. 2020;48:102157. doi:10.1016/j.msksp.2020.102157

102.	Gilleard W, Crosbie J, Smith R. Effect of pregnancy on trunk range of motion when sitting and standing. *Acta Obstet Gynecol Scand*. 2002;81(11):1011-1020. doi:10.1034/j.1600-0412.2002.811104.x

103.	Marnach ML, Ramin KD, Ramsey PS, Song S-W, Stensland JJ, An K-N. Characterization of the relationship between joint laxity and maternal hormones in pregnancy. *Obstet Gynecol*. 2003;101(2):331-335. doi:10.1016/s0029-7844(02)02447-x

104.	Garshasbi A, Faghih Zadeh S. The effect of exercise on the intensity of low back pain in pregnant women. *Int J Gynecol Obstet*. 2005;88(3):271-275. doi:10.1016/j.ijgo.2004.12.001

105.	Lindgren A, Kristiansson P. Finger joint laxity, number of previous pregnancies and pregnancy induced back pain in a cohort study. *BMC Pregnancy Childbirth*. 2014;14(1):61. doi:10.1186/1471-2393-14-61

106.	Cherni Y, Desseauve D, Decatoire A, et al. Evaluation of ligament laxity during pregnancy. *J Gynecol Obstet Hum Reprod*. 2019;48(5):351-357. doi:10.1016/j.jogoh.2019.02.009

107.	Butler EE, Colon I, Druzin ML, Rose J. Postural equilibrium during pregnancy: Decreased stability with an increased reliance on visual cues. *Am J Obstet Gynecol*. 2006;195(4):1104-1108. doi:10.1016/j.ajog.2006.06.015

108.	Ribas I S, Guirro ECO. Analysis of plantar pressure and postural balance during different phases of pregnancy. *BRAZILIAN J Phys Ther*. 2007;11(5):391-396.

109.	Nagai M, Isida M, Saitoh J, Hirata Y, Natori H, Wada M. Characteristics of the control of standing posture during pregnancy. *Neurosci Lett*. 2009;462(2):130-134. doi:10.1016/j.neulet.2009.06.091

110.	Oliveira LF, Vieira TMM, Macedo AR, Simpson DM, Nadal J. Postural sway changes during pregnancy: A descriptive study using stabilometry. *Eur J Obstet Gynecol Reprod Biol*. 2009;147(1):25-28. doi:10.1016/J.EJOGRB.2009.06.027

111.	Karadag-Saygi E, Unlu-Ozkan F, Basgul A. Plantar Pressure and Foot Pain in the Last Trimester of Pregnancy. *FOOT ANKLE Int*. 2010;31(2):153-157. doi:10.3113/FAI.2010.0153

112.	Yu Y, Chung HC, Hemingway L, Stoffregen TA. Standing body sway in women with and without morning sickness in pregnancy. *Gait Posture*. 2013;37(1):103-107. doi:10.1016/j.gaitpost.2012.06.021

113.	Ersal, T., McCrory, J. L., & Sienko KH, Ersal T, McCrory JL, Sienko KH. Theoretical and experimental indicators of falls during pregnancy as assessed by postural perturbations. *Gait Posture*. 2014;39(1):218-223. doi:10.1016/j.gaitpost.2013.07.011

114.	Opala-Berdzik A, Bacik B, Markiewicz A, et al. Comparison of static postural stability in exercising and non-exercising women during the perinatal period. *Med Sci Monit*. 2014;20:1865-1870. doi:10.12659/MSM.890846

115.	Opala-Berdzik A, Błaszczyk JW, Bacik B, et al. Static Postural Stability in Women during and after Pregnancy: A Prospective Longitudinal Study. McCrory JL, ed. *PLoS One*. 2015;10(6):e0124207. doi:10.1371/journal.pone.0124207

116.	Öztürk G, Geler Külcü D, Aydoğ E, Kaspar Ç, Uğurel B. Effects of lower back pain on postural equilibrium and fall risk during the third trimester of pregnancy. *J Matern Neonatal Med*. 2016;29(8):1358-1362. doi:10.3109/14767058.2015.1049148

117.	Shibayama Y, Kuwata T, Yamaguchi J, et al. Changes in standing body sway of pregnant women after long-term bed rest. *J Obstet Gynaecol (Lahore)*. 2016;36(4):479-482. doi:10.3109/01443615.2015.1086983

118.	Takeda K, Yoshikata H, Imura M. Changes in Posture Control of Women That Fall During Pregnancy. Published online 2018. doi:10.15296/ijwhr.2018.43

119.	Moreira LS, Elias LA, Gomide AB, Vieira MF, Do Amaral WN. A longitudinal assessment of myoelectric activity, postural sway, and low-back pain during pregnancy. *Acta Bioeng Biomech*. 2017;Vol. 19(nr 3). doi:10.5277/ABB-00753-2016-02

120.	Opala-Berdzik A, Błaszczyk JW, Świder D, Cieślińska-Świder J. Trunk forward flexion mobility in reference to postural sway in women after delivery: A prospective longitudinal comparison between early pregnancy and 2- and 6-month postpartum follow-ups. *Clin Biomech*. 2018;56:70-74. doi:10.1016/j.clinbiomech.2018.05.009

121.	Catena RD, Campbell N, Wolcott WC, Rothwell SA. Anthropometry, standing posture, and body center of mass changes up to 28 weeks postpartum in Caucasians in the United States. *Gait Posture*. 2019;70:196-202. doi:10.1016/j.gaitpost.2019.03.009

122.	Fontana Carvalho AP, Dufresne SS, de Oliveira M, et al. Effects of lumbar stabilization and muscular stretching on pain, disabilities, postural control and muscle activation in pregnant woman with low back pain. *Eur J Phys Rehabil Med*. 2020;56(3):297-306. doi:10.23736/S1973-9087.20.06086-4

123.	Valerio PM, Goncalves VE, Zordao CC, Rezende MS, Dantas Moises EC, de Olveira Guirro EC. Influence of type 1 diabetes on the postural control of women in the third gestational trimester. *Clin Biomech*. 2020;77. doi:10.1016/j.clinbiomech.2020.105062

124.	Davies J, Fernando R, McLeod A, Verma S, Found P. Postural stability following ambulatory regional analgesia for labor. *Anesthesiology*. 2002;97(6):1576-1581. doi:10.1097/00000542-200212000-00033

125.	McCrory JLL, Chambers AJJ, Daftary A, Redfern MSS. Dynamic postural stability during advancing pregnancy. *J Biomech*. 2010;43(12):2434-2439. doi:10.1016/j.jbiomech.2009.09.058

126.	Branco M, Santos-Rocha R, Aguiar L, Vieira F, Veloso A. Kinematic analysis of gait in the second and third trimesters of pregnancy. *J Pregnancy*. 2013;2013:718095. doi:10.1155/2013/718095

127.	Cakmak B, Inanir A, Nacar MC, Filiz B. The Effect of Maternity Support Belts on Postural Balance in Pregnancy. *PM&R*. 2014;6(7):624-628. doi:10.1016/j.pmrj.2013.12.012

128.	Inanir A, Cakmak B, Hisim Y, Demirturk F. Evaluation of postural equilibrium and fall risk during pregnancy. *Gait Posture*. 2014;39(4):1122-1125. doi:10.1016/j.gaitpost.2014.01.013

129.	Wu X, Yeoh HT. Intrinsic factors associated with pregnancy falls. *Workplace Health Saf*. 2014;62(10):403-408. doi:10.3928/21650799-20140902-04

130.	Forczek W, Staszkiewicz R. Changes of kinematic gait parameters due to pregnancy. *Acta Bioeng Biomech*. 2012;14(4):113-119. doi:10.5277/abb120413

131.	Takeda K. A Kinesiological Analysis of the Stand-to-Sit during the Third Trimester. *J Phys Ther Sci*. 2012;24(7):621-624. doi:10.1589/jpts.24.621

132.	Gottschall JS, Sheehan RC, Downs DS. Pregnant women exaggerate cautious gait patterns during the transition between level and hill surfaces. *J Electromyogr Kinesiol Off J Int Soc Electrophysiol Kinesiol*. 2013;23(5):1237-1242. doi:10.1016/j.jelekin.2013.04.011

133.	McCrory JL, Chambers AJ, Daftary A, Redfern MS. The pregnant “waddle”: An evaluation of torso kinematics in pregnancy. *J Biomech*. 2014;47(12):2964-2968. doi:10.1016/j.jbiomech.2014.07.009

134.	Krkeljas Z. Changes in gait and posture as factors of dynamic stability during walking in pregnancy. *Hum Mov Sci*. 2018;58:315-320. doi:10.1016/j.humov.2017.12.011

135.	Catena RD, Bailey JP, Campbell N, Music HE. Stand-to-sit kinematic changes during pregnancy correspond with reduced sagittal plane hip motion. *Clin Biomech*. 2019;67:107-114. doi:10.1016/j.clinbiomech.2019.05.014

136.	Catena RD, Campbell N, Werner AL, Iverson KM. Anthropometric Changes During Pregnancy Provide Little Explanation of Dynamic Balance Changes. *J Appl Biomech*. 2019;35(3):232-239. doi:10.1123/jab.2018-0345

137.	Forczek W, Ivanenko Y, Curyło M, et al. Progressive changes in walking kinematics throughout pregnancy-A follow up study. *Gait Posture*. 2019;68:518-524. doi:10.1016/j.gaitpost.2019.01.004

138.	Forczek W, Masłoń A, Frączek B, Curyło M, Salamaga M, Suder A. Does the first trimester of pregnancy induce alterations in the walking pattern? *PLoS One*. 2019;14(1):e0209766. doi:10.1371/journal.pone.0209766

139.	Catena RD, Bailey JP, Campbell N, Stewart BC, Marion SJ. Correlations between joint kinematics and dynamic balance control during gait in pregnancy. *Gait Posture*. 2020;80:106-112. doi:10.1016/j.gaitpost.2020.05.025

140.	Gimunova M, Zvonar M, Sebera M, Turcinek P, Kolarova K. Special footwear designed for pregnant women and its effect on kinematic gait parameters during pregnancy and postpartum period. *PLoS One*. 2020;15(5). doi:10.1371/journal.pone.0232901

141.	McCrory JL, Chambers AJ, Daftary A, Redfern MS. Torso kinematics during gait and trunk anthropometry in pregnant fallers and non-fallers. *Gait Posture*. 2020;76:204-209. doi:10.1016/j.gaitpost.2019.11.012

142.	Rothwell SA, Eckland CB, Campbell N, Connolly CP, Catena RD. An analysis of postpartum walking balance and the correlations to anthropometry. *Gait Posture*. 2020;76:270-276. doi:10.1016/j.gaitpost.2019.12.017

143.	Forczek W, Ivanenko Y, Salamaga M, et al. Pelvic movements during walking throughout gestation - the relationship between morphology and kinematic parameters. *Clin Biomech*. 2020;71:146-151. doi:10.1016/j.clinbiomech.2019.11.001

144.	Sawa R, Doi T, Asai T, Watanabe K, Taniguchi T, Ono R. Differences in trunk control between early and late pregnancy during gait. *Gait Posture*. 2015;42(4):455-459. doi:10.1016/j.gaitpost.2015.07.058

145.	Opala-Berdzik A, Bacik B, Markiewicz A, et al. Comparison of Static Postural Stability in Exercising and Non-Exercising Women During the Perinatal Period. *Med Sci Monit*. 2014;20:1865-1870. doi:10.12659/MSM.890846

146.	Evensen NM, Kvale A, Braekken IH, Kvåle A, Braekken IH. Reliability of the Timed Up and Go test and Ten-Metre Timed Walk Test in Pregnant Women with Pelvic Girdle Pain. *Physiother Res Int*. 2015;20(3):158-165. doi:10.1002/pri.1609

147.	Evensen NM, Kvåle A, Brækken IH. Convergent validity of the Timed Up and Go Test and Ten-metre Timed Walk Test in pregnant women with pelvic girdle pain. *Man Ther*. 2016;21:94-99. Accessed November 10, 2018. https://www.sciencedirect.com/science/article/pii/S1356689X15001319

148.	Christensen L, Vollestad NK, Veierod MB, Stuge B, Cabri J, Robinson HS. The Timed Up & Go test in pregnant women with pelvic girdle pain compared to asymptomatic pregnant and non-pregnant women. *Musculoskelet Sci Pract*. 2019;43:110-116. doi:10.1016/j.msksp.2019.03.006
